# Single cell long read whole genome sequencing reveals somatic transposon activity in human brain

**DOI:** 10.1101/2024.11.11.24317113

**Authors:** Michal B Izydorczyk, Ester Kalef-Ezra, Dominic W Horner, Xinchang Zheng, Nadine Holmes, Marco Toffoli, Zeliha Gozde Sahin, Yi Han, Heer H Mehta, Donna M Muzny, Adam Ameur, Fritz J Sedlazeck, Christos Proukakis

**Affiliations:** Human Genome Sequencing Center, Baylor College of Medicine, Houston, TX, USA; Department of Clinical and Movement Neurosciences, Royal Free Campus, Queen Square Institute of Neurology, University College London, London, UK; Aligning Science Across Parkinson’s (ASAP) Collaborative Research Network, Chevy Chase, MD, USA; DeepSeq, Nottingham, UK; Science for Life Laboratory, Department of Immunology, Genetics and Pathology, Uppsala University, Uppsala, Sweden; Department of Computer Science, Rice University, 6100 Main Street, Houston, TX, USA

## Abstract

The advent of single cell DNA sequencing revealed astonishing dynamics of genomic variability, but failed at characterizing smaller to mid size variants that on the germline level have a profound impact. In this work we discover novel dynamics in three brains utilizing single cell long-read sequencing. This provides key insights into the dynamic of the genomes of individual cells and further highlights brain specific activity of transposable elements.

## Main

Single cell whole genome amplification (WGA) enables single cell whole genome sequencing (scWGS) typically carried out using short reads at low coverage^1^, which generally only detects Mb-scale CNVs, although identification of CNVs > 50kbp was reported^2^. In any case, many variants that would be expected such as Alu or LINE variants are missed. These transposable elements (TE) families are the most abundant and active transposons, collectively accounting for approximately 27% of the human genome^3^ and contributing to recombination in healthy neurons^4^ and neurodegenerative diseases^5–7^. Simultaneously, the advent of long-read sequencing enables the accurate detection of Alu or other transposon-mediated mutations^8^.

Long-read scWGS (scWGS-LR) was recently reported on T-cells, after WGA using isothermal Multiple Displacement Amplification (MDA) in droplets (dMDA), to assemble one genome of a single cell. However, it came at significant cost, and with limited completeness due to chimera and restricted amplicon size^9^. Nevertheless, that opens up the field for further exploration of whether a similar approach can provide novel insights into the genomic variation of single cells, and thus shed light into mosaic mechanisms with potential impact in disease. This is highly relevant since somatic mutations may contribute to neurodegenerative disorders such as synucleinopathies, which include Parkinson’s disease and Multiple System Atrophy (MSA). MSA is a sporadic disease of low heritability^10^ with some possible genetic predispositions^11^. We have already reported somatic CNVs (gains) of the *SNCA* gene in PD and MSA^12,13,14^, associated with regional and single cell pathology in MSA^15^, and Mb-scale single cell CNVs genome-wide in two MSA cases^13,16^.

In this study, we apply scWGS-LR as a proof of concept to reveal novel insights into the dynamic of variants across individual brain samples. To enable this, we have developed novel filtering methods and strategies to avoid known amplification biases. Furthermore, we are comparing the single cell long-read with bulk long-read and single cell short read to assess the variants we identified. Our results highlight multiple classes of variants on a single cell level including SNVs, small InDels and larger insertions and deletions. Particularly, we find variants containing sequences of transposable elements implicated in neurodegeneration and normal brain tissue mosaicism. Altogether, this leads to deeper insights into the variability of important genes but further highlights a notable transposon activity in the brain.

To assess this, we utilized dMDA to amplify DNA from a single cell, aiming to reduce amplification bias while maintaining a relatively long molecule length. This variation of MDA method compartmentalizes single cell DNA fragments into individual droplets, and was already shown to reduce sequencing coverage bias in blood cells^9^. We employed two different library preparations for all cells: T7 debranching, the standard method to remove displaced strands created by MDA; and the PCR rapid barcoding protocol (RBP), which creates linear molecules, albeit with limited length, on multiple ONT sequencing devices (see **Supplement Section 1**). We sequenced a total of 18 single cells across the cortex of three brains, two MSA (cingulate cortex) and one control (frontal cortex), where we previously performed Illumina scWGS on other cells using an hybrid WGA method (PicoPLEX)^13^ to enable a comparison. To avoid uneven genome coverage, we pooled and barcoded 6 single cells per ONT flow cell, obtaining 144M reads, with up-to 5.8M reads longer than 3kb, and some as long as 300kb, with the average N50 of 2.8kb (**Supplementary table 1**). Overall, this resulted in covering up-to ∼46% of the human genome at 5x coverage or higher across 6 single cells with ONT sequencing. This is only slightly less than Illumina scWGS of the same dMDA cells at ∼60% (**Figure 1A**). Thus, overall yielding enough long-read single cell coverage to identify small to mid size variants. Furthermore, to enable comparisons we sequenced each of the brain regions from the same donors using bulk ONT sequencing (metrics in **Supplementary table 2**).

**Figure 1.**
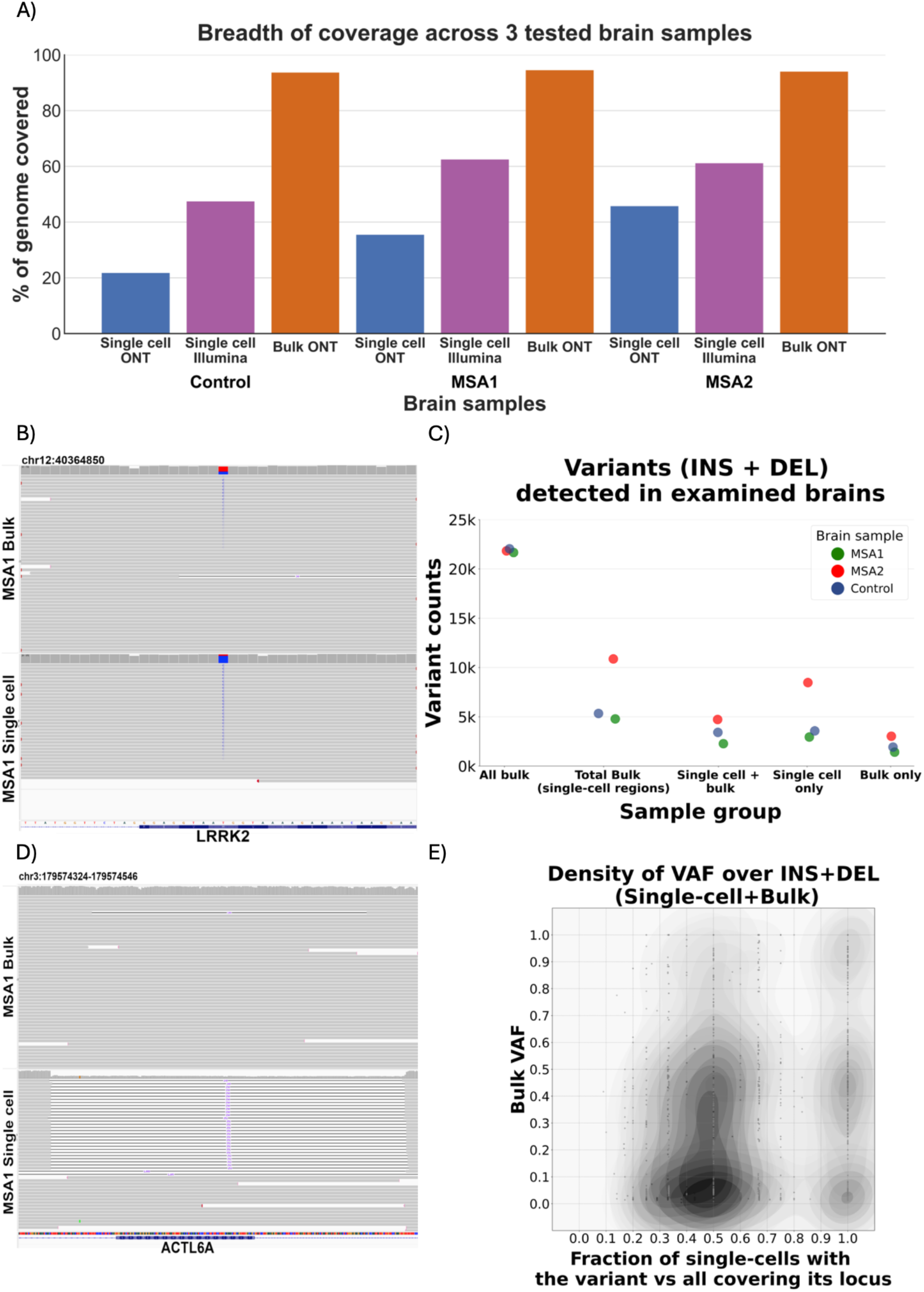
Overall metrics of genomic data obtained in scWGS experiments of studied brains **A)**. Breadth of coverage for each of the analyzed cell types, including long-read ONT sequencing and short-read scWGS Illumina sequencing of the same dMDA single-cell DNA. For single cells, analyzed regions are limited to >=5x depth of coverage, while for bulk all covered regions are included. **B)** Mosaic SNV detected in long-read scWGS and bulk ONT MSA1 sample located within an exon of LRRK2 - a gene important in monogenic Parkinson’s disease. **C)** SVs (insertions and deletions) detected across the 3 studied brains in long-read scWGS and bulk ONT samples. The first category shows all bulk variants across the genome, while the remaining 4 categories are limited to regions covered >=5x in single cell-samples and variants from those regions in corresponding bulk. **D)** Mosaic deletion detected in long-read MSA1 ONT single cell samples as well as in low frequency in corresponding long-read ONT bulk tissue sample. The deletion overlaps ACTL6A gene, encoding actin-related protein associated with Non-Specific Syndromic Intellectual Disability and Torticollis. **E)** A comparison of variant allele frequency (VAF) in long-read ONT bulk samples and the ratio of single cells where the variant is detected to the total number of single cells covering the variant locus in long-read ONT scWGS experiments. High VAF population variants were removed. Most of the remaining variants are mosaic in bulk, as represented by the heatmap concentration in the lower half of the chart.

To establish our filters and sequencing strategy we utilized the GIAB benchmark (GIAB SNV v4.2.1), performing ONT WGS of low-input HG002 sample after MDA amplification. Doing so we achieved an F-score of 93.4% for SNV/InDels (**Supplementary table 3**). Given these encouraging results, we called variants across single cell and bulk brain samples using the same strategy (see methods, **Supplementary table 4**). For bulk sequencing alone we obtained on average 4.72M SNV/InDels (**Supplementary table 2**). When comparing this to the single cell data, we observed similar numbers of SNV/InDels for regions that were covered >5x in both datasets as 1.43M SNVs/InDels in bulk and 1.35M SNVs/InDels for single cell data. Overall we observed a high overlap of 69.9% of bulk SNVs/InDels confirmed in single cell data (for examples, see **Supplementary figures 1-8**). As expected however, we also observed allelic dropouts on the single cell data, with 88.7% of missing SNVs/InDels being heterozygous in bulk data (**Figure 1B** shows an example of mosaic SNV confirmed in bulk at VAF=0.35). On average, over 74% of SNVs/indels in single cell samples were shared with bulk. The remaining single cell only patterns (**Supplementary figures 9-11**) might be true single cell events, but a fraction could also represent amplification errors **(Supplementary tables 4-6**). The single cell-only events comprised 30.2% each C to T and T to C, consistent with previous reports of high quality neuronal somatic SNVs^17^, and distinct from the typical MDA amplification artifacts of C to T^18^, suggesting that our filtering alleviates dMDA artifacts (**Supplementary table 7**). Moreover, on average 34,578 shared between single cells and bulk, 14,999 bulk-only, and 12,431 single-cell specific SNVs/indels overlapped with exons **(Supplementary table 8)**, which was significantly depleted compared to genomic distribution in randomized permutation test (5/5 tests, p<0.05, **Supplementary table 9)**. We found that loci of 7 genes (exons) previously linked to neurodegenerative diseases contained more than 2 SNVs/InDels, found only in single cells **(Supplementary table 10)**. One of the most prominent candidates affected was *LRRK2*, which exhibited 87 small variants (less than 50bp long) found within exons of the gene, with 67 in MSA brain samples (33 found only in single cells), across all the performed sequencing experiments. *LRRK2* is the commonest cause of monogenic Parkinson’s disease^19–22^. *LRRK2* gene variants also modulate the risk of sporadic PD, and it has an apparent role in progression of progressive supranuclear palsy (PSP), another PD-like illness but with different pathology^23^. Overall, we could show that we are able to identify SNVs/InDels in a single cell, some of them being mosaic (**Supplementary figures 1-8**) or absent in bulk (**Supplementary figures 9-11**).

Next, we assessed SV calling on scWGS-LR data. This is complicated as potential chimera from the MDA amplification can lead to false positives^24^. To identify this, we benchmarked our approach with the well established HG002 and the GIAB benchmark. Overall we reached 87.8% F-score for genome-wide SV (GIAB SV v0.6^25^) (see **Supplementary tables 11-13**). Even when assessing the more challenging CMRG benchmark, we achieved an F-score for SV of 89.5% ^26^, thus showing a low rate of chimera after our filtering impacting SV results. Using the same approach on brain samples, we discovered an average of 12,514 insertions and 9,331 deletions in bulk samples (**Supplementary table 2**). For regions where bulk and single cell long-read have >5x coverage we identified an average of 2,414 insertions and 1,965 deletions in bulk samples compared to 2,250 insertions and 2,821 deletions for single cell samples (**Figure 1C**). We found that 78.5% of single cell insertions and 50.9% of deletions can be recalled in corresponding bulk samples (**Figure 1D**) across the experiments, including the previously reported mosaic deletion between a reference Alu and a germline Alu insertion^27^. After removing common population variants, most of the remaining are mosaic in bulk and can be found in about 50% of the studied single cells, with a smaller proportion of variants found in all of them (**Figure 1E**). This is also affected by the fact that, on average, most reads at a given locus come from only two individual cells. The interesting result from this figure is that it highlights the fact that multiple single cell SVs are indeed identifiable via low VAF SV in bulk tissue. Nevertheless, the SVs identified specifically in scWGS-LR once again underscore the diversity of brain tissue. Out of the discovered bulk only variants ∼80% (on average, 73.1% insertions and 87.3% deletions) were heterozygous, consistent with allelic dropout (**Supplementary tables 14-15**). Interestingly, in contrast to small InDels (**Supplementary table 16**) we found an excess of single cell-only deletions; structural variants discovered in single cells-only averaged 525 insertions and 1,433 deletions. Visual inspection confirms the presence of these variants in tested single cell samples and absent in bulk (For examples of deletions, refer to **Supplementary figures 12-35**). For validation purposes, we reviewed short-read paired-end scWGS data after PicoPLEX WGA from the same brains and regions. Despite the limited resolution of short reads, 6.37% of deletions present in single cells and bulk long-read data were also found in short-read PicoPLEX data and confirmed by visual review. Among the long-read single-cell only deletions, although 2.3% of MSA1 and 0.92% of MSA2 were supported in at least one PicoPLEX cell, none were confirmed visually (**Supplementary table 17**). Comparing variant patterns detected in brains to clonally expanded CD8+ T-cells, studied by Hård et al. ^9^, we found that single-cell only deletion/insertion ratio is on average 5.73x higher in tested brains. In contrast, the bulk sequencing of our brains shows the expected imbalance towards the insertions ^28^; genome-wide ratio of DEL/INS in bulk MSA and control brain is 0.75 (**Supplementary table 2**). Moreover, on average 3.47% of single cell deletions confirmed in bulk can be found in exonic sequences, while 4.94% of all bulk deletions overlapped exons (**Supplementary table 18**). The difference was smaller in case of insertions, as 2.50% of single cell insertions confirmed in bulk and 2.40% of all bulk insertions were located within exons (**Supplementary table 19**). For most single cell-only variants, we observed significant depletion in exons compared to genomic exon distribution (permutation test, p<0.05 - 3/5 tests involving single cell-only insertions (**Supplementary tables 20-21**) and 5/5 tests involving single cell-only deletions indicate significant depletion in exons; **Supplementary tables 22-23**). Across the examined variants, we found several deletions overlapping genes, albeit within their intronic sequences, involved in various neurological processes, including *LRRK2* discussed earlier^22^ and *NOTCH2NLB*, involved in prefrontal cortex development^29^. Furthermore, we detect insertions into relevant genes, including *RELN*, that maintains signaling in neurodevelopmental processes in the early postnatal subventricular zone, critical for brain development^30^ (**Supplementary table 24**).

Transposable elements have been previously implicated in neurogenesis and neurodegeneration, as they have a propensity to facilitate Non-Allelic Homologous Recombination (NAHR) events leading to deletions in neurons^31^, and can also create de novo somatic insertions if active. Nevertheless, their occurrence has only been studied in single cell data from the brain with short reads^32,33^. To investigate transposon activity in the neuronal tissue, we annotated members of two most abundant human TEs, LINE/L1 and SINE/Alu within the detected SV. On average, across the 2 MSA brains and 1 control we found 332 bulk insertions and 378 bulk deletions to contain LINE/L1 sequences. Collectively, in the corresponding single cell samples of these brains we captured 261 insertions and 686 deletions that represent LINE/L1 fragments (for detailed breakdown of the reported TEs see **Supplement Section 3**). We only managed to recover truncated fragments of LINE/L1 insertions due to limited read length. Nevertheless, we identified many full length SINE/Alu members (**Supplementary table 25**). Single cell Alu insertions confirmed in bulk were mostly represented by the evolutionarily youngest AluY family, on average found in 382 variants’ sequences (69% of Alu-containing insertions found both in bulk and single cells, on average). Nevertheless, for single cell-only, the majority of Alu insertions (134, 55% of total single cell-only Alu-containing insertions, on average) were AluS (**Figure 2A**). We speculate that this is due to their abundance (∼5:1 ratio of AluS to AluY in reference genome^34^ combined with recombinational activity, which surpasses the propensity of other SINE/Alu members to proliferate in the genome (**Supplementary table 26**).

**Figure 2.**
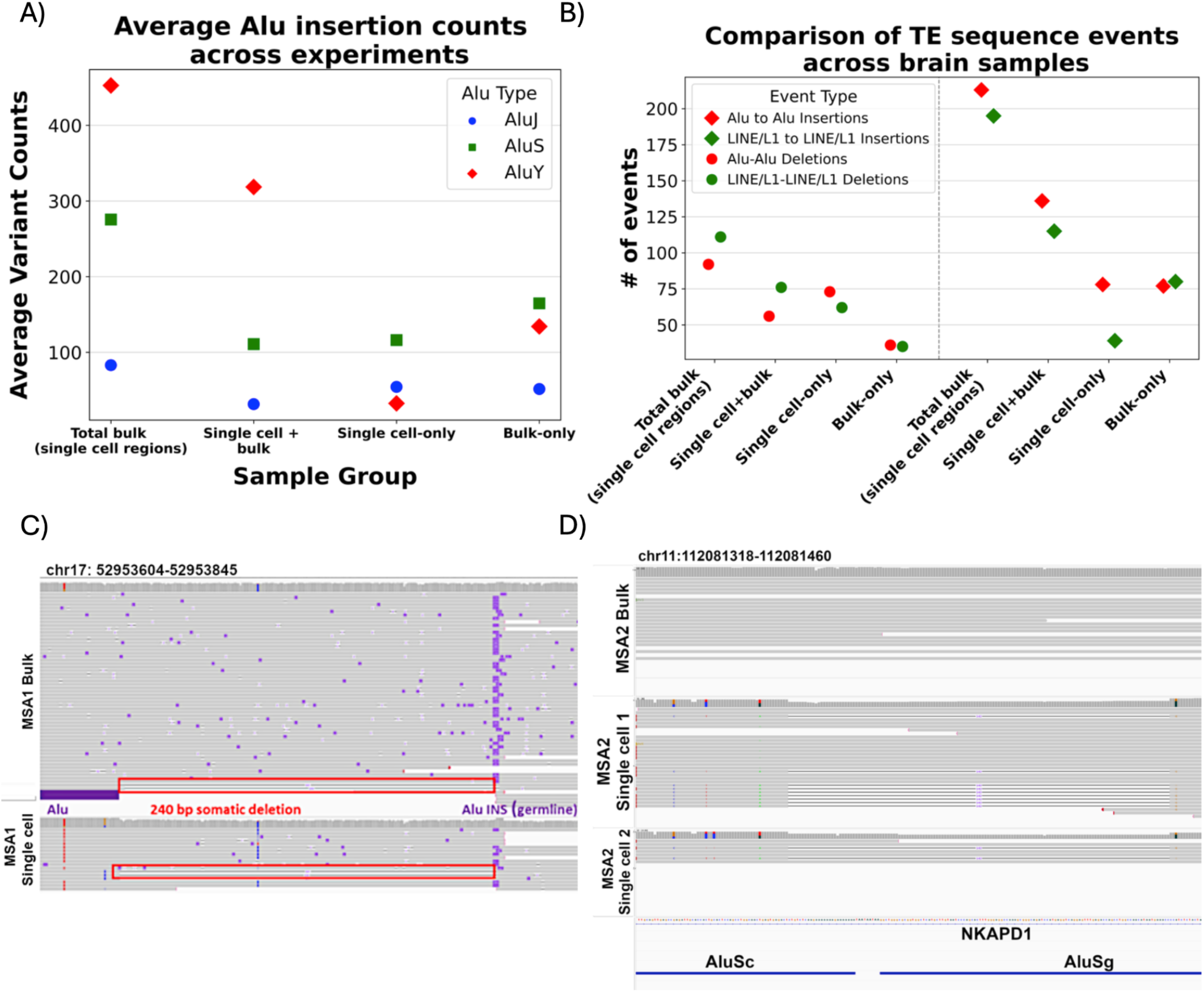
Common transposable element families can influence structural variation by inserting or deleting from the genome, especially via NAHR-mediated deletions. A) Subfamilies of SINE/Alu insertions detected in the single cell samples. The majority of insertions found in bulk or single cells and confirmed in bulk belonged to AluY family, however insertions private to single cells predominantly belonged to AluS subfamily. B) Comparison of detected recombinational deletion events and TEs inserting into loci occupied by the same TE type in the reference genome. We find more SINE/Alu elements to insert within similar loci, presumably due to their abundance and ongoing transpositional activity. However, recombinational activity is higher for LINE/L1 in all samples found in bulk, with slightly less events private to single cells, where SINE/Alu is more active. C) Confirmed germline insertion of Alu element, which overlaps a breakpoint of subsequent somatic deletion; its other breakpoint overlaps reference Alu element, which then causes recombinational deletion. D) Recombinational deletion detected in MSA2 single cell sample and absent in bulk. The deletion present in a subset of tested single cells overlaps with AluSc and AluSg TEs on its breakpoints, suggesting that it occurred due to their recombination. It is within NKAPD1 gene, involved in delayed development syndrome, ataxia and hypotonia.

Further, we were interested in investigating locations in the reference genome where TE-related SVs were found. Initially, we searched for the presence of reference TEs at the locations of detected insertions (**Figure 2B**), as existing germline Alu and LINE1 sequences have been said to attract novel somatic TEs within the same family^34,35^. Analysis of TE insertion locations in our brain samples showed 28.7% of single cell Alu insertions confirmed in bulk and 51.1% single cell-specific SINE/Alu insertions occurred within Alu reference elements (for LINE/L1, 67.9% single cell confirmed in bulk and 30.5% of single cell-specific and occurred within reference INE/L1, respectively). This represents an average enrichment of 3.26-fold of Alu and LINE1 insertion into a similar reference sequence, rather than randomly in the genome (assuming 17% of the reference genome is LINE1 and 11% is SINE/Alu), consistent with the data showing that reference elements attract novel SINE/Alu and LINE/L1 insertions (**Supplementary table 14**). Conversely, of the insertions within SINE/Alu and LINE/L1 in reference, 46.9% of single cell-specific events contained SINE/Alu sequence (79.6% of single cell confirmed in bulk), while for single cell-specific LINE/L1 it was 46.5% (39.5% for LINE/L1 confirmed in bulk).

To gain insight into the nature of observed deletions we examined their sequences, specifically searching for pairs of transposable elements belonging to the same families/types and overlapping deletions’ breakpoints, thus suggestive of being able to facilitate recombinational events. Our previous findings already hinted at NAHR mediated repeat recombinants events^36^ between two SINE/Alu elements (**Figure 2C**). As before we could identify these events as mosaic in bulk but also identifiable in single cell data sets. Across three brains, we detected an average of 56 deletions with SINE/Alu pairs and 76 deletions with LINE/L1 pairs at their breakpoints. These deletions are present in single-cell sequencing and mosaic in the corresponding bulk samples. Thus, maybe not surprisingly, on average across 3 tested brains we also identified NAHR mediated repeat recombinants (**Figure 2D**) present in single cells only with 73 for SINE/Alu (17.6% of single cell-specific Alu deletions; see **Supplementary figures 36-38**) and 62 for LINE/L1 (12.9% of single cell-specific LINE/L1 deletions; see **Supplementary figures 39-41**). Some of these events overlapped disease-relevant genes, including Alu-Alu mediated deletion overlapping *EVA1C* gene^37^, which deleted exon 1 and thus might have an impact on the gene itself. Similarly, a LINE-LINE deletion was detected in the intronic sequence of *DOCK3* gene; mutations of which have been linked to ADHD^38^. To validate the reported associations between transposable elements and detected variants within single cells we performed a permutation test over deletions; we found that in 17 out of 20 single-cell comparisons, these were significantly transposon-associated (see methods, **Supplementary tables 27-30**). This overall highlights the dynamic nature of transposons and their importance to be studied from bulk down to single cell data sets.

To summarize, we demonstrated the utility of long-read single cell whole genome sequencing using Oxford nanopore to study SNV, InDel and SV mutations across 18 single cells derived from 3 brains. To enable this, multiple innovations were necessary, including an improved filtering pipeline to mediate the effects of MDA amplification such as chimera formation and single base errors. These novel approaches lead to potentially the best characterisation of mutational landscape in single cells from the brain thus far revealing novel insights into SV and transposon formation and impact. We highlighted multiple new insights across repeat recombinants in the brain which are identifiable as low VAF in bulk but easier to identify in single cell sequencing. These mutations are regularly overlooked in standard short read single cell sequencing, given its focus on large CNVs. This is despite the fact that these mutations seem to impact exons that are getting deleted in a fraction of cells, however the variants are generally depleted in exonic locations. While this study lacks the power to conclude something for MSA, it clearly highlights the lack of awareness of these mosaic or single cell SV. Overall, we found that there is indeed an increased level of deletions in brain single cell data compared to T-cell single cell data (5.73-fold increase) that was previously published^9^. Further studies need to investigate if this is due to the limited regeneration capacity of neurons. Nevertheless, these findings can be significant given previous reports for the potential mosaic or single cell mutational impact on neurological diseases^12,15,16,18^.

## Methods

### Sequencing

#### Single cell sequencing

We used previously generated dMDA amplicons from single nuclei from two MSA samples (cingulate cortex) and one control (frontal cortex), n=6 cells from each, which had already undergone short read low coverage WGS^13^. These were derived from fresh frozen *post-mortem* brain samples provided by the Queen Square Brain Bank, London, UK. All donors had given informed consent for the use of their brain in research and ethics approval was provided tissue bank by the UK National Research Ethics Service (07/MRE09/72). All ethical regulations relevant to human research participants were followed. dMDA products were purified using Ampure XP beads, using a 0.8x bead to sample ratio, and underwent library preparation using two different methods. RBP (rapid barcoding PCR;^39^ or T7 debranching^40^. For RBP, 5 ng of purified DNA was used with the ONT rapid barcoding PCR kit (Oxford Nanopore Technologies, SQK-RBP004) following the manufacturer’s instructions. Libraries were pooled to a final molarity around 100 fmol in 10 μl, assuming an average library size around 2 kb. Pools were sequenced on a MinION MK1b (Oxford Nanopore Technologies, MIN-101B) using an R9.4.1 flow cell (Oxford Nanopore Technologies, FLO-MIN106D). For T7, 500 ng DNA input was used with 1 unit T7 endonuclease (NEB) in 30 μl and incubated at 37° C for 1 hour, followed by bead purification. All recovered DNA was used for library preparation according to the “sequencing gDNA – Native Barcoding Kit 24 V14; Version: NBE_9169_v114_revQ_15Sep2022” protocol, followed by native barcode (SQK- NBD114.96) and sequencing adapter ligation. The MSA1 pool was sequenced on a MinION MK1b R9.4.1 flow cell (FLO-MIN106D), but, due to the low yield, the MSA2 and control pools underwent PromethION sequencing using a R10.4.1M flow cell for each (FLO-PRO114M) at the Nottingham University DeepSeq core, with a nuclease flush and reload to maximize yield. Note that, due to the high DNA input required, the MSA1 T7 sequencing could not be repeated. For bulk DNA analysis, we extracted DNA using the Qiagen MagAttract kit as per manufacturer protocol, using 20 mg starting material. MSA1 long-read WGS data was available previously ^27^, and MSA2 and control ONT long-read samples were processed at the Baylor College of Medicine (BCM) core using the same protocol as MSA1. Sample MSA2 had an average size of 8.7 kb and did not require shearing. The control sample had an average size of 52.6 kb and was sheared using g-tubes (Covaris). Three µg of DNA was added to the g-tube and centrifuged at 3,800 rpm (4- passes) to achieve an average sheared size of 15-20 kb.

#### Bulk sequencing

Library preparation and sequencing for bulk ONT sequencing of MSA2 and control brain cortex was performed at the BCM core; data from MSA1 was already available^36^. Genomic DNA was quantified using the Qubit dsDNA Quantification Broad Range Assay (ThermoFisher Scientific) according to the manufacturer’s instructions. DNA size was determined using the Agilent Femtopulse. Sample MSA2 had an average size of 8.7 kb and did not require shearing. The control sample had an average size of 52.6 kb and was sheared using g-tubes (Covaris Part number 520079). Three µg of DNA was added to the g-tube and centrifuged at 3,800 rpm (4- passes) to achieve an average sheared size of 15-20 kb. Following bead purification using AMPure XP beads (Beckman Coulter), the sheared DNA was size-selected on the PippinHT instrument (Sage Science) using the 6-10 kb High-Pass definition. The minimum size selection threshold was set at 10 kb. Three µg of MSA2 was size selected on the Blue Pippin instrument (Sage Science) using the 4-10 kb High-Pass definition with a minimum size threshold of 4 kb. The loading and elution of samples from the PippinHT/ Blue Pippin cassettes were performed following the manufacturer’s instructions. The eluted samples were bead purified using AMPure XP beads (Beckman Coulter) and resuspended in 50 µL nuclease free water. 1 µL of this eluate was run on the Agilent 2100 Bioanalyzer using the DNA 12000 chip to determine the average size of the sample. 48 µL of this eluate was used as input for the ONT SQK-LSK114 (Lot# LSK114.20.0001) library preparation kit. End repair/damage repair and adapter ligation were performed as per manufacturer’s instructions. Final libraries were quantified using the Qubit dsDNA Quantification Broad Range Assay (Thermo Fisher Scientific).

### Processing of HG002 sample

Data were aligned to GRCh37/38 reference genomes using minimap2 v2.22 (RRID:SCR_018550; https://github.com/lh3/minimap2)^41^ and converted to binary format using Samtools v1.19.2 (RRID:SCR_002105; https://github.com/samtools/)^42^. Mosdepth v0.3.3 (RRID:SCR_018929; https://github.com/brentp/mosdepth)^43^ was used to determine the per-base coverage for each sample. Clair3 v1.0.0 (RRID unavailable; https://github.com/HKU-BAL/Clair3)^44^ was used to obtain SNV calls for each sample. Calls were split into substitutions and small In/Dels, which were further filtered for those encompassing 5 or more reads of coverage using BEDTools v2.30.0 (RRID:SCR_006646; https://github.com/arq5x/bedtools2)^45^, with at least 3 reads supporting each call and “PASS” in the VCF filter field. Results were compared to HG002 GiaB reference SNV calls using RTGtools v3.12.1 (https://github.com/RealTimeGenomics/rtg-tools)^46^. The SV calls were compared to HG002 ONT sequencing calls obtained from Oxford Nanopore Technologies, by running Sniffles v.2.2 (RRID:SCR_017619; https://github.com/fritzsedlazeck/Sniffles)^27^ with available reference genomes (GRCh37 and GRCh38 respectively). To remove calls presumably resulting from chimeric reads, we ignored all duplications and inversions, and applied similar chimera filtering as to single-cell samples (**see Supplement section 2**). Resulting SV calls were filtered using PASS filter in the *vcf* file; sequences were selected for those longer than 50bp, supported by 3 or more reads in locations covered by at least 5 reads. The detected structural variants were compared to a HG002 GiaB SV dataset using Truvari benchmark v4.0.0 (RRID unavailable, https://github.com/ACEnglish/truvari)^47^. Custom scripts were used to annotate and summarize results.

### Benchmarking with H002 low-input sample

The single-cell and corresponding bulk samples were analyzed similarly to HG002 sample. Reads were aligned to GRCh38/Hg38 reference genome using minimap2 and converted to binary format using Samtools (software versions same as above). Initial sequencing metrics were assessed with Cramino v0.13.1 (RRID unavailable, https://github.com/wdecoster/cramino^48^; for read length distribution, see **Supplementary figures 42-47**). Binary files were merged within the same experiment and tissue type with samtools. To reduce chimera signal, initial reads were filtered using methodology described in **Supplement section 2**. Mosdepth v0.3.3 (RRID:SCR_018929; https://github.com/brentp/mosdepth)^43^ was used to determine the per-base coverage for each sample. Clair3 v1.0.0 (RRID unavailable; https://github.com/HKU-BAL/Clair3)^44^ was used to obtain SNV calls for each of the samples. SNV calls were merged with GLNexus, version 1.4.1 (RRID unavailable, https://github.com/dnanexus-rnd/GLnexus)^49^. Calls were split into substitutions and small In/Dels, which were further filtered for those encompassing 5 or more reads of coverage, with at least 3 reads supporting each call and “PASS” in the VCF filter field. Single-cell results were compared with their bulk counterparts using custom scripts. Results were traced to their original cells and the proportion of single-cell and multi-cell variants absent in bulk tissue was calculated.

### Detection of variants across analyzed brain samples

Binary sequencing files were analysed for SVs using Sniffles v2.2 ^27^. Initial SV calls were summarized with Python scripts to characterize signal from all types of SVs (insertions, deletions, duplications, inversions and breakends/translocations). To avoid chimeras, we filtered inversions and duplications^50^ (summary for these can be found in **Supplementary table 31**), as well as applied other filters described in **Supplement section 2** (For sizes of initially called inversions and duplications, refer to **Supplementary figures 48-53**).

All SV filtering steps were collated into a single script that obtained support and coverage information from the vcf tags. The individual single-cell samples from RBP MinION and T7 PromethION experiments were summarized with that script and ratios of INS/DEL were compared between single-cell samples. Variants detected in CD8+ T-cell ^9^ were analysed with a custom set of scripts, reflecting the steps applied to MSA brain samples. SVs were split into insertions and deletions, and for each merged experiment they were compared with results from corresponding bulk samples. Based on the resulting numbers, we determined the variants private to single cells, variants shared between single cell samples and corresponding bulk as well as bulk-only variants (limited to regions of >=5x coverage recovered from corresponding single cells). We calculated the proportions of bulk-only mosaic variants and variants with 0/1 bulk-only genotype with custom scripts (for variant size distribution refer to **Supplementary figures 54-65, for coverage see Supplementary figures 66-67**. To cross-compare the detected variants with short-read Illumina scWGS samples, we used Illumina Manta v1.6.0 (RRID: SCR_022997, https://github.com/Illumina/manta)^51^ on data from each of the 3 brains (MSA1: 15 cells, MSA2: 12, control: 7). Using SVtyper v0.7.1 (RRID unavailable, https://github.com/hall-lab/svtyper)^52^, we determined how many LR-scWGS reported variants overlapped with SR-scWGS samples.

### Annotation of brain variants

Sequences of detected insertions and deletions were extracted from the VCF file and corresponding reference genome with custom scripts into FASTA format. RepeatMasker v4.1.5 (RRID:SCR_012954, https://www.repeatmasker.org/)^53^ was used to annotate the recovered variant sequences for presence of retrotransposons. TE annotation of brain samples was analyzed using custom Python scripts, to define the variants containing most abundant and active transposon families, SINE/Alu and LINE/L1. The purpose of these was to parse RepeatMasker output and select results containing target TEs, track variant counts, and provide statistics on variants/elements as an output. The scripts are available via github repository, linked below. To collapse results into TE elements and determine the completeness of detected transposons, a “One code to find them all’’ v1.0 tool was used (RRID unavailable, https://doua.prabi.fr/software/one-code-to-find-them-all)^54^, followed by custom scripts to summarize results. Furthermore, we analyzed the proportion of TE insertions into preexisting repeated elements of the same family. SINE/Alu elements’ subfamily was determined from the annotation, as well as the actual number of fragmented elements/elements taking more than 80% of the entire insertion. Next, we detected the proportion of deletions containing SINE/Alu or LINE elements and deletions in which the same type of transposable element can be found on the opposite breakpoints of the variant. The distribution of deletions containing SINE/LINE TEs was used as template for permutation test; BEDTools shuffle algorithm was utilized to simulate single-cell deletion distribution across the reference Hg38 genome, which was compared to brain samples results’ with Z-test. RepeatMasker results were filtered for Alu subfamilies, using custom scripts.

To determine the propensity of variants for mutating known genes, we used AnnotSV version 3.4.2 (RRID unavailable, https://www.lbgi.fr/AnnotSV/)^55^ to find the overlap between INS/DEL and gene locations, particularly with a list of genes involved with neurodegeneration compiled from literature. Furthermore, we used bcftools intersect version 1.19 (RRID:SCR_002105; https://github.com/samtools/bcftools)^42^ to find the overlap between variants (SVs and SNVs/InDels) and known exon locations extracted from Gencode v45 (RRID:SCR_014966; https://ftp.ebi.ac.uk/pub/databases/gencode/Gencode_human/)^56^. Similarly to TEs, we performed permutation tests using locations of deletions and insertions, as well as single-cell SNVs/InDels, in order to determine the correlation with reference exons with Z-test. We determined a number of genes related with MSA in the literature, and used scripts to filter the AnnotSV/Gencode results for genes affected by SVs and related to MSA. To determine population frequency of the variants detected in single cells and bulk simultaneously, we used stix-suite 1.0.1 (RRID unavailable, https://github.com/ryanlayer/stix?tab=readme-ov-file#stix-suite)^57^. Visual inspection of detected variants as well as overlap with genomic annotations was performed using IGV version 2.18.2 (RRID: SCR_011793, http://www.broadinstitute.org/igv/)^58^

## Data availability

All single cell ONT data, and bulk ONT data for MSA2 and control, will be available at the European Genome-Phenome Archive (EGA) (xx_). MSA1 bulk ONT data were previously deposited in the Sequence Read Archive (SRA) bioproject ID PRJNA985263.

GIAB HG002 benchmark data links: https://www.nist.gov/programs-projects/genome-bottle https://ftp-trace.ncbi.nlm.nih.gov/ReferenceSamples/giab/release/AshkenazimTrio/HG002_NA24385_son/latest/ https://ftp-trace.ncbi.nlm.nih.gov/ReferenceSamples/giab/release/AshkenazimTrio/HG002_NA24385_son/CMRG_v1.00/

## Code availability

For detailed methodology, including instructions for every step of the analysis and all custom code, please refer to the github repository (https://github.com/michalizydo/scONT).

## Contributions

M.B.I., F.J.S. and C.P. designed the study. E.K-E., D.W.H., M.T. and N.H. performed single cell sequencing, supervised by C.P. Y.H, H.H.M. and D.M.M performed bulk sequencing. M.B.I performed *in-silico* analysis, supervised by F.J.S and C.P, with additional data processing and analysis by X.Z., A.A., D.W.H and Z.G.S. M.B.I, F.J.S and C.P wrote the manuscript. All authors contributed to reviewing the manuscript.

## Acknowledgements

We would like to thank Richard Gibbs for constructive discussions and input for this study. The study is funded in part by the joint efforts of The Michael J. Fox Foundation for Parkinson’s Research (MJFF) and the Aligning Science Across Parkinson’s (ASAP) initiative. MJFF administers the grant [Grant ID 000430] on behalf of ASAP and itself. This study is in part supported by NIH (UG3NS132105).

