## Supplementary text for "Single cell long read whole genome sequencing reveals somatic transposon activity in human brain"

### Supplement

**Supplement Section 1: Application of T7 and PRB protocols to remove secondary structures**

In this study, we utilized two different library preparations for all cells: T7 debranching, the standard method to remove displaced strands created by MDA; and the PCR rapid barcoding protocol (RBP), which should eliminate all side chains by creating linear molecules, at the possible risk of smaller read lengths impacting SV detection.

As T7 debranching performed on a MinION device for MSA1 yielded only 2.64Gb, T7 debranching for MSA2 and control benefited from the high sequencing throughput of the PromethION device (**Supplementary table 1).** Additionally, we used RBP protocol, which led to high yield from MinION - over 7.4*10^10 bases on (~62.5M reads) and 10.6*10^10 bases on PromethION T7 (~81.5M reads) across the 6 cells of each brain. The average read N50 was 2.79kb for both MinION RBP and Promethion T7 library samples, with minor differences across individual experiments, although the T7 provided a wider range of read sizes, with 4.5M reads > 3kb (MSA2; ~250k reads for MSA1 T7, 5.9M for control), and some as long as 300kb (**Supplementary figures 42-47**). Due to low yield of the MSA1 T7 MinION, and the lack of additional material for T7 resequencing, it was excluded from further comparisons. Merging single-cell data from the same brain region produced more uniform coverage in each sample, while retaining the ability to trace individual variants to cells they originate from. In MSA2 and control, where we have T7 PromethION and RBP MinION, the PromethION as expected provided increased coverage to >=5x depth (in MSA2 MinION RBP - 36.4% and PromethION T7 - 45.7%, and in control 15.0% and 21.7%, respectively. For MSA1 MinION RBP produced 35.4% of the genome). In contrast, Illumina MDA sequencing run of MSA1 covers 61.1% of the reference genome, MSA2 covers 62.5% while control brain covers 47.4% at >=5x (total Illumina >=1x coverage: 81.8% for MSA1, 86.3% for MSA2, 72.6% for control brain). Overall, the PromethION device combined with T7 debranching delivers largest yield of reads with high, sustained N50 of over 2.8kb, covering up-to 46% of the Human genome with high-quality assembly from single-cell data and we recommend it as it captures the most signal within the tested pool of single cells.

**Supplement Section 2: Filter for chimeric SV caused by MDA amplification**

During our initial analysis of the Sniffles2 SV calls, we observed that some of them were overlapping regions of the reads that did not align to the reference genome correctly, showing a large proportion of misaligned bases and particularly, numerous deletions over a short span of the read, often close to its end (see **Supplementary figure 68**). Furthermore, some of the reads had multiple splits, with the fragments clearly misaligned. Therefore, after testing multiple approaches, we determined to filter sequencing data, removing reads that had more than 1 split based on SAM “SA:” tag, as well as those, which more than 5% of their total bases differed from the reference genome, based on “MD:” tag. This filtering was applied to initial alignments of reads for each of the analyzed single-cell experiments, and the remaining reads were further re-evaluated for variants according to steps described in methods sections.

**Supplement Section 3: Detailed breakdown of transposable elements detected in brain samples.**

On average, across the 2 MSA brains and 1 in control, we found 332 bulk insertions and 378 bulk deletions to contain LINE/L1 sequences (out of 2414 insertions and 1965 deletions, respectively). Collectively, in the corresponding single cell samples of these brains we captured 261 insertions and 686 deletions (out of 2249 and 2821, respectively) that similarly showed LINE/L1 fragments. Out of these variants, 94 insertions and 479 deletions were single-cell only, 168 insertions and 208 deletions were shared between bulk and single-cells and 165 insertions/170 deletions were bulk-only. Next, we performed a similar search for SINE/Alu elements. On average 689 insertions and 534 deletions detected in bulk contained SINE/Alu sequences, whereas for single-cell samples, 646 insertions and 775 deletions contained SINE/Alu. This breaks down into 164 insertions and 418 deletions containing SINE/Alu found only in single cells, with the remaining 482 insertions and 357 deletions shared between single cells and bulk and 207 insertions/177 deletions being bulk-specific.
