## Supplementary figures for "Single cell long read whole genome sequencing reveals somatic transposon activity in human brain"

1. Example of mosaic SNV detected in MSA2 bulk brain and corresponding single cells.

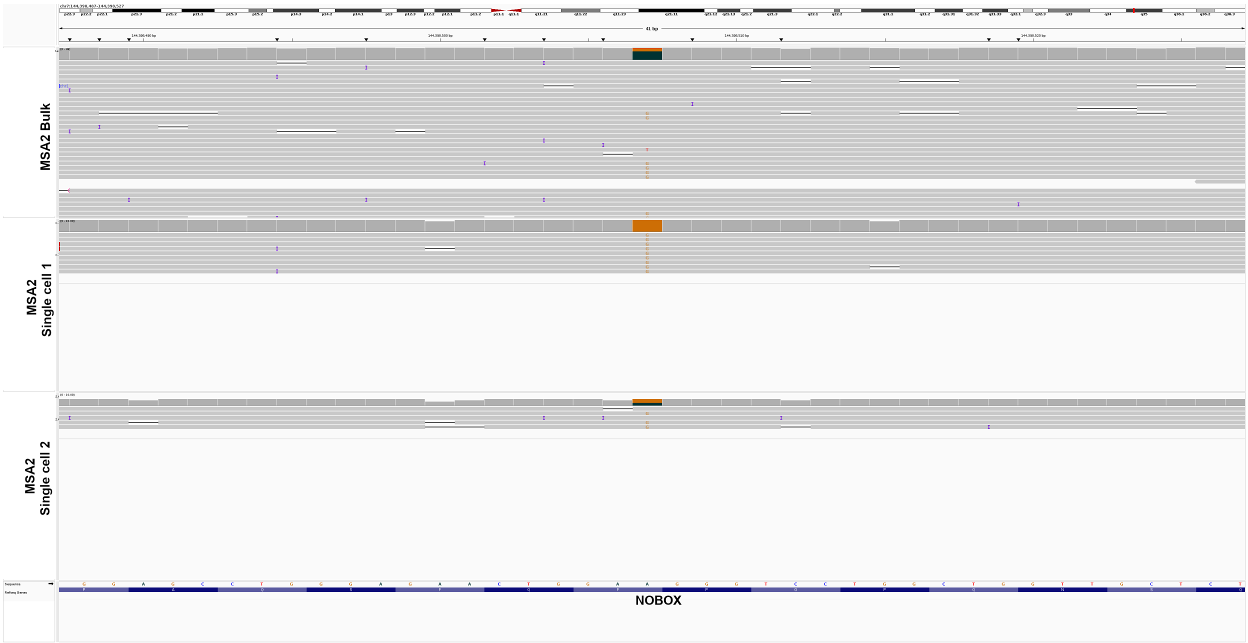


2. Example of mosaic SNV detected in MSA2 bulk brain and corresponding single cells.

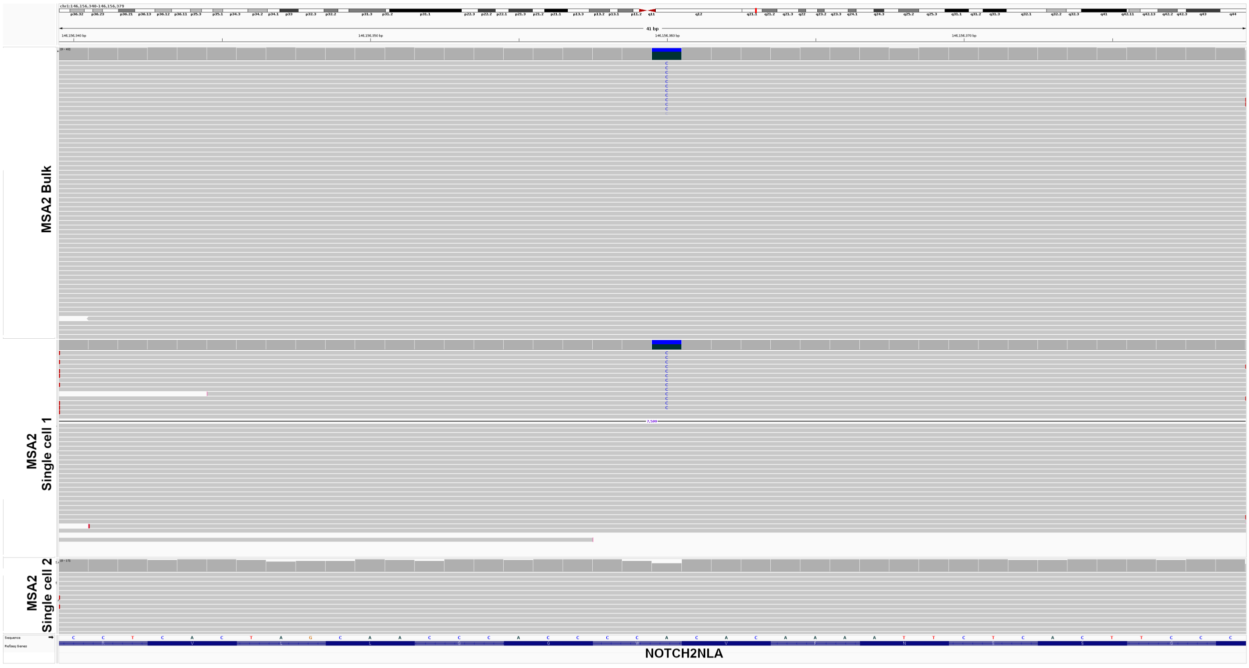

3. Example of mosaic SNV detected in MSA2 bulk brain and corresponding single cells.

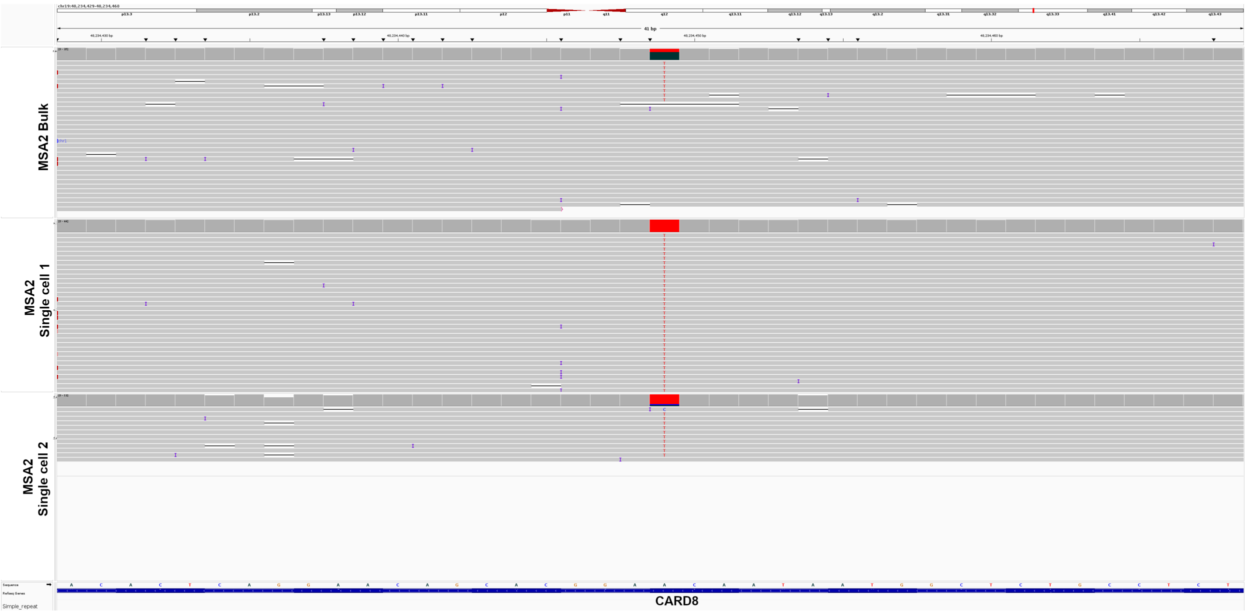

4. Example of mosaic SNV detected in MSA1 bulk brain and corresponding single cells.

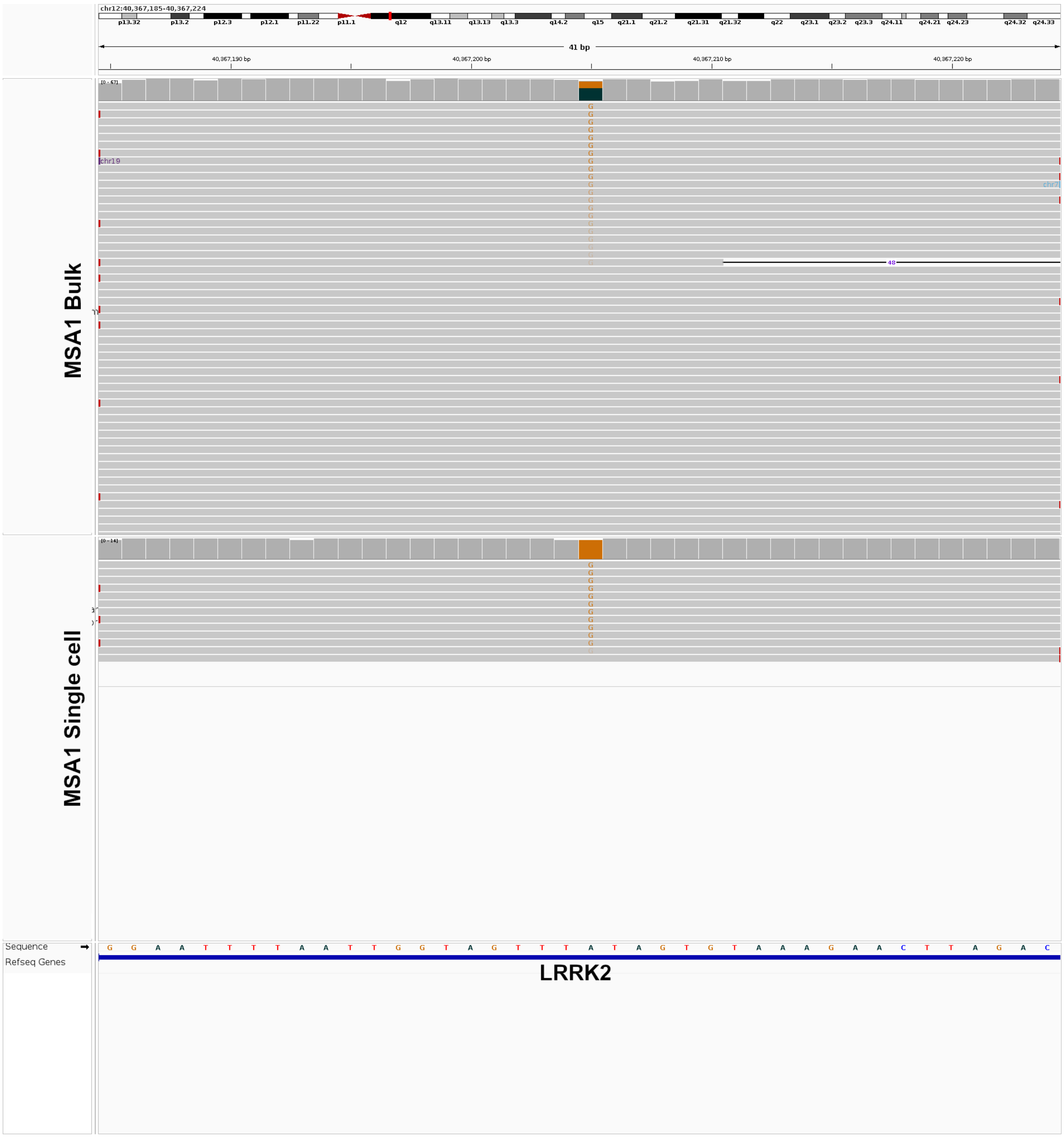

5. Example of mosaic SNV detected in MSA1 bulk brain and corresponding single cells.

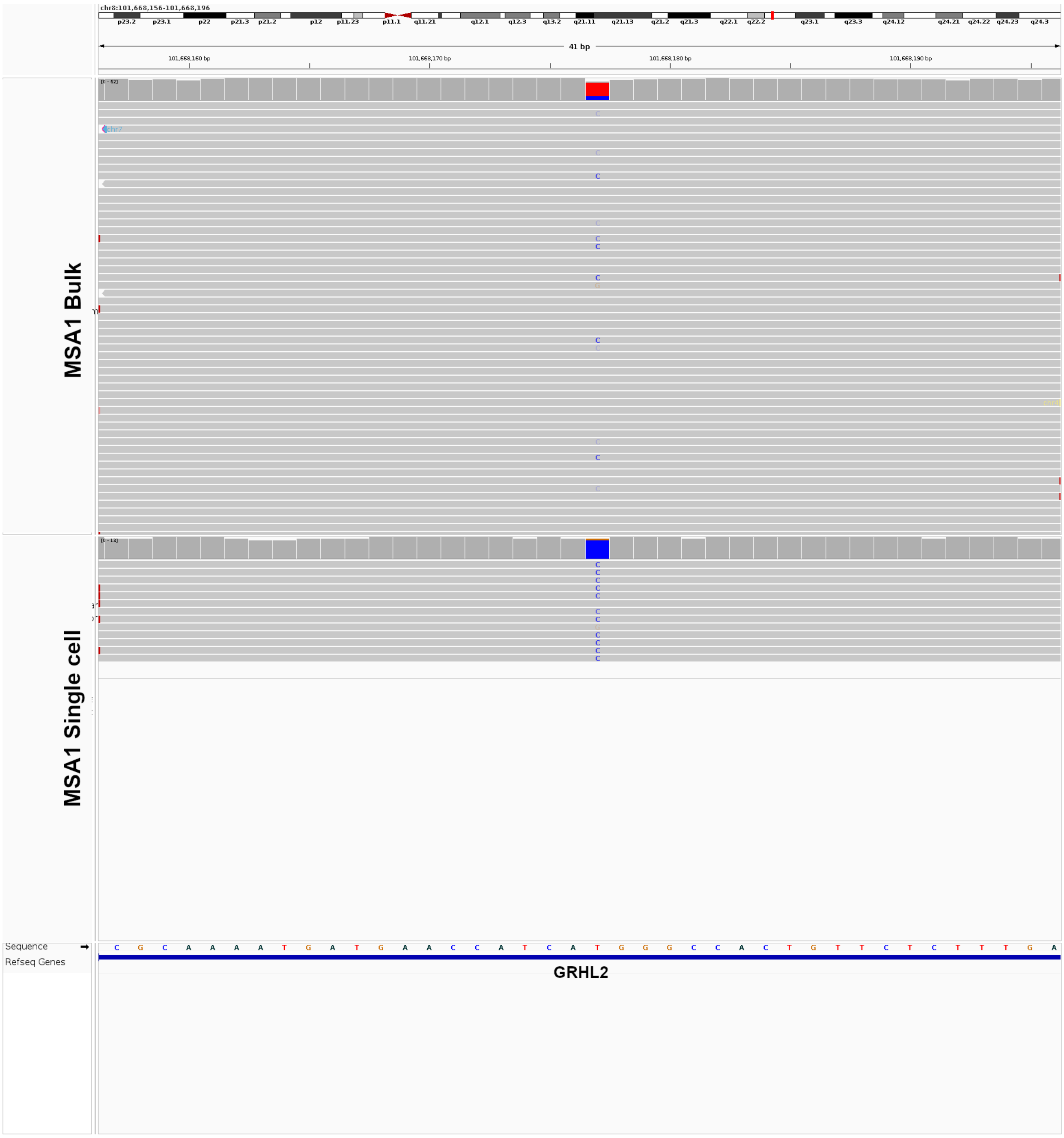

6. Example of a mosaic SNV detected in MSA1 bulk brain and corresponding single cells.

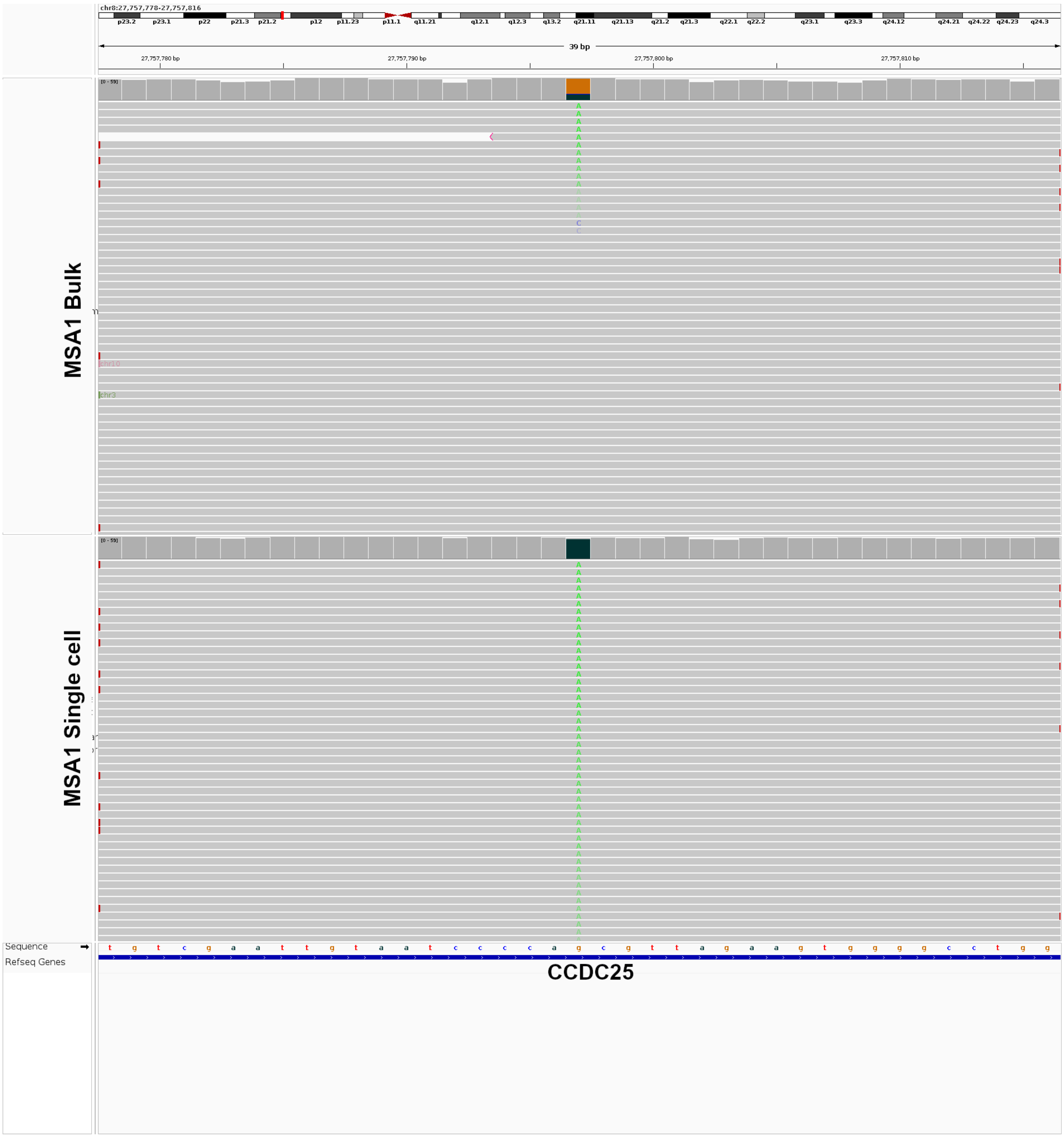

7. Example of a mosaic SNV detected in MSA1 bulk brain and corresponding single cells.

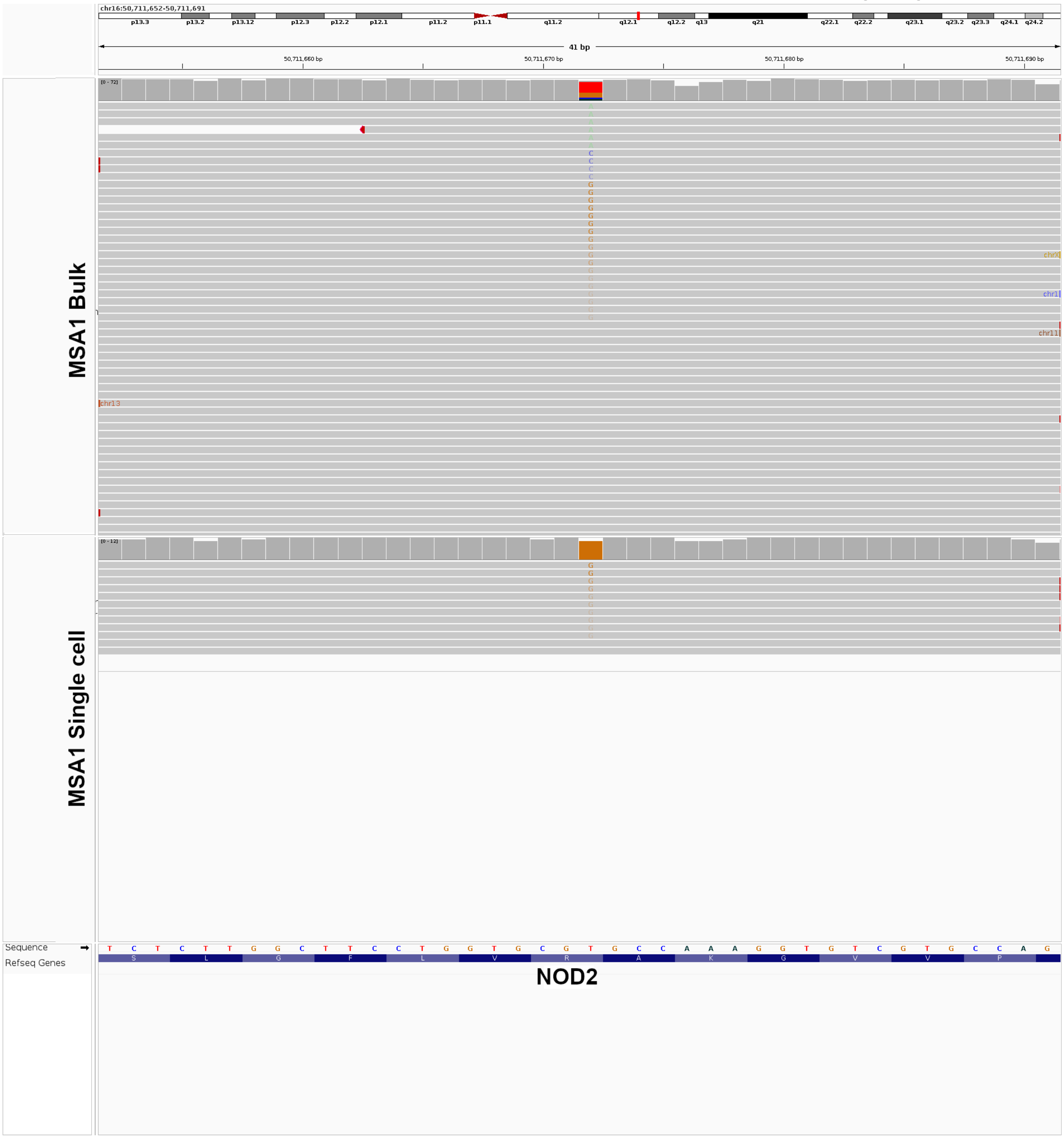

8. Example of a mosaic SNV detected in MSA1 bulk brain and corresponding single cells.

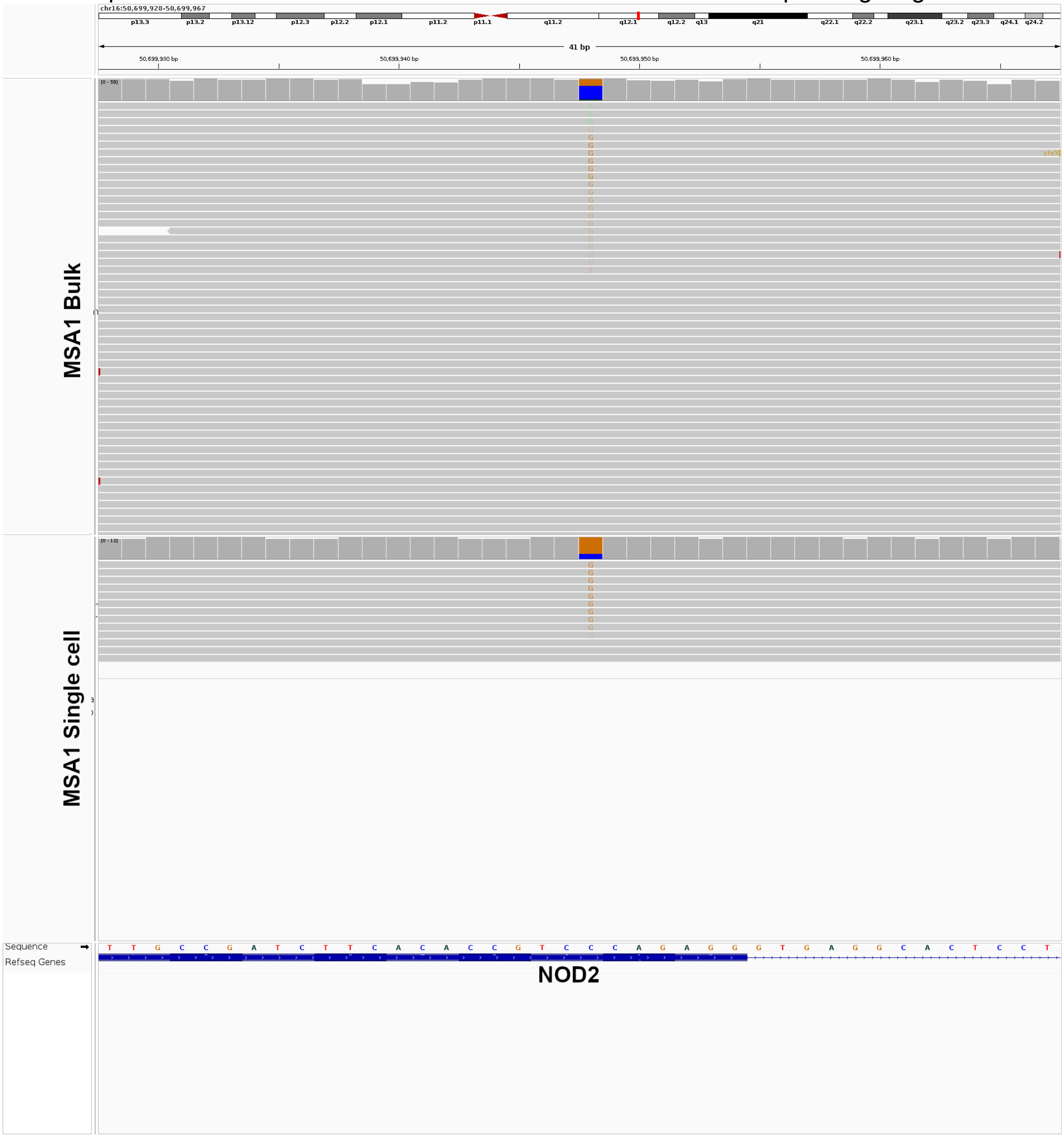

9. Example of single-cell only SNV detected in MSA2 brain.

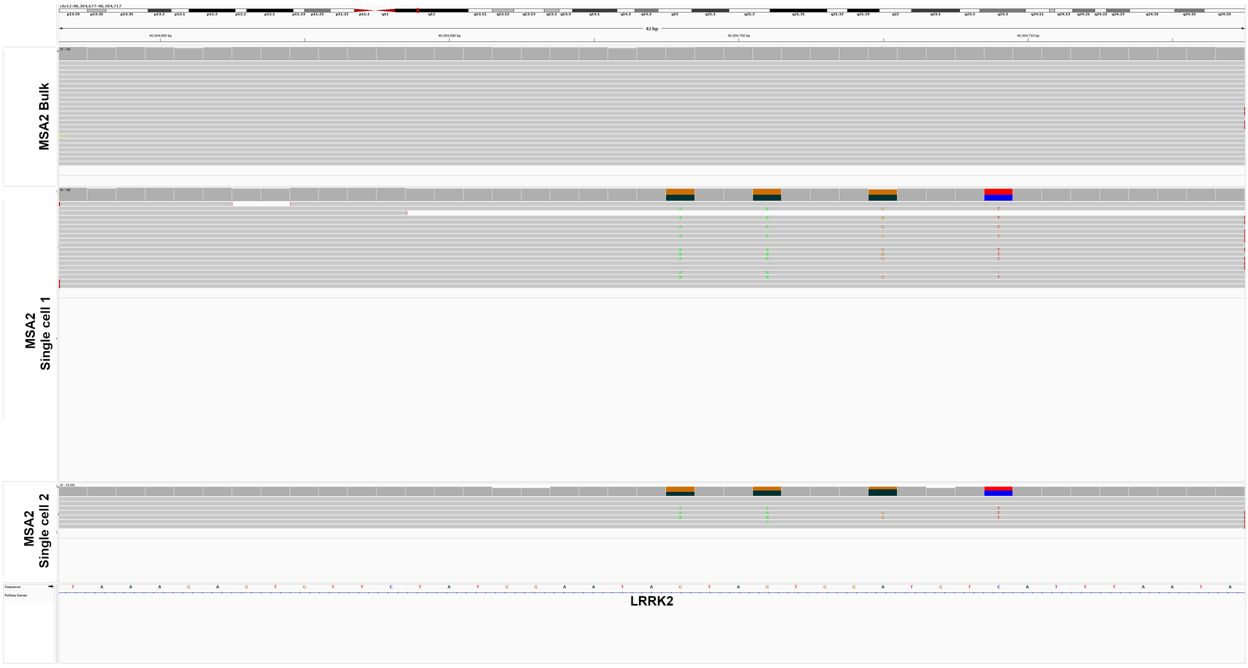


10. Example of single-cell only SNV detected in MSA2 brain.

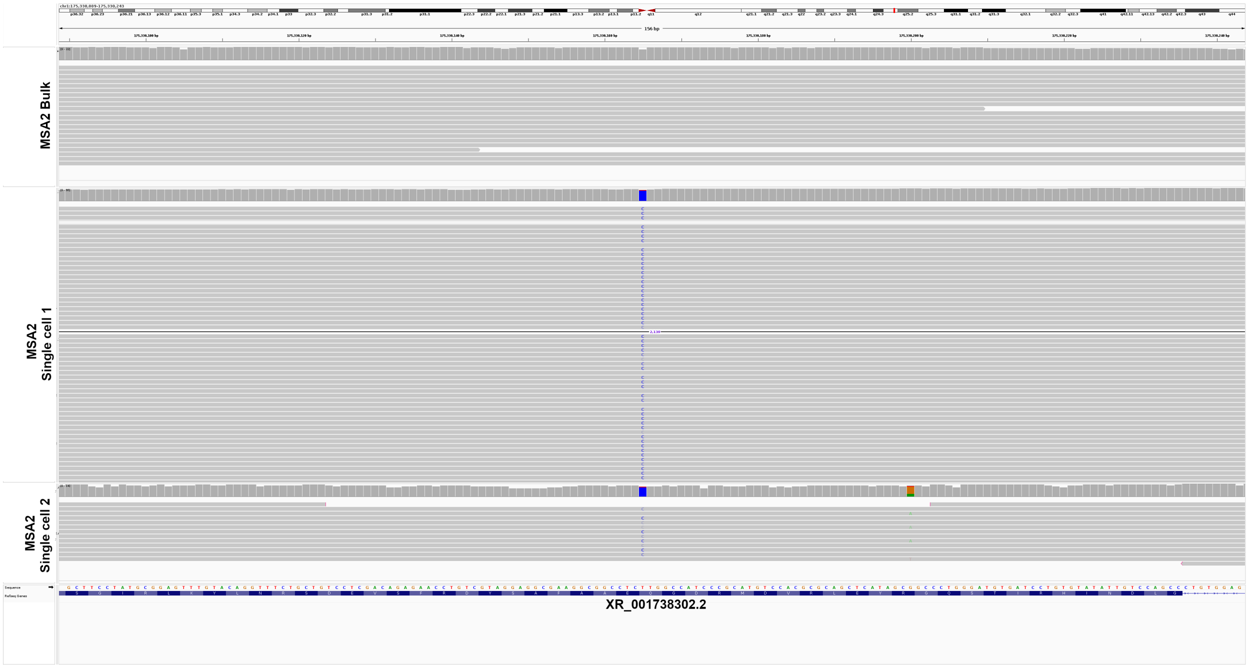

11. Example of single-cell only SNV detected in MSA2 brain.

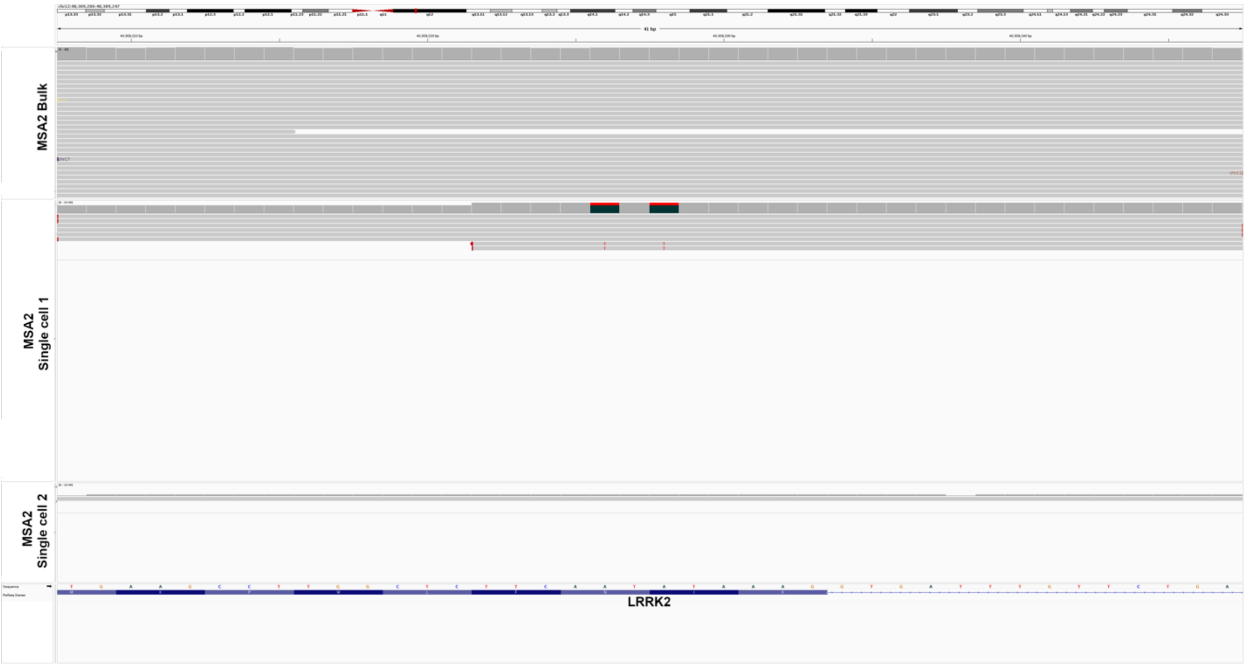


12. Example of a single cell-specific deletion detected in MSA1 brain.

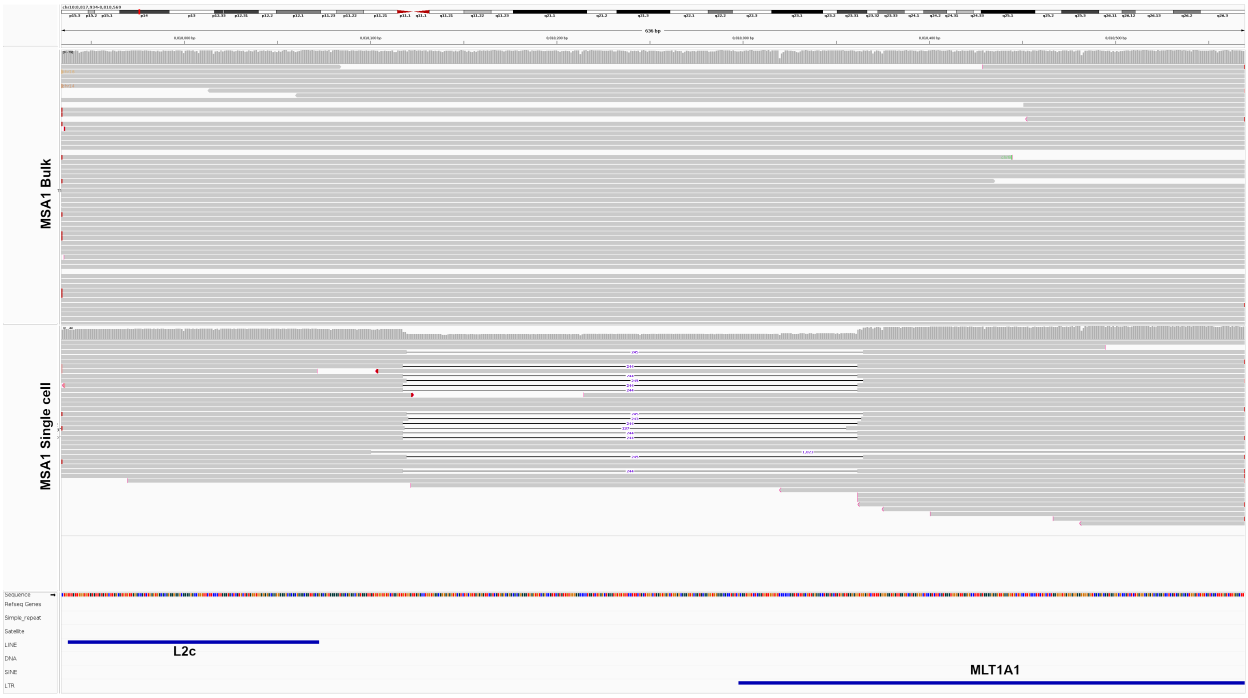

#### 13. Example of a single cell-specific deletion detected in MSA1 brain.

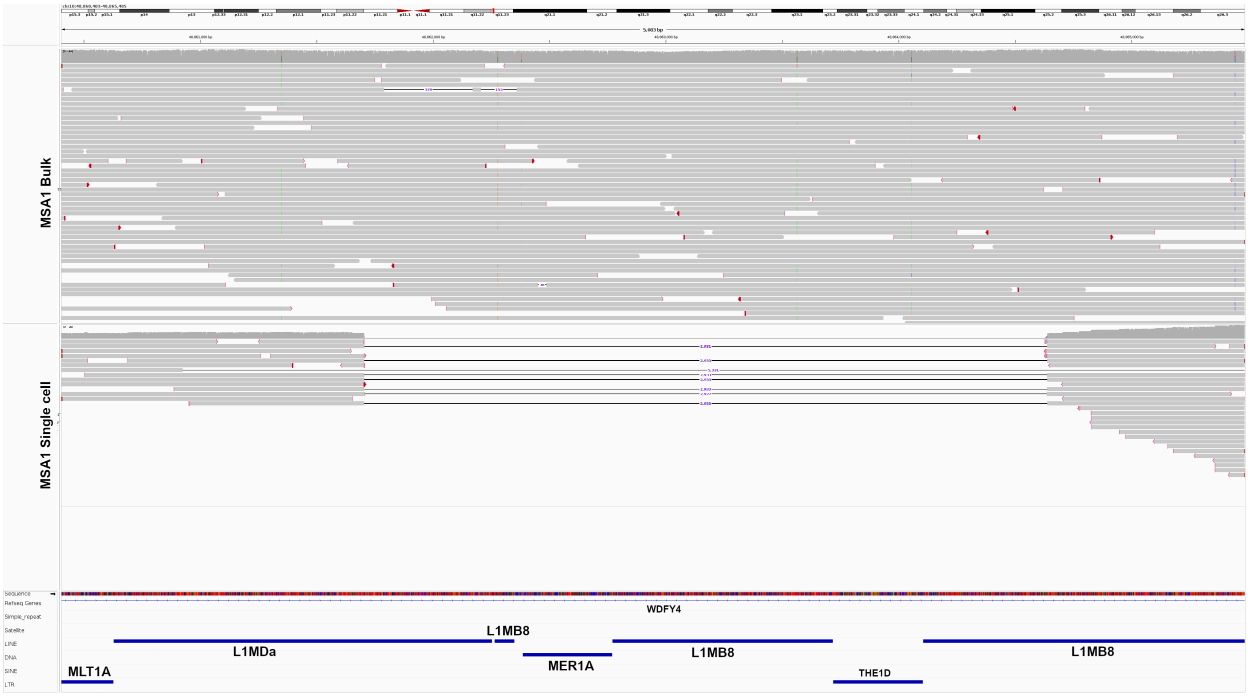


#### 14. Example of a single cell-specific deletion detected in MSA1 brain.

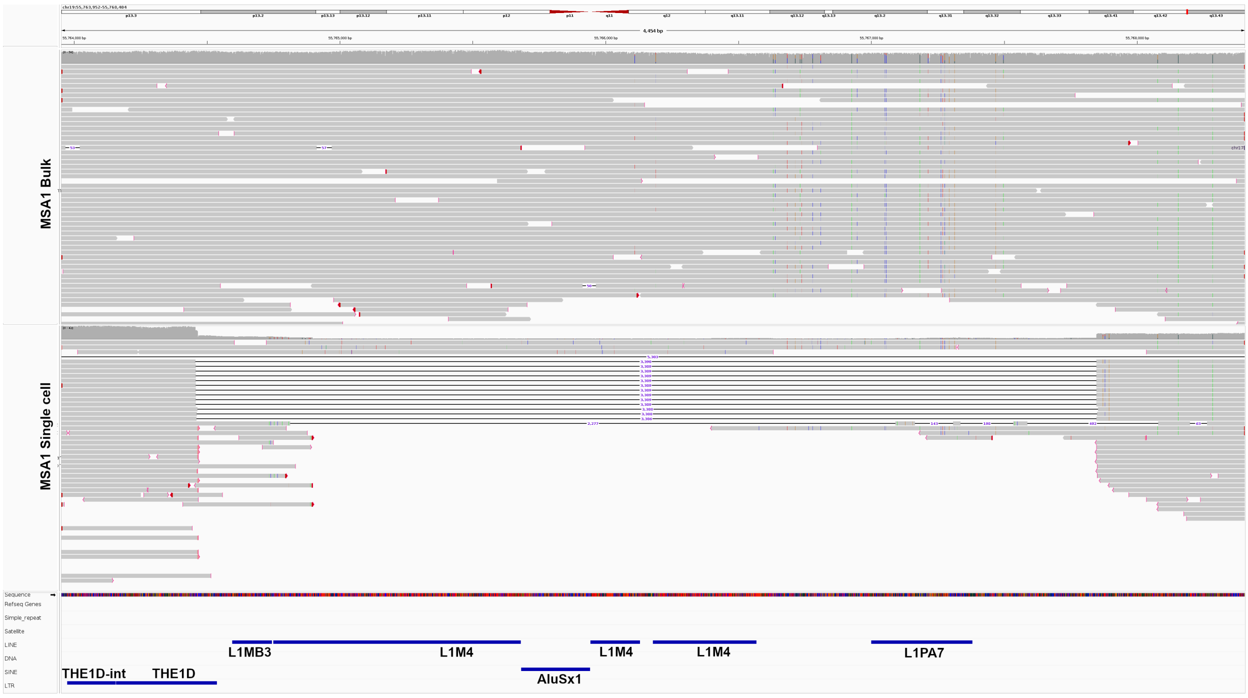

15. Example of a single cell-specific deletion detected in MSA1 brain.

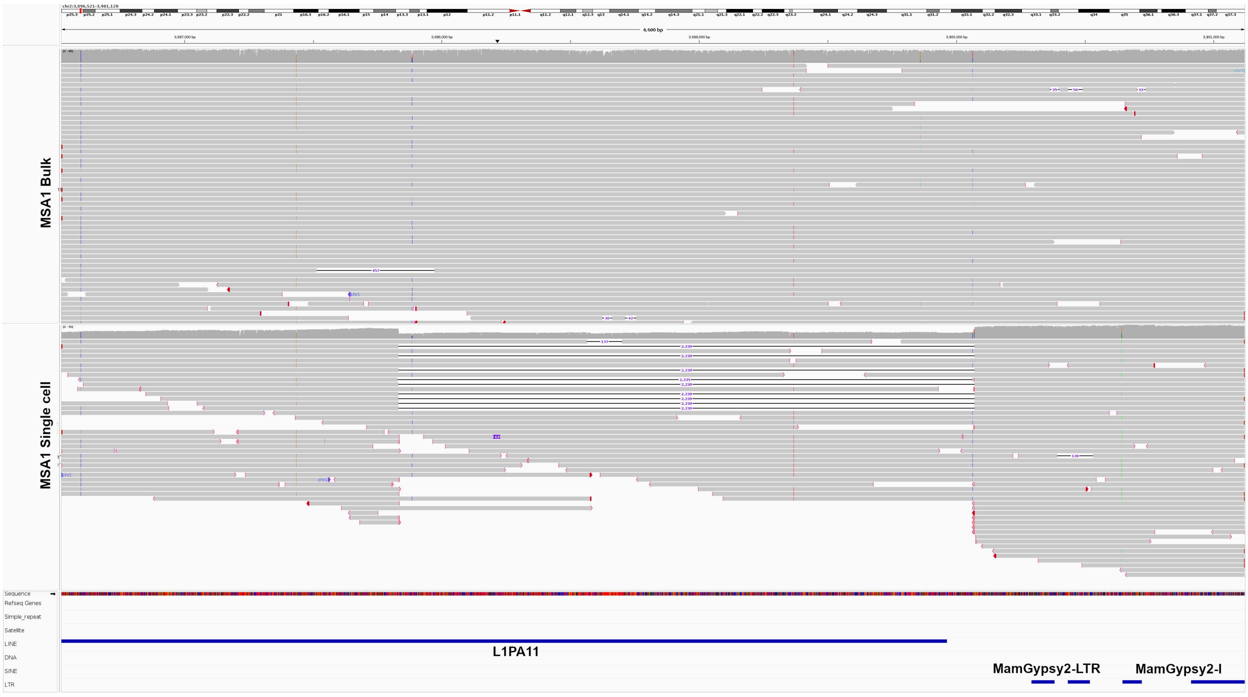


16. Example of a single cell-specific deletion detected in MSA1 brain.

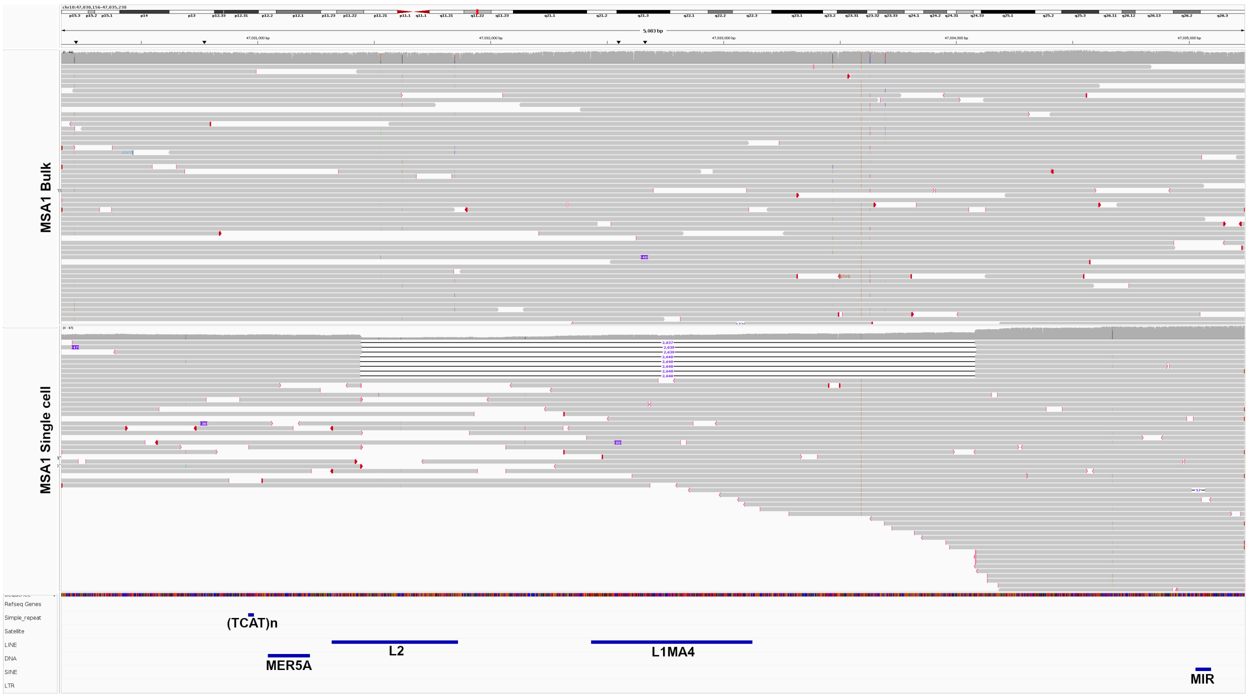

17. Example of a single cell-specific deletion detected in MSA1 brain.

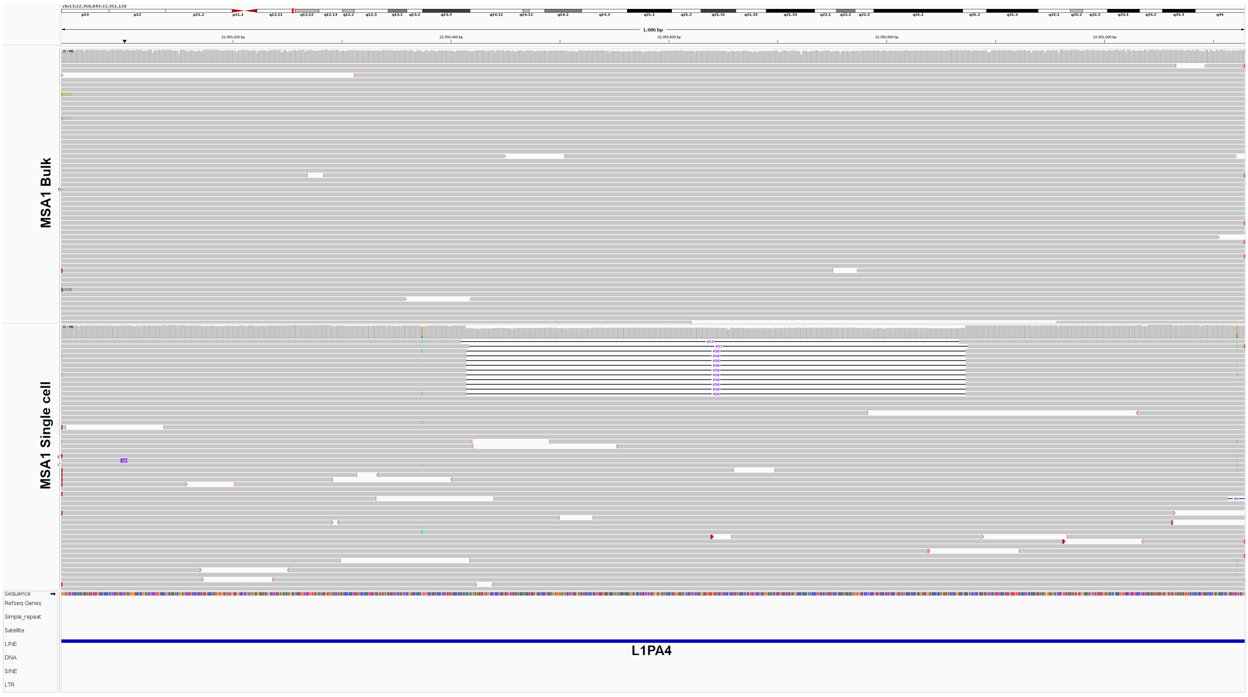


18. Example of a single cell-specific deletion detected in MSA1 brain.

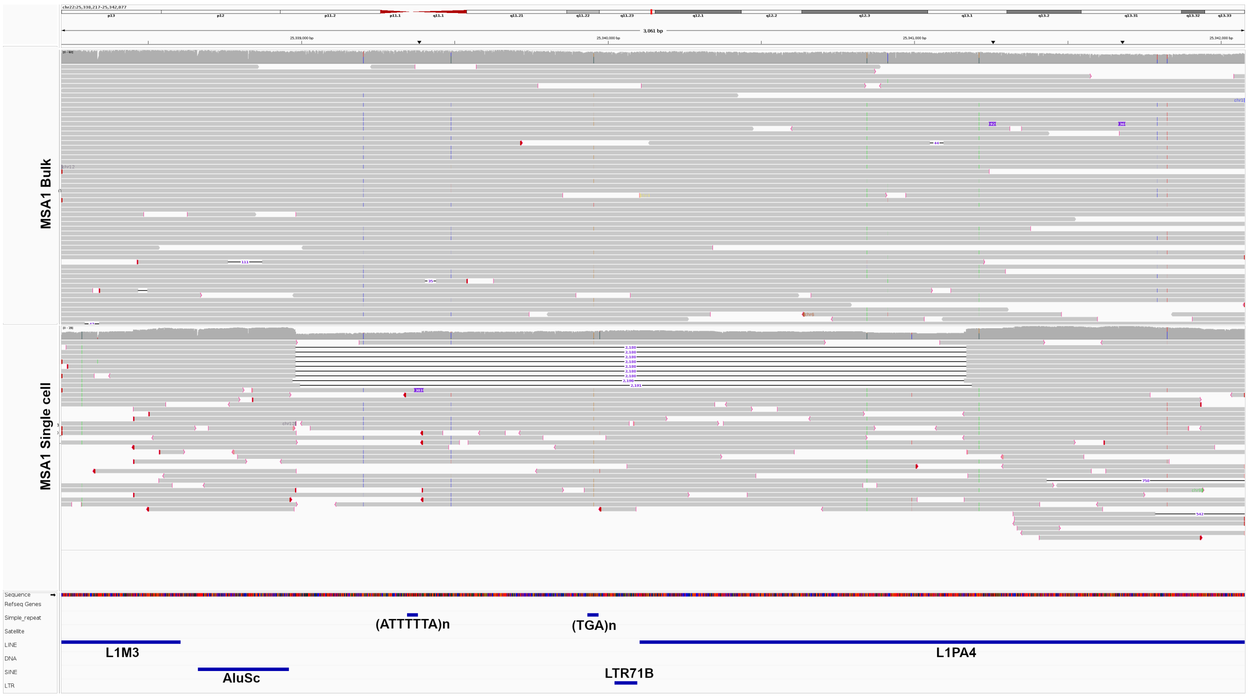

19. Example of a single cell-specific deletion detected in MSA1 brain.

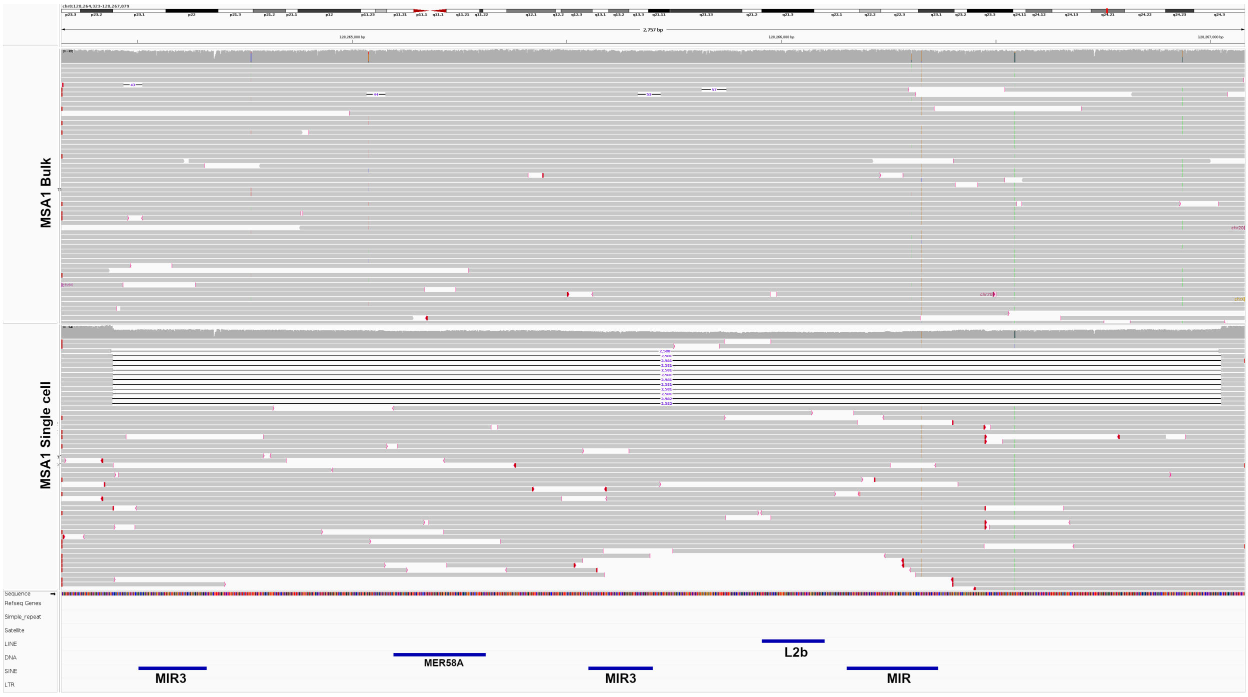


20. Example of a single cell-specific deletion detected in MSA2 brain.

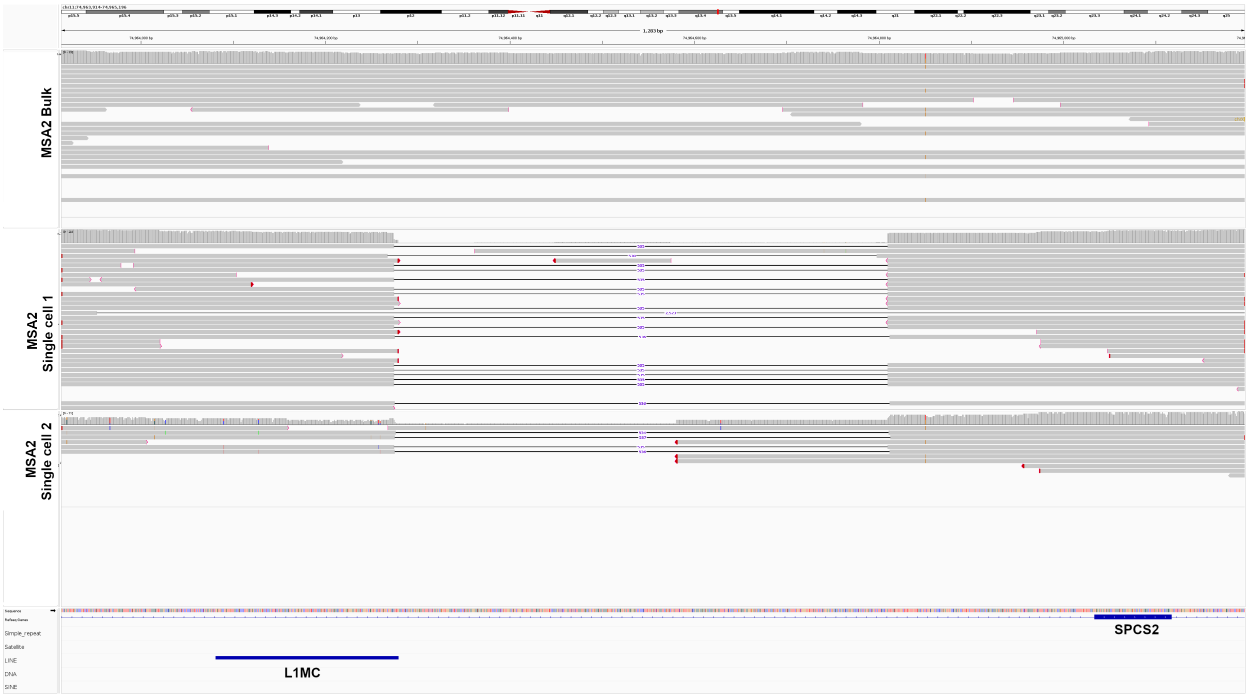

#### 21. Example of a single cell-specific deletion detected in MSA2 brain.

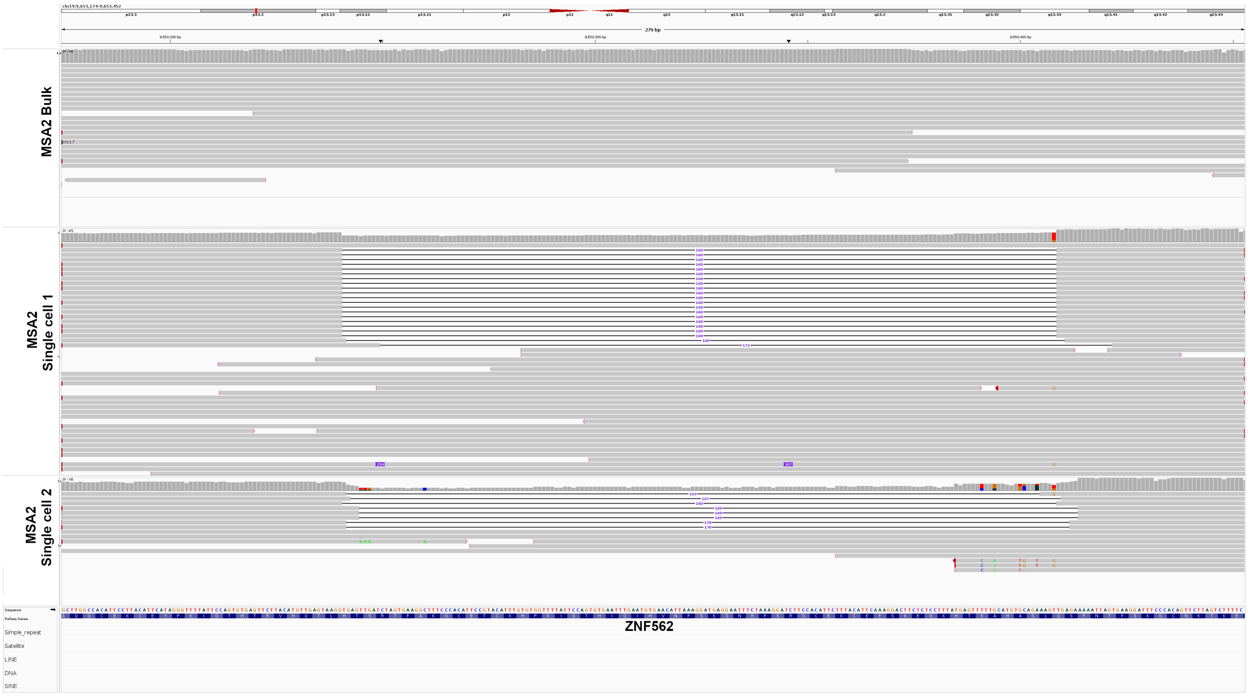


#### 22. Example of a single cell-specific deletion detected in MSA2 brain.

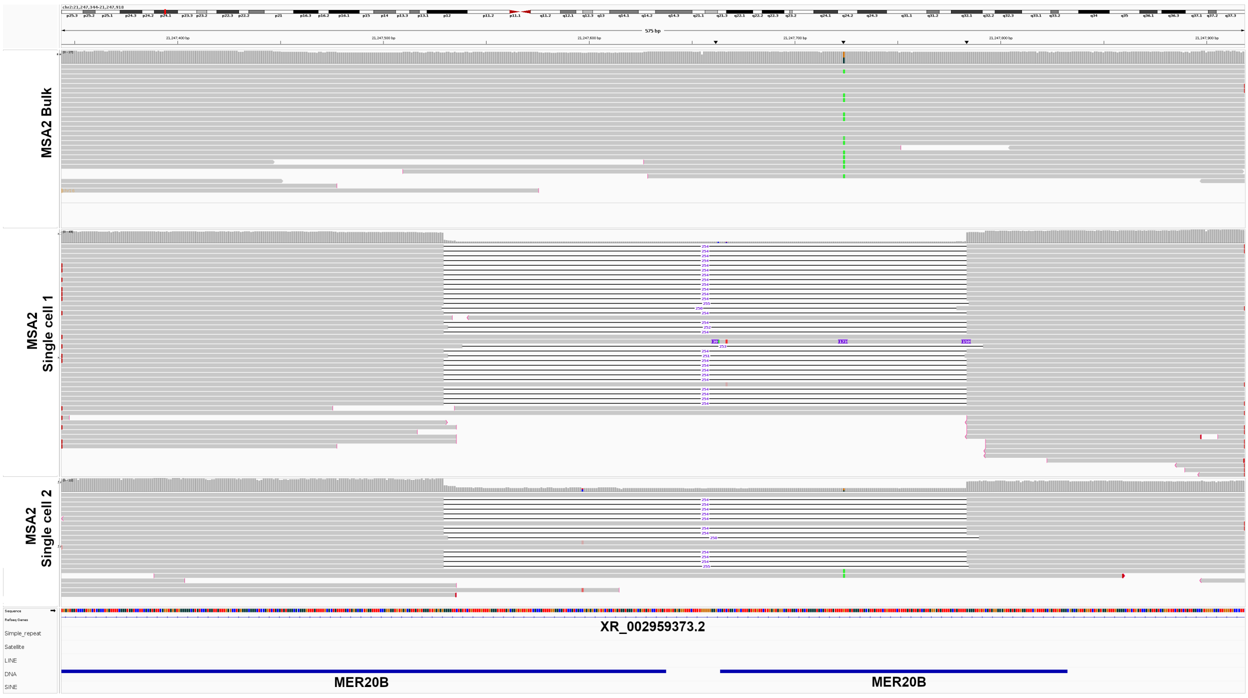

23. Example of a single cell-specific deletion detected in MSA2 brain.

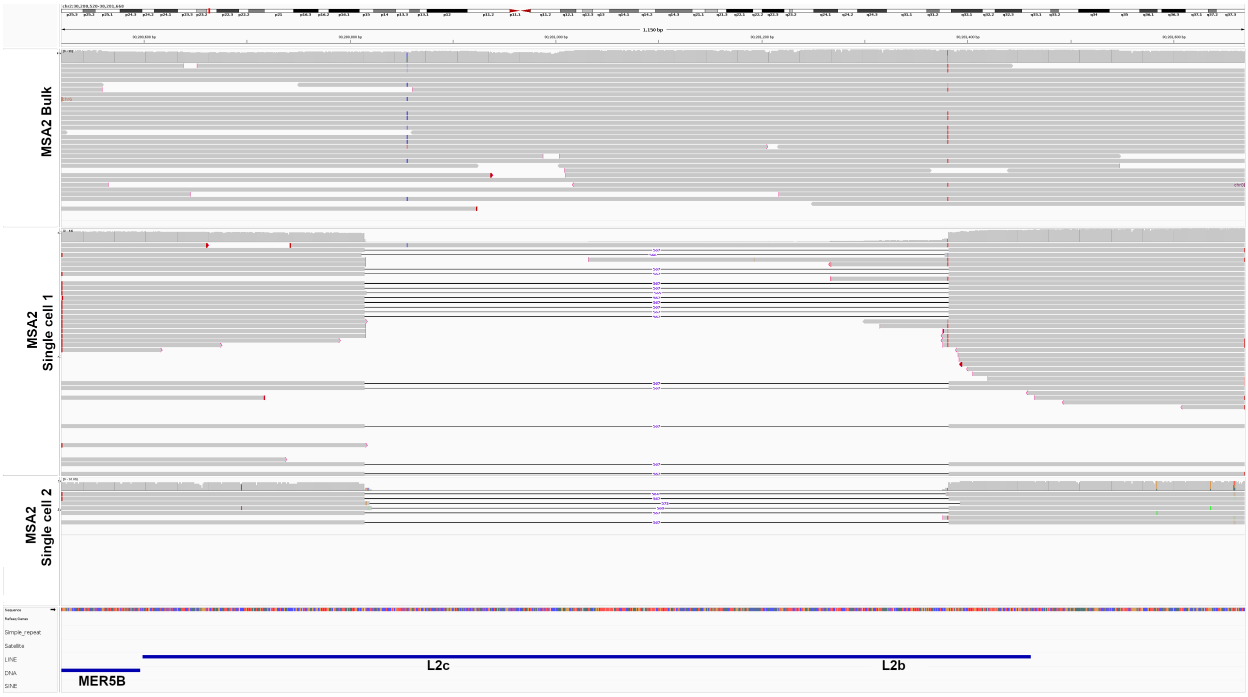


24. Example of a single cell-specific deletion detected in MSA2 brain.

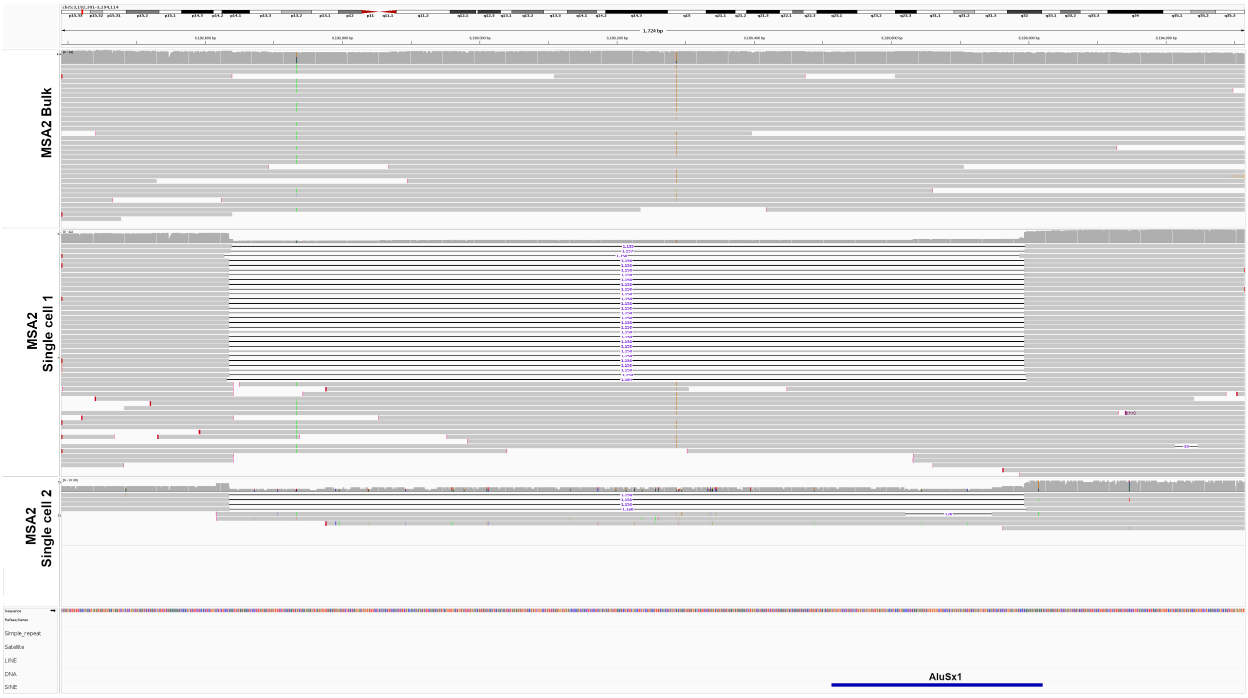

25. Example of a single cell-specific deletion detected in MSA2 brain.

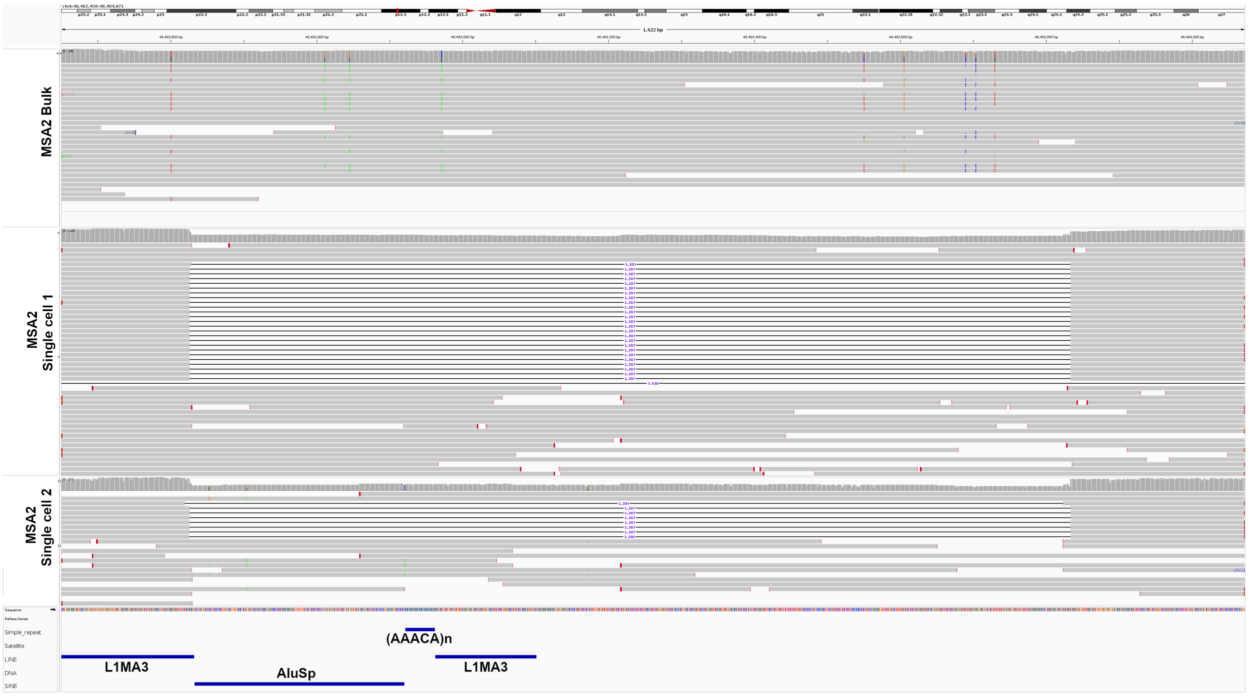


26. Example of a single cell-specific deletion detected in MSA2 brain.

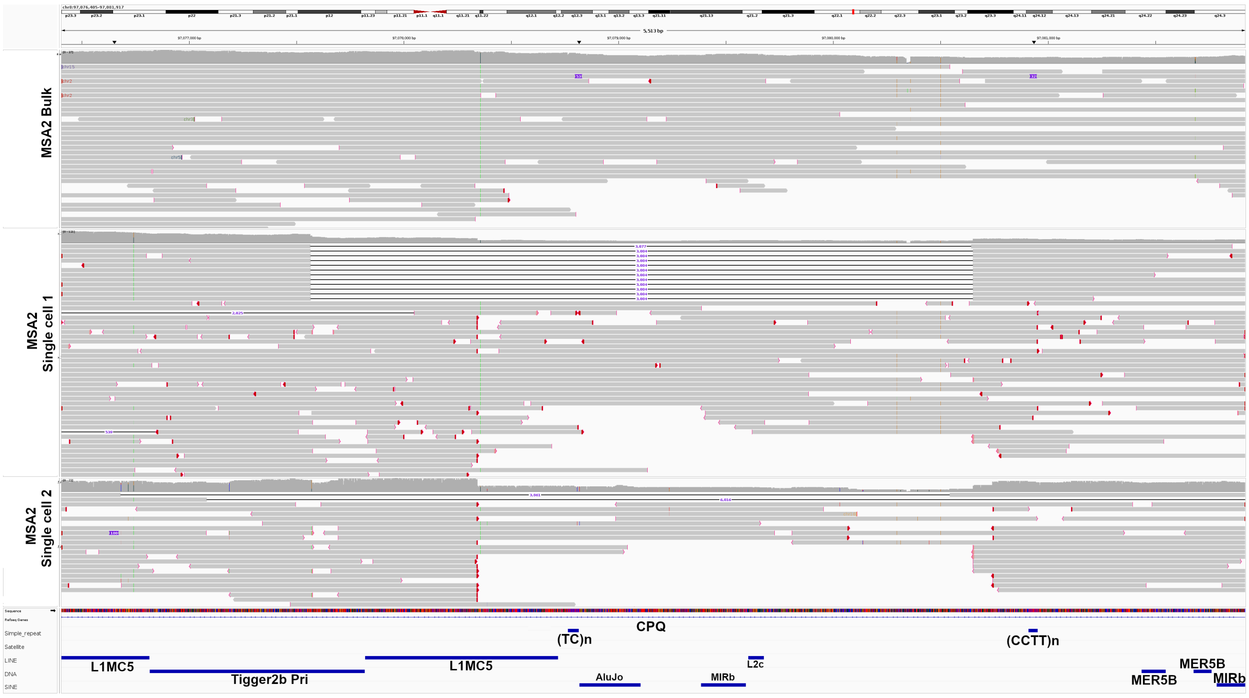

27. Example of a single cell-specific deletion detected in MSA2 brain.

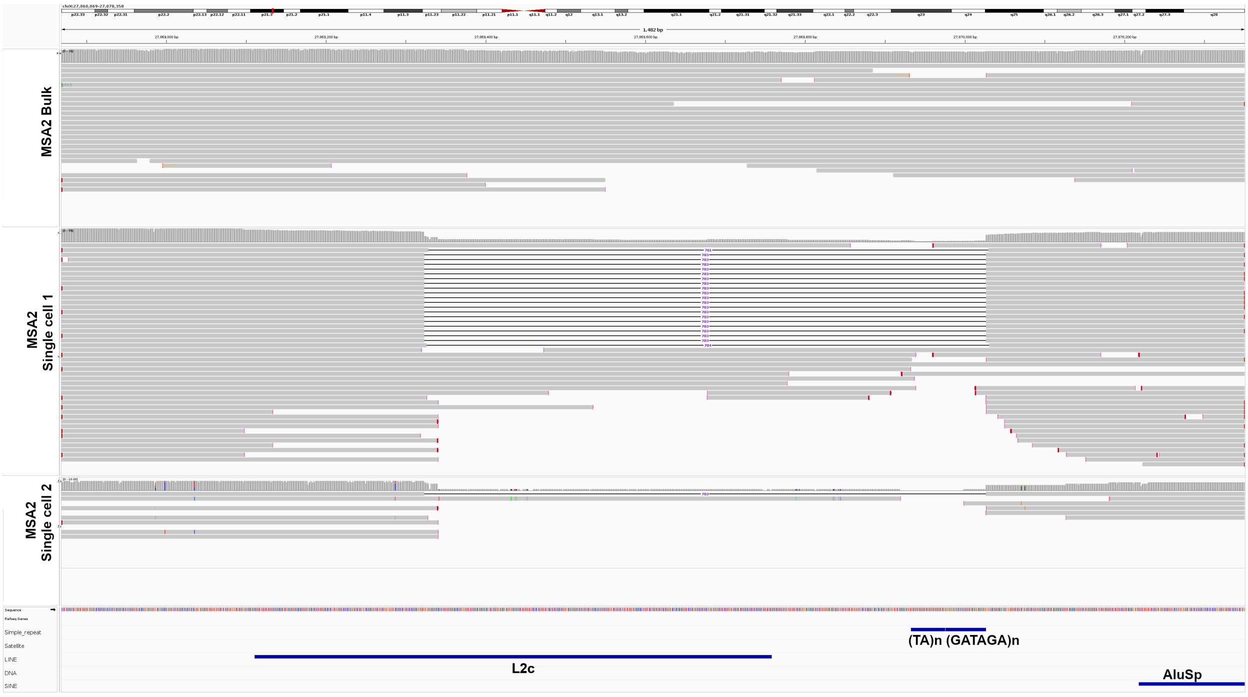


28. Example of a single cell-specific deletion detected in control brain.

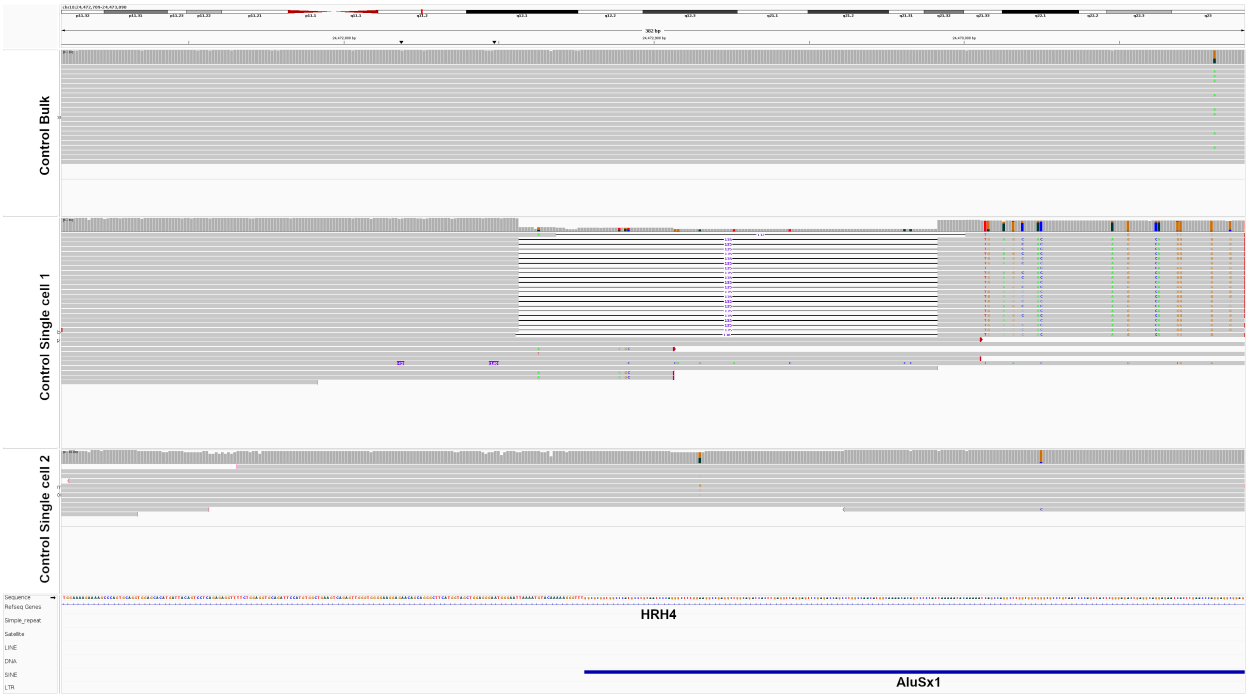

29. Example of a single cell-specific deletion detected in control brain.

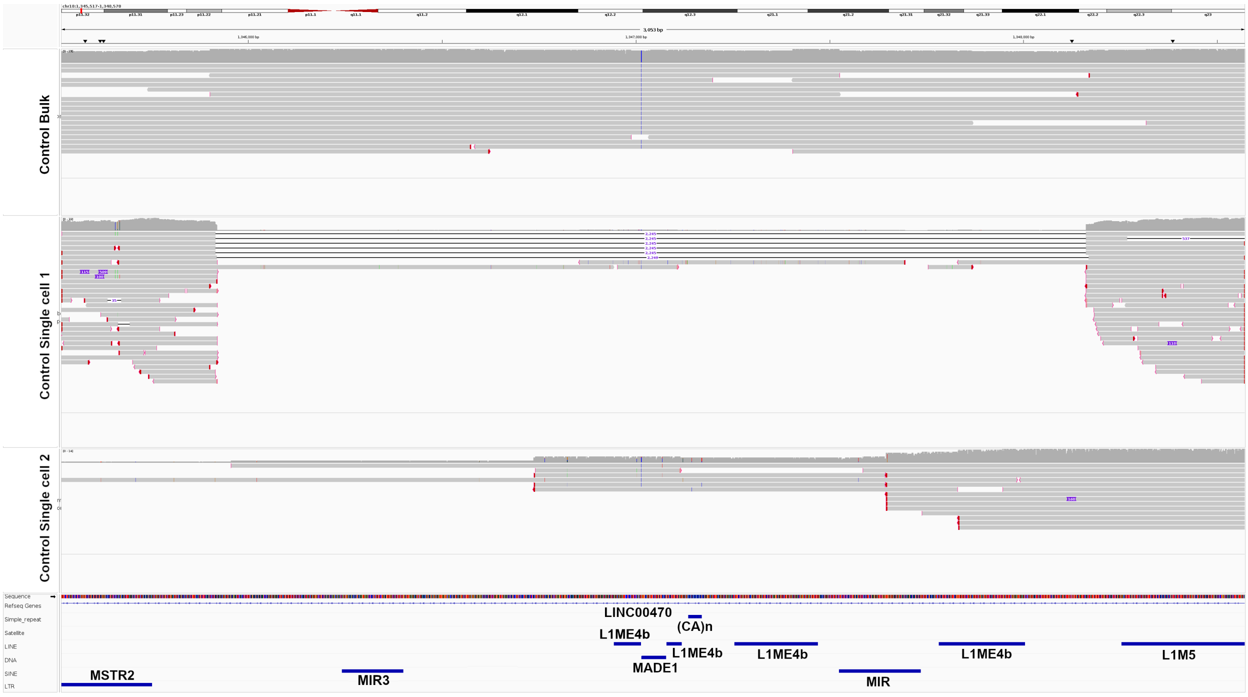


30. Example of a single cell-specific deletion detected in control brain.

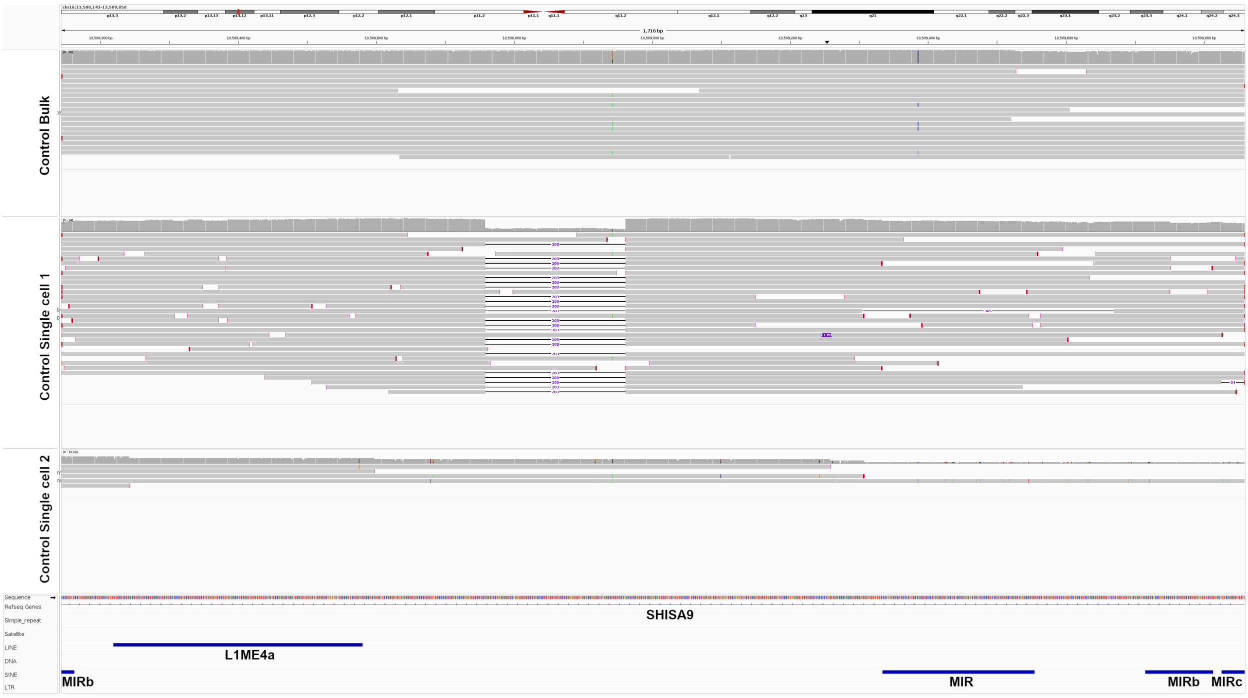

#### 31. Example of a single cell-specific deletion detected in control brain.

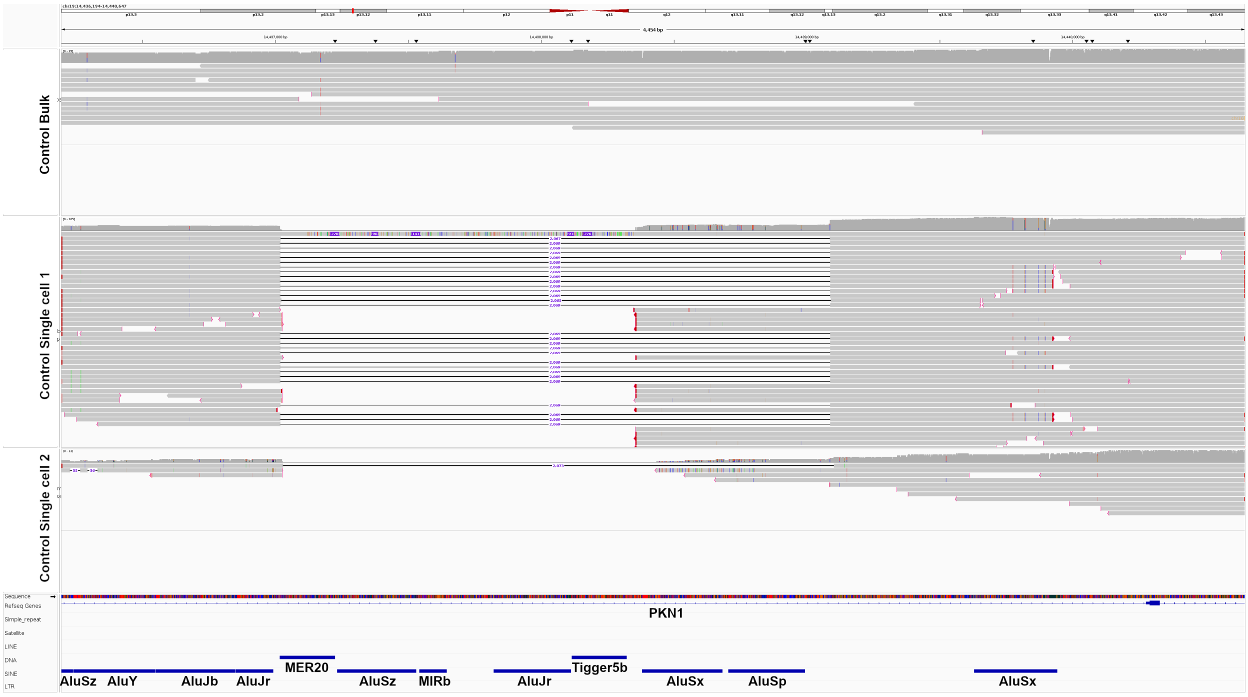


#### 32. Example of a single cell-specific deletion detected in control brain.

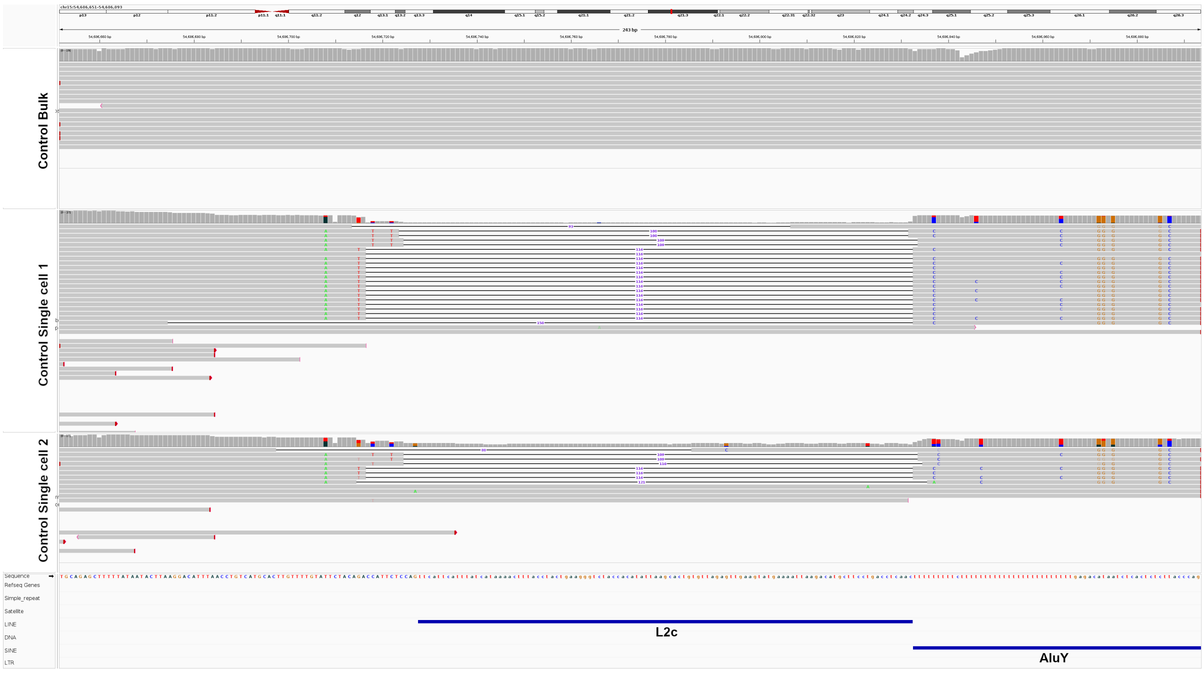

#### 33. Example of a single cell-specific deletion detected in control brain.

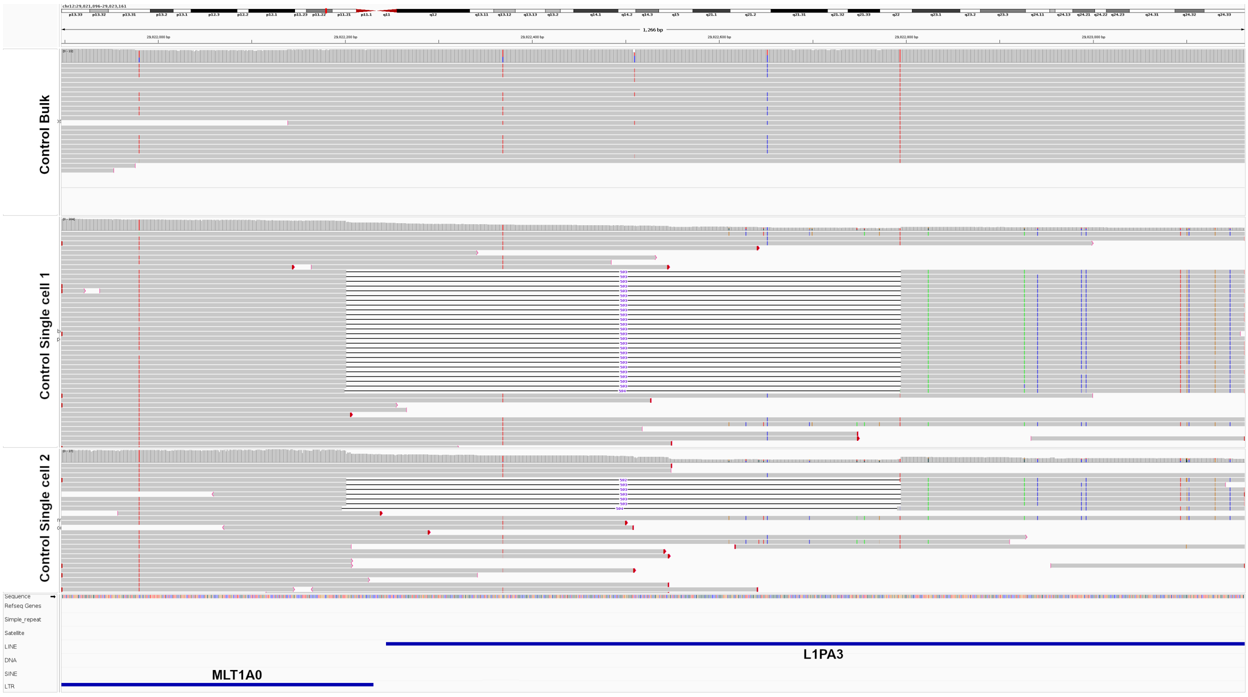


#### 34. Example of a single cell-specific deletion detected in control brain.

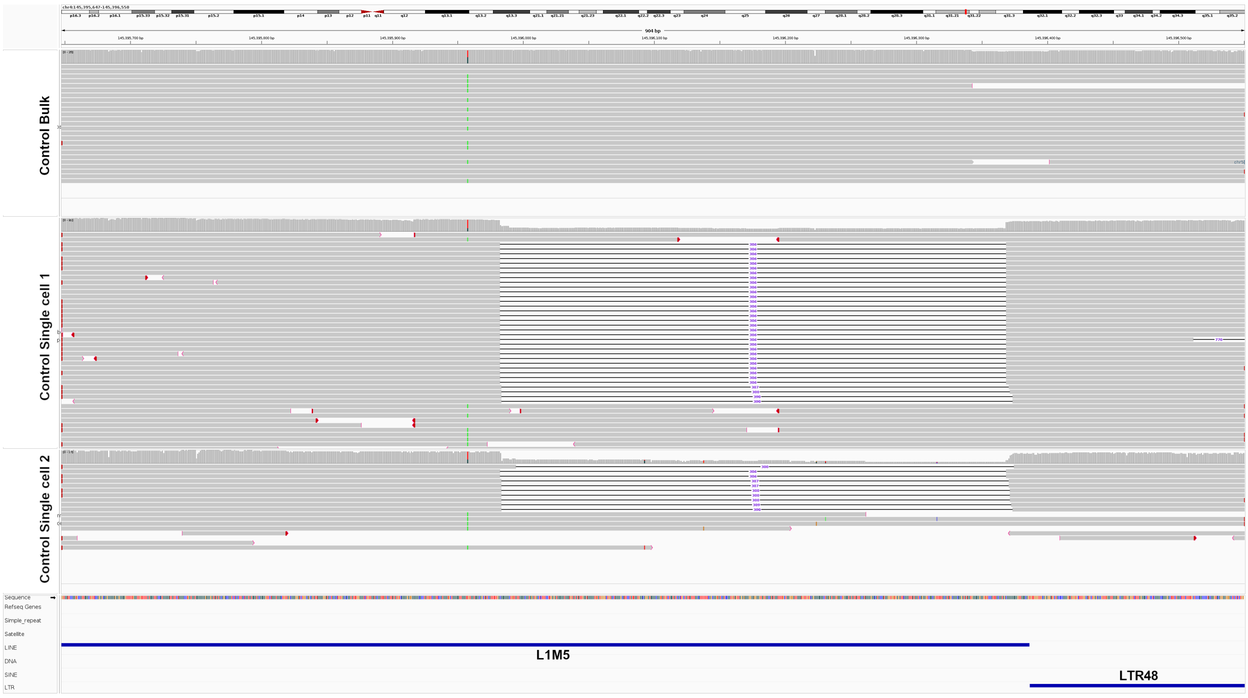

35. Example of a single cell-specific deletion detected in control brain.

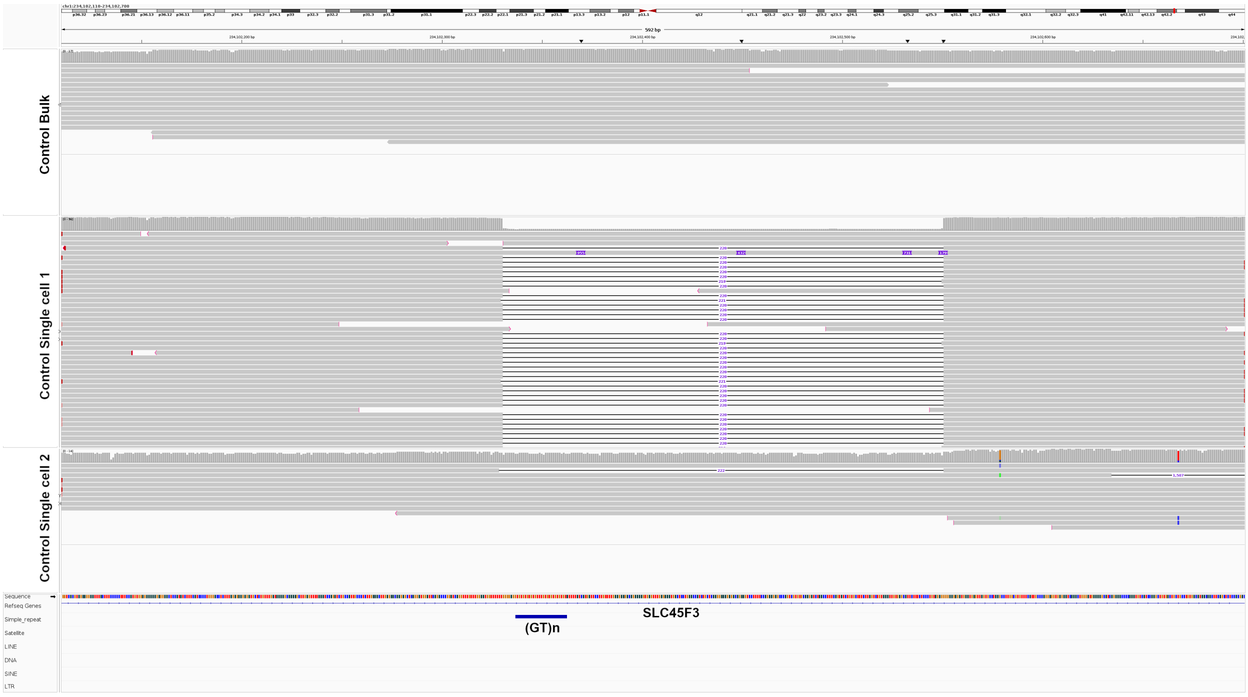


36. Example of recombinational deletion between two SINE/Alu elements (AluY - AluJb) – MSA1.

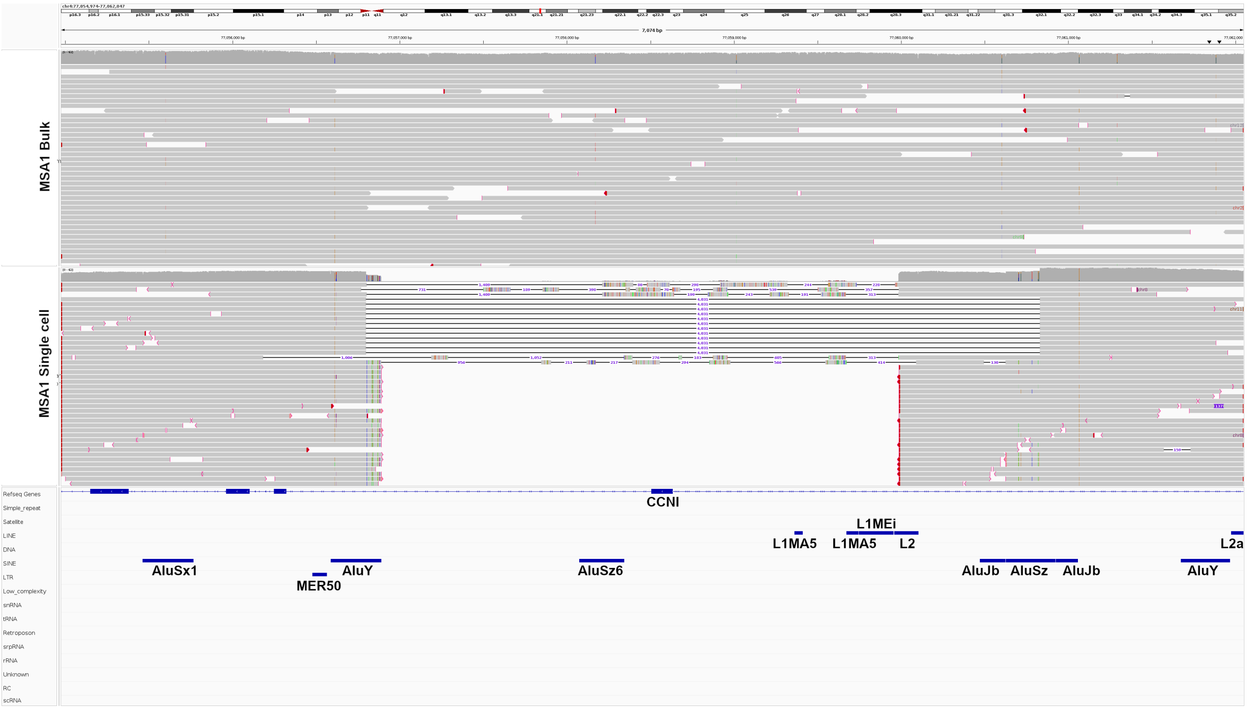

37. Example of recombinational deletion between two SINE/Alu (AluJo – AluJb) elements – MSA2.

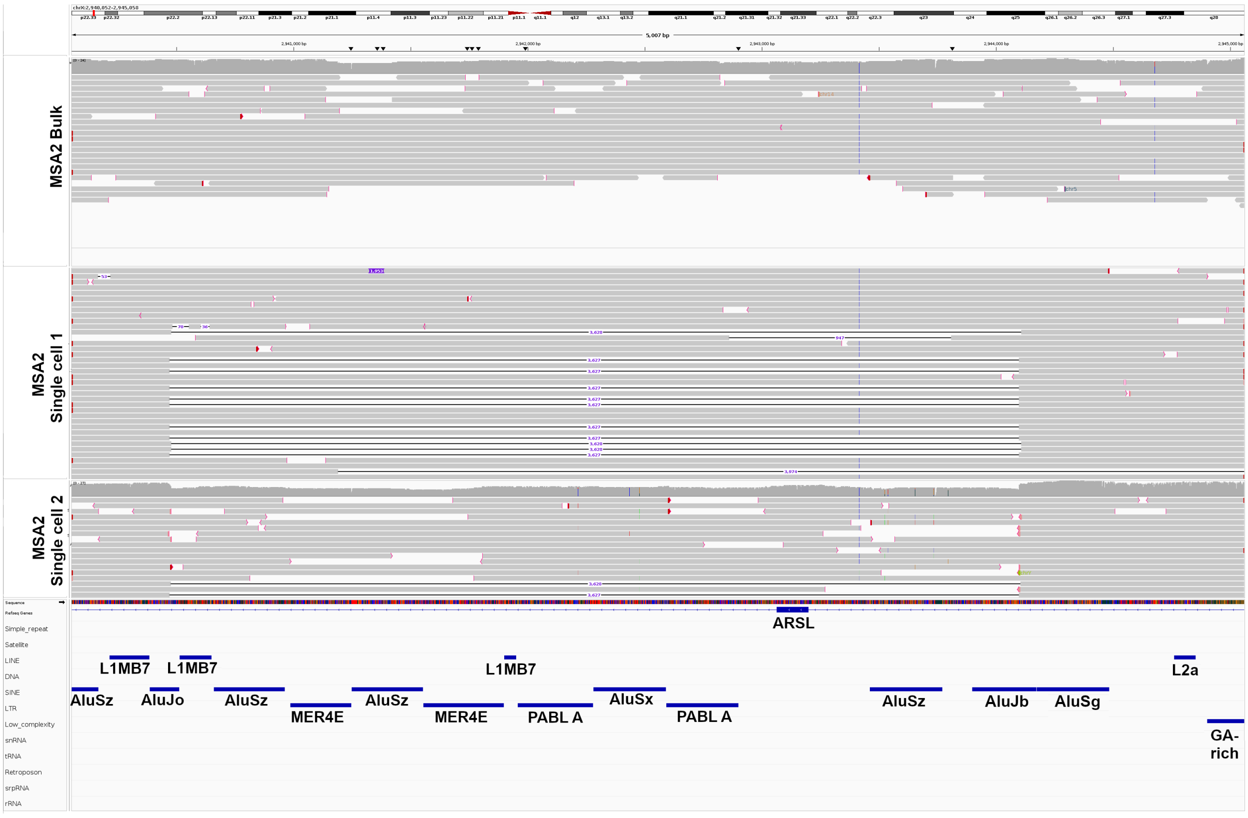


38. Example of recombinational deletion between two SINE/Alu (AluSz6 – AluSz6) elements – MSA2.

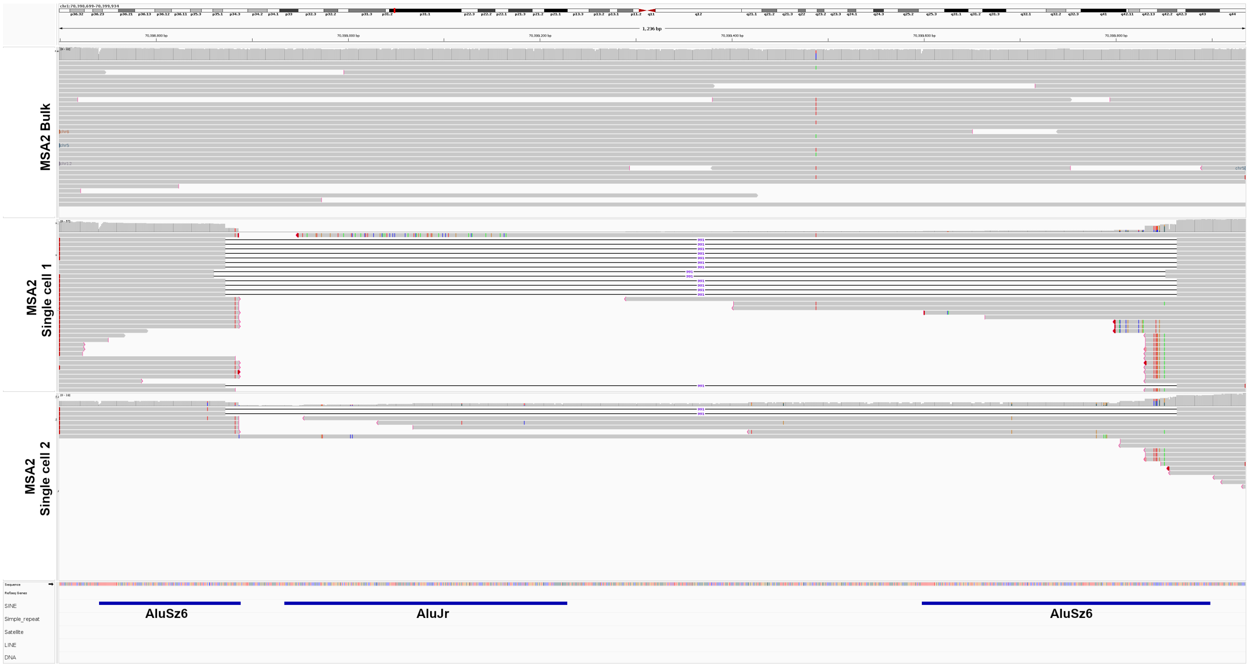

39. Example of recombinational deletion between two LINE/L1 (L1M4 – L1MC) elements – MSA1.

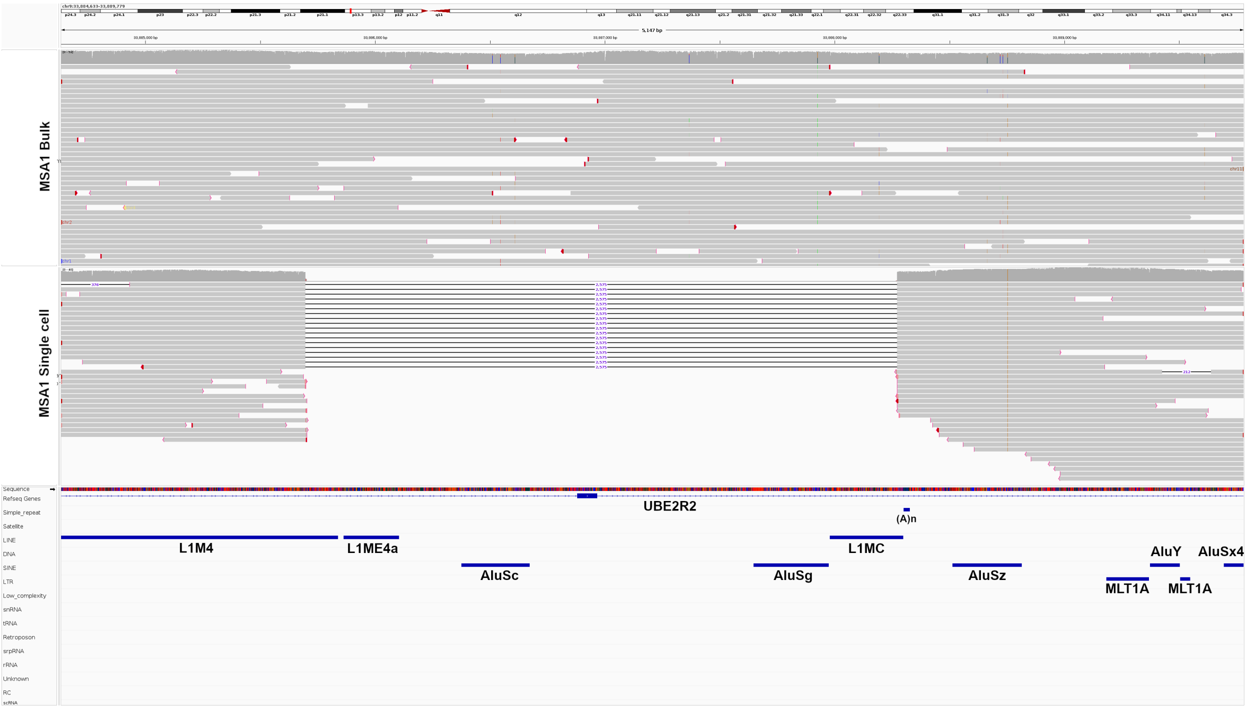


40. Example of recombinational deletion between two LINE/L1 (L1MEg – L1MEg) elements – MSA2.

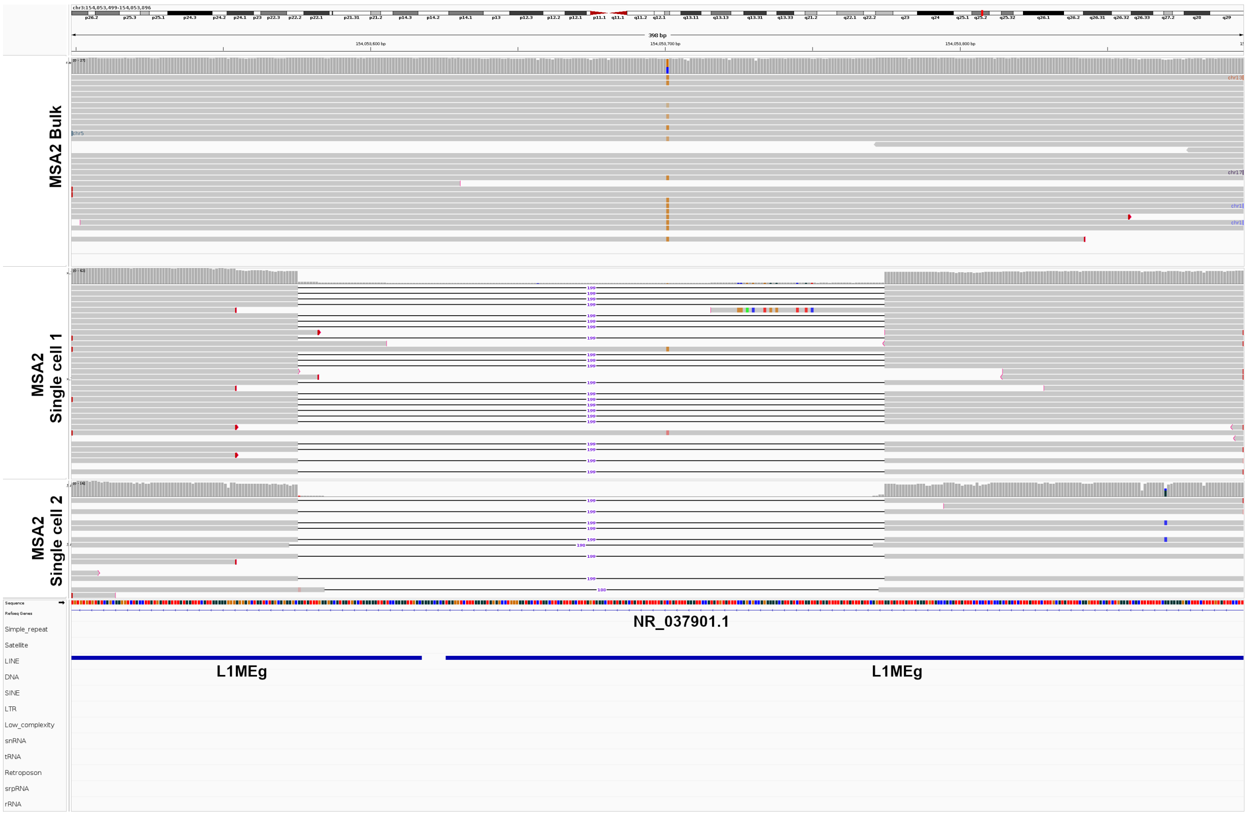

41. Example of recombinational deletion between two LINE/L1 (L1MA2 – L1M5) elements – MSA2.

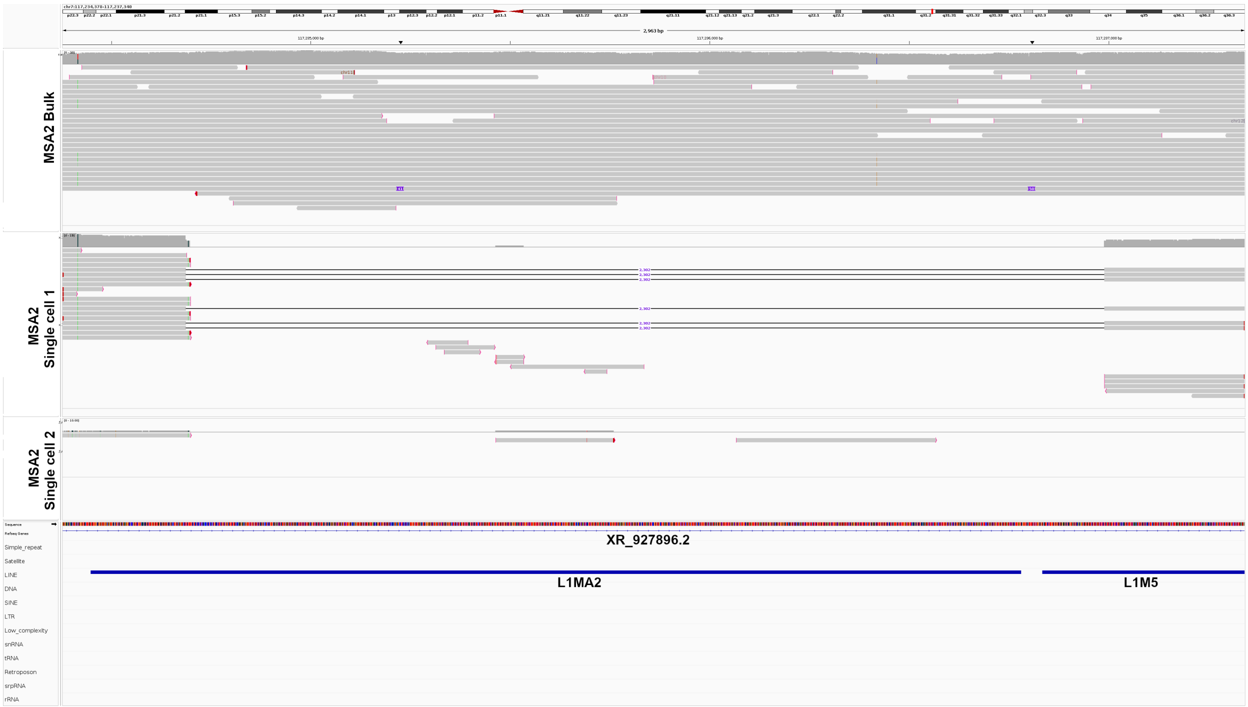


42. A) MSA1 T7 read length distribution, limited to 70,000bp; the MSA1 T7-6 sample peaks at ~90,000bp. Due to low yield, we omit these samples from the analysis.

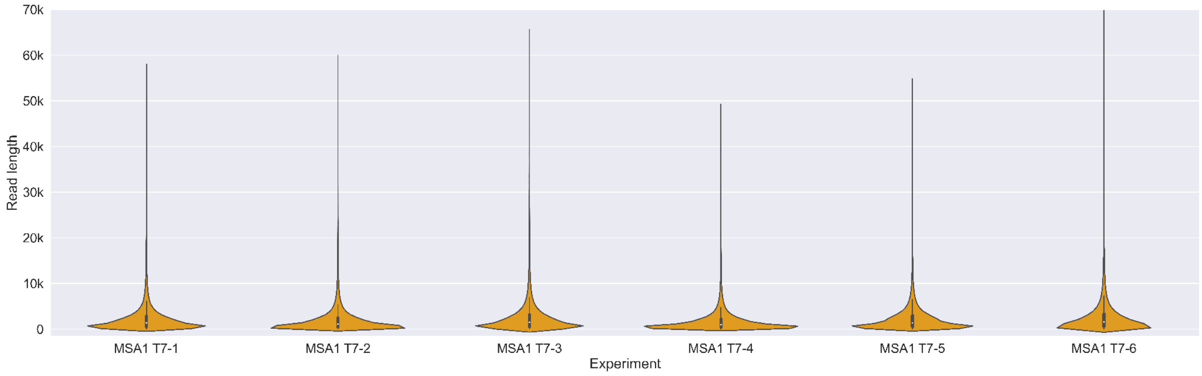


B) MSA1 T7 read length distribution, limited to 10,000bp.

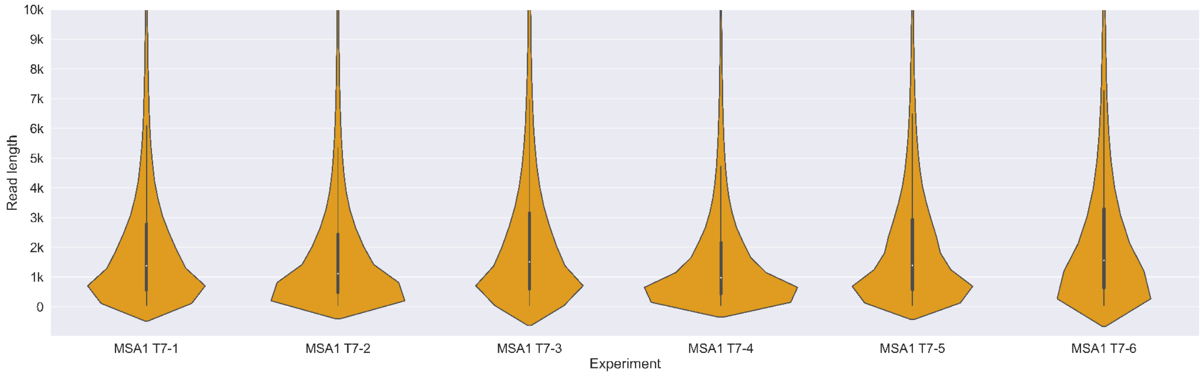

43. A) MSA1 RBP read length distribution, limited to 70,000bp. We sequenced 6 single cells twice with MinION device.

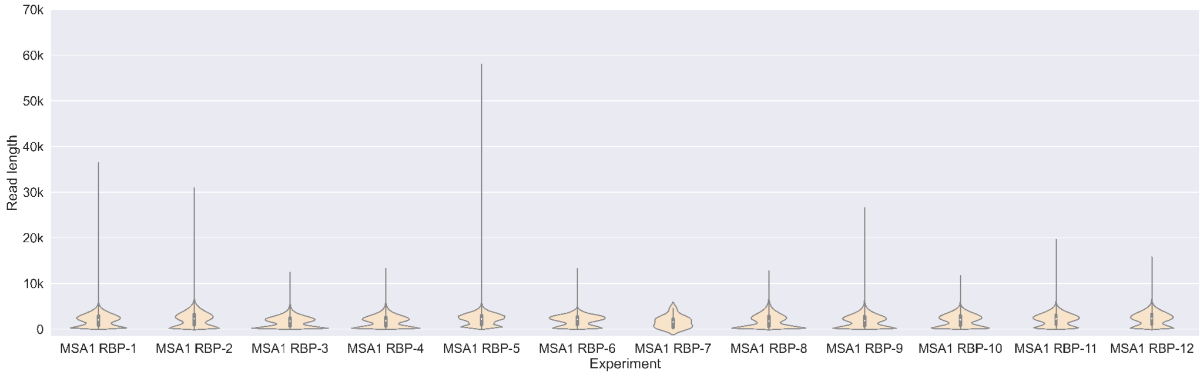


B) MSA1 RBP read length distribution, limited to 10,000bp.

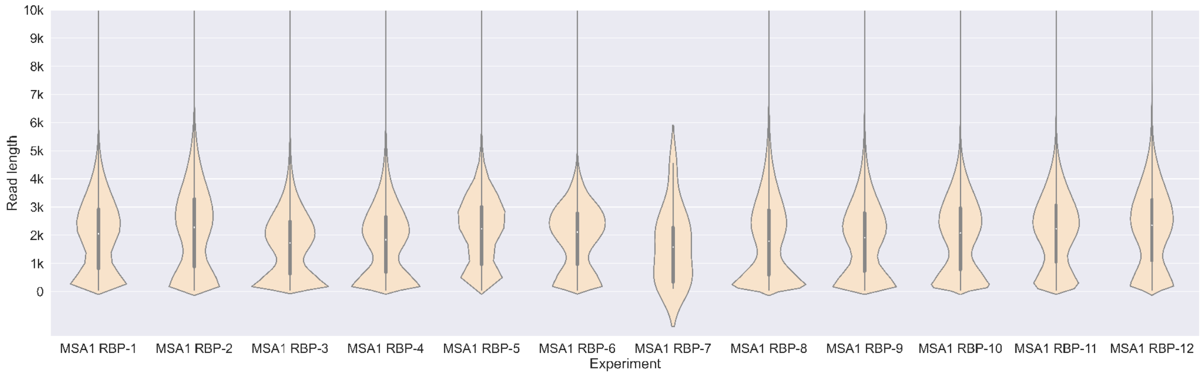


44. A) MSA2 RBP read length distribution, limited to 70,000bp; the MSA2 RBP-3 sample longest read is at ~1,500,000bp.

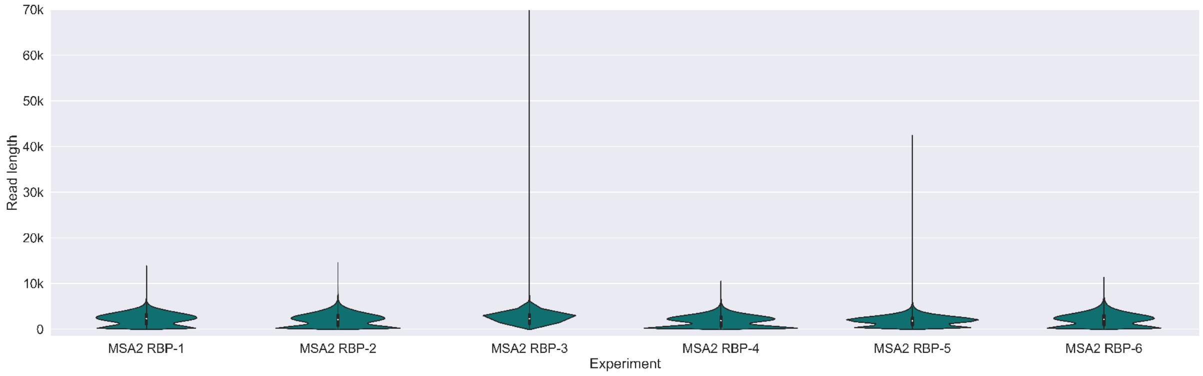


B) MSA2 RBP read length distribution, limited to 10,000bp

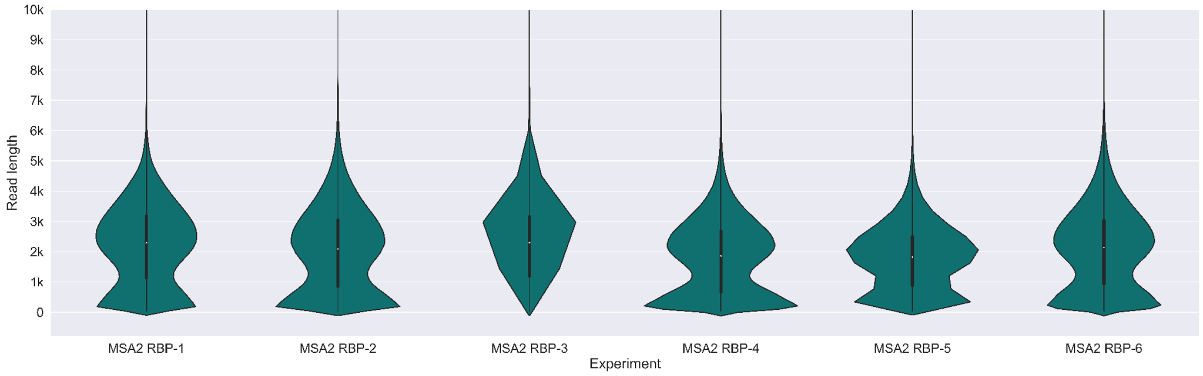

45. A) MSA2 T7 read length distribution, limited to 70,000bp; the MSA2 T7-1 sample peaks at ~100,000bp, MSA2 T7-3 at ~120,000bp and MSA2 T7-4 at ~120,000bp

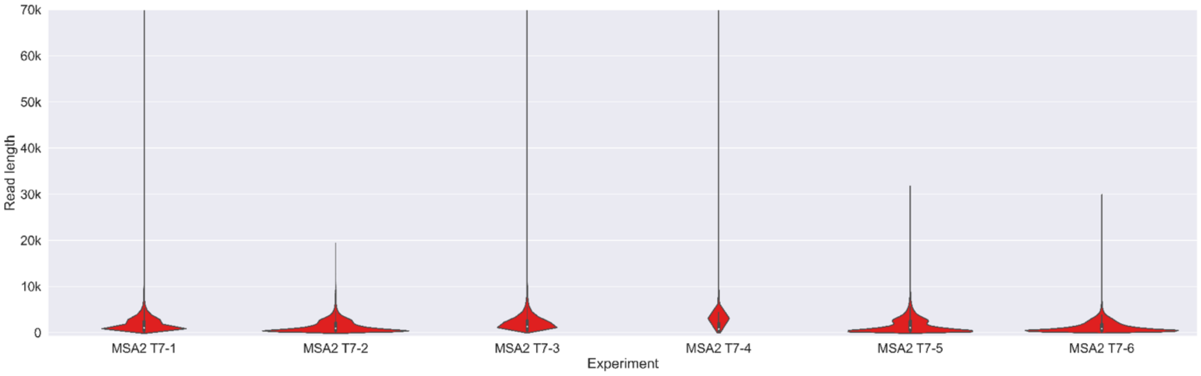


B) MSA2 T7 read length distribution, limited to 10,000bp.

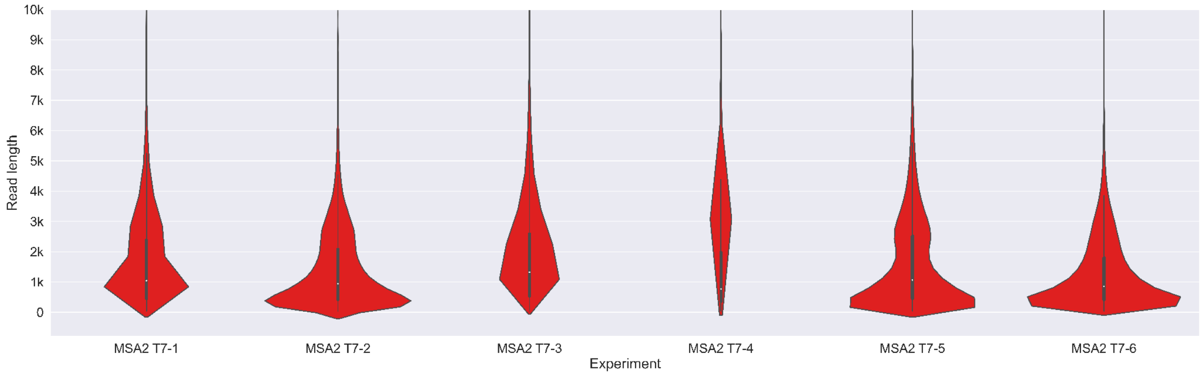


46. A) Control RBP read length distribution, limited to 70,000bp.

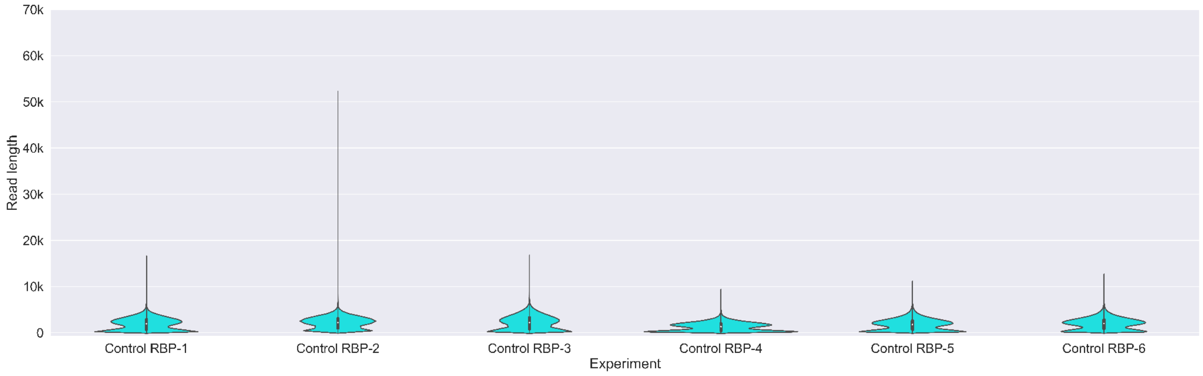


B) Control RBP read length distribution, limited to 10,000bp.

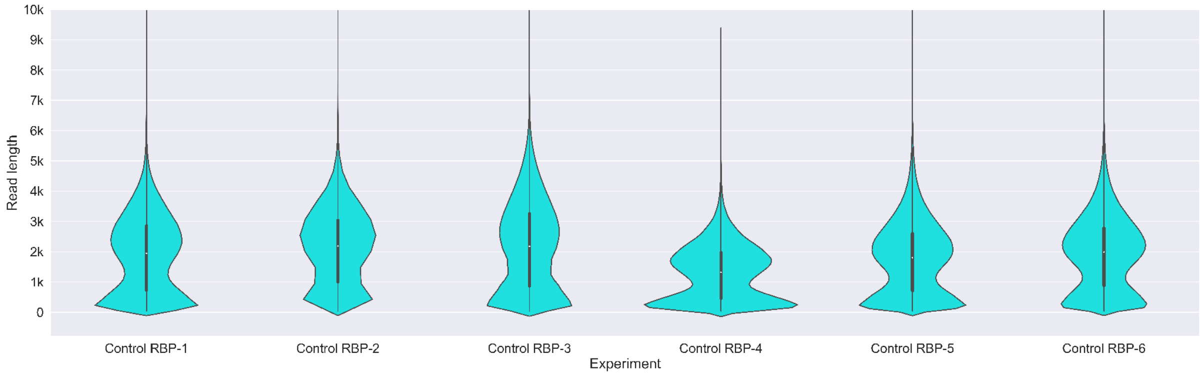

47. A) Control T7 read length distribution, limited to 70,000bp; the Control T7-2 sample peaks at ~100,000bp, Control T7-3 at ~90,000bp and Control T7-5 at ~140,000bp

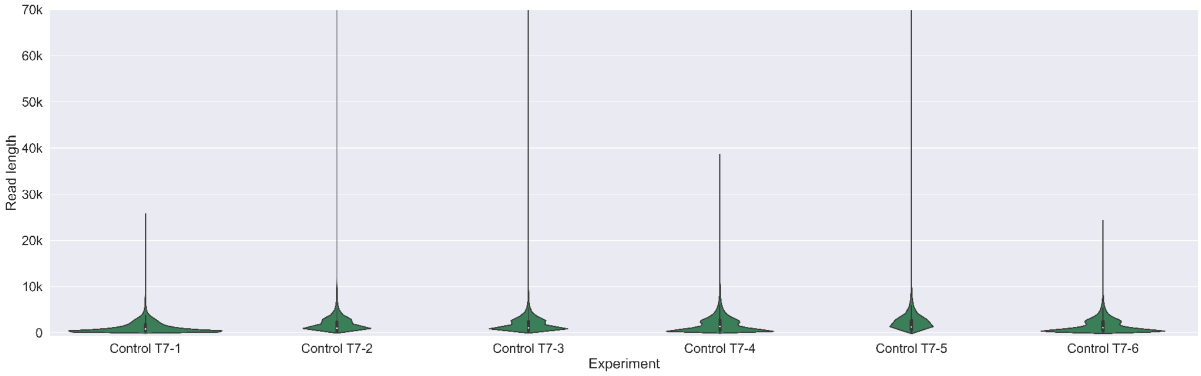


B) Control T7 read length distribution, limited to 10,000bp.

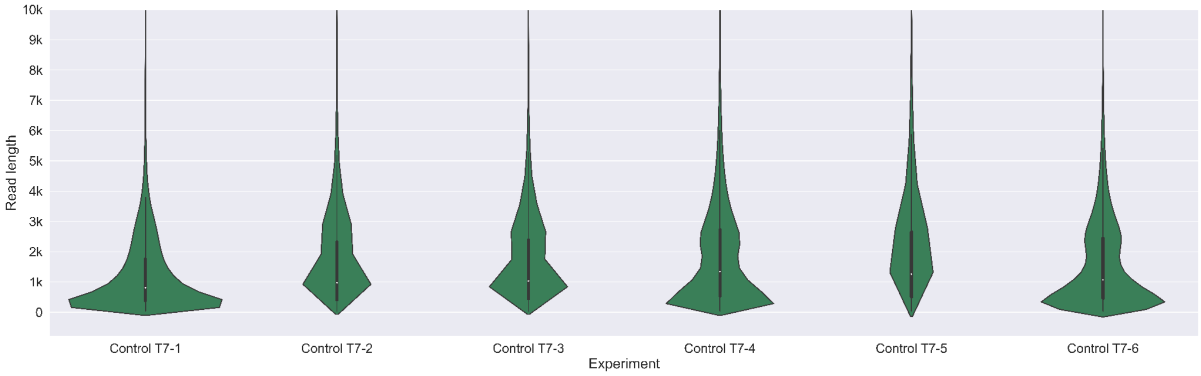

#### 48. Inversion and duplication sizes - unfiltered - MSA1 T7

A)

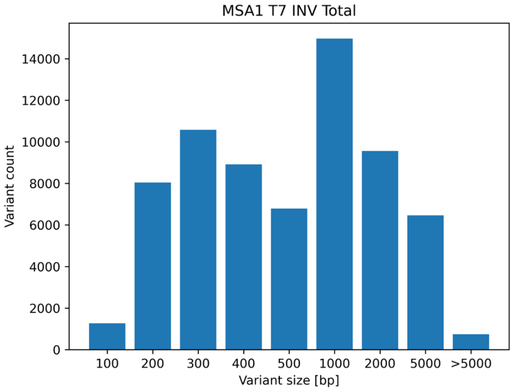


B)

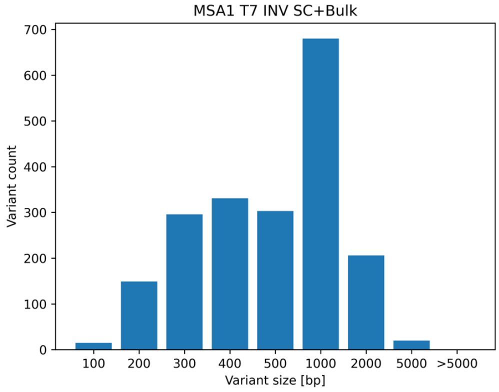


C)

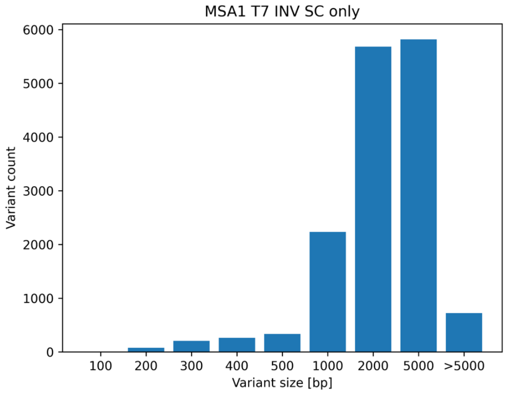


D)

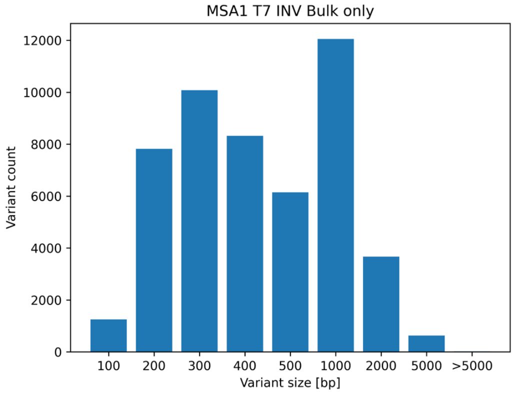


E)

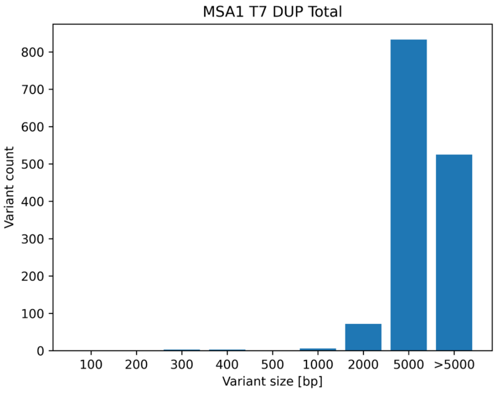


F)

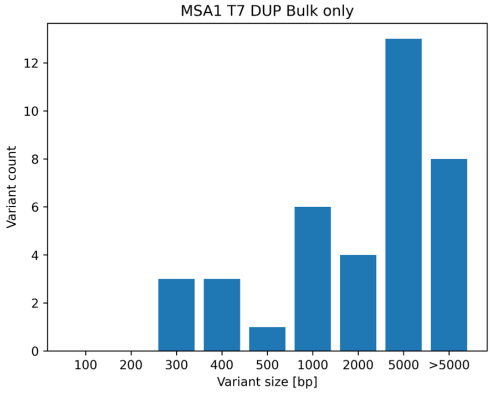


G)

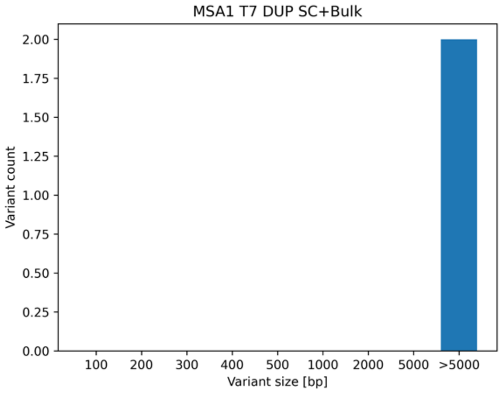


H)

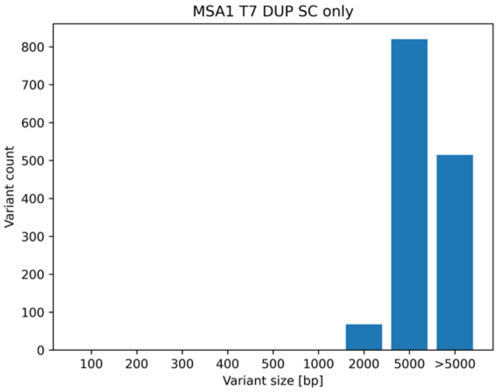

#### 49. Inversion and duplication sizes - unfiltered - MSA1 RBP

A)

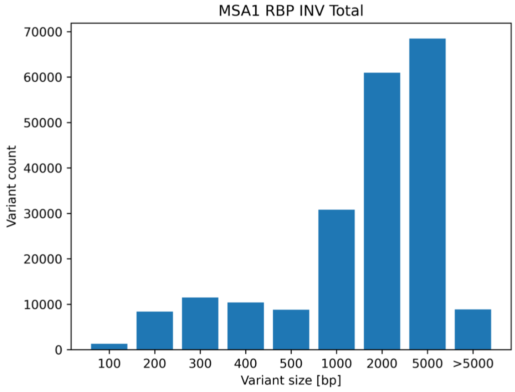


B)

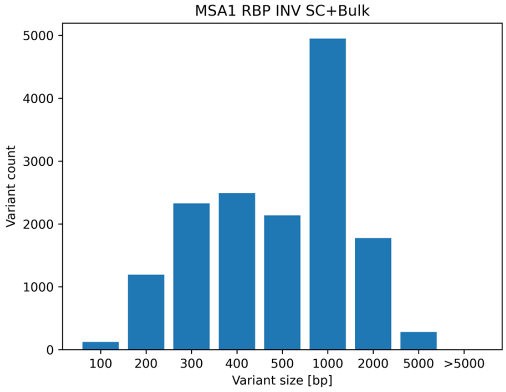


C)

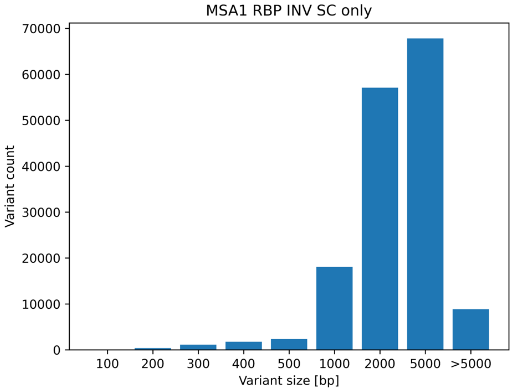


D)

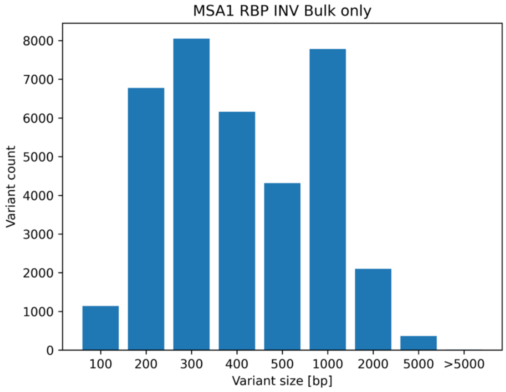


E)

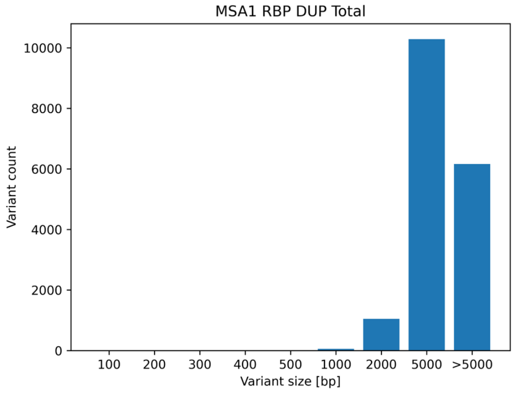


F)

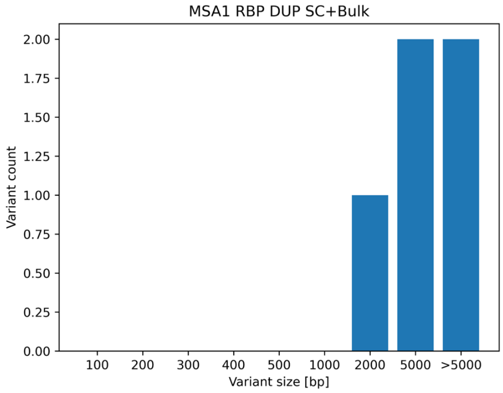


G)

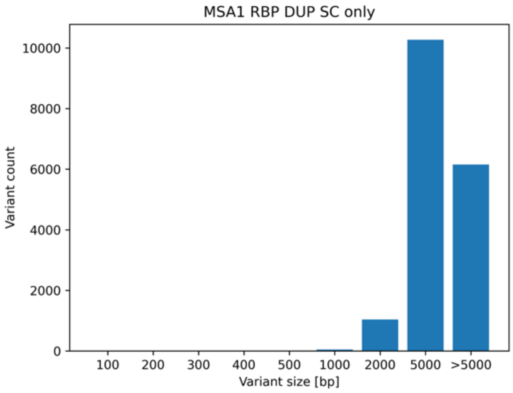


H)

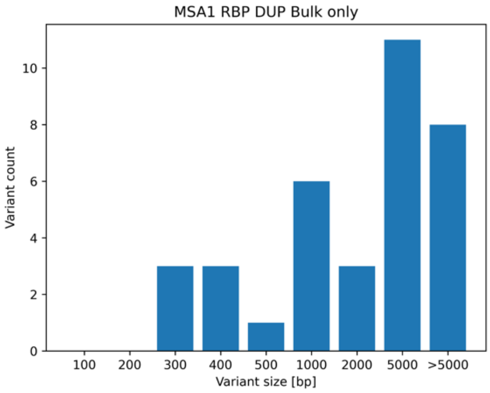

### 50. Inversion and duplication sizes - unfiltered - Control RBP

A)

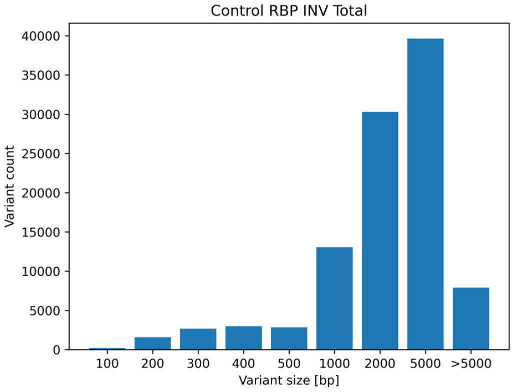


B)

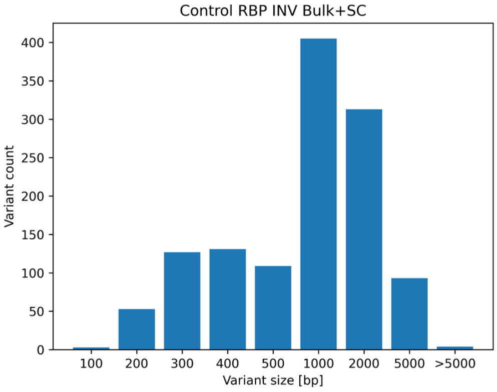


C)

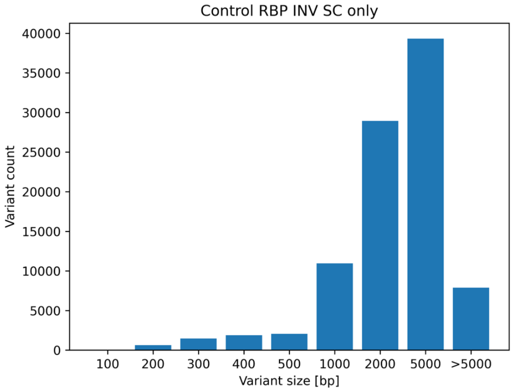


D)

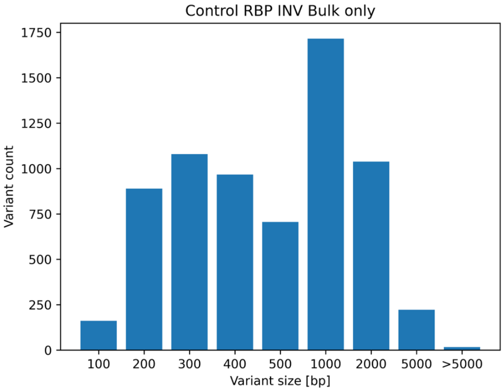


E)

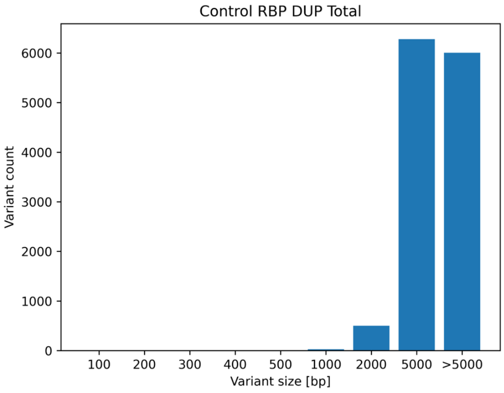


F)

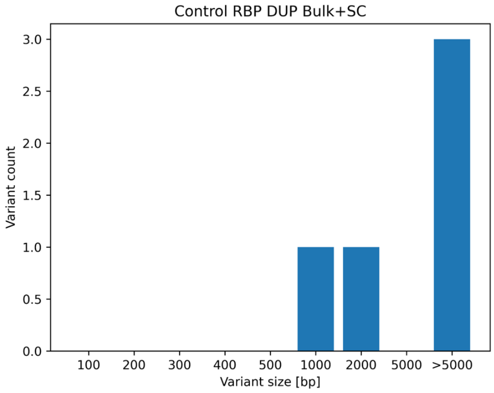


G)

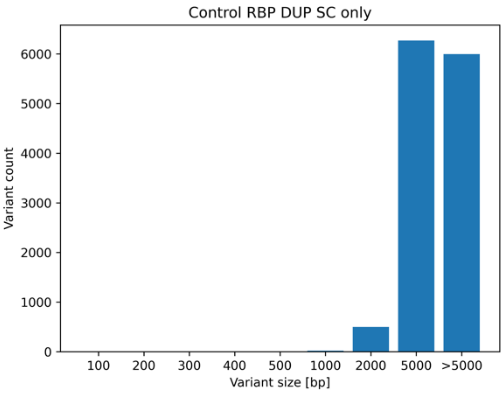


H)

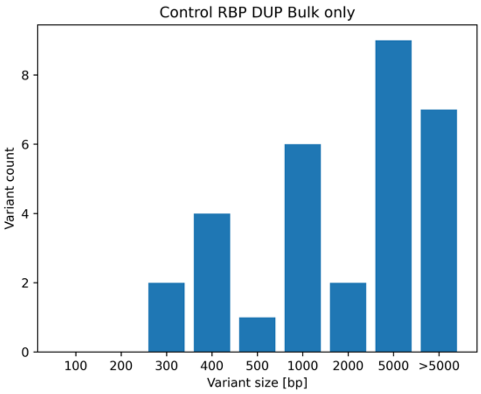

### 51. Inversion and duplication sizes - unfiltered - Control T7

A)

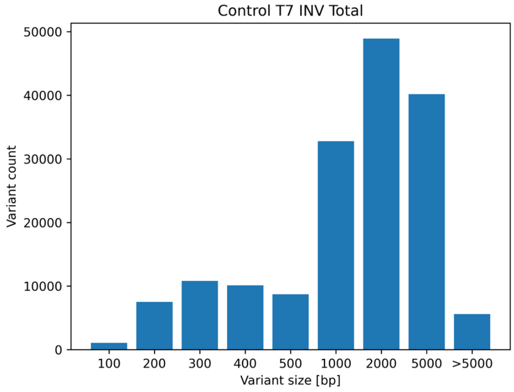


B)

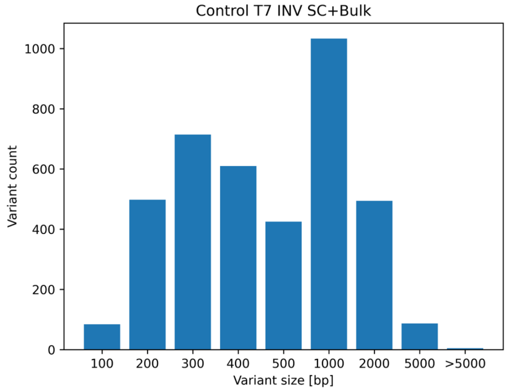


C)

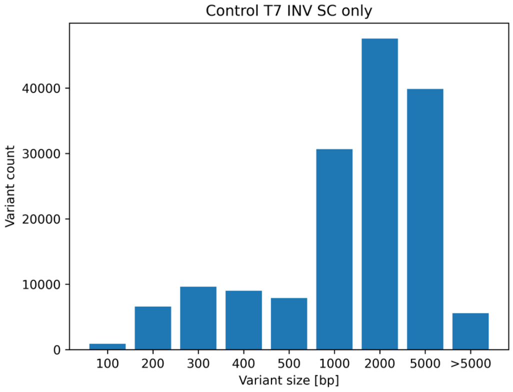


D)

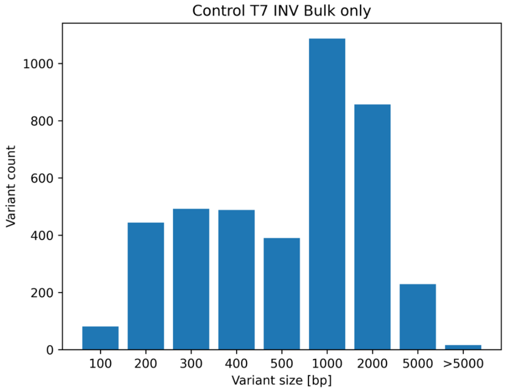


E)

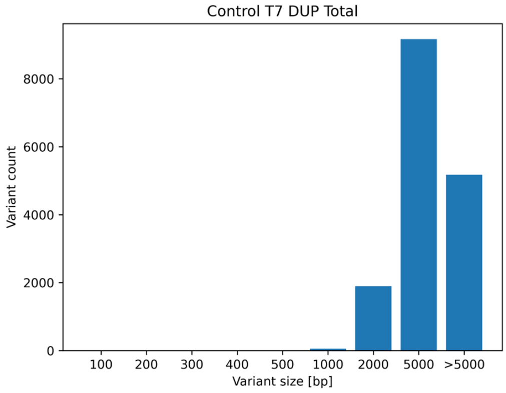


F)

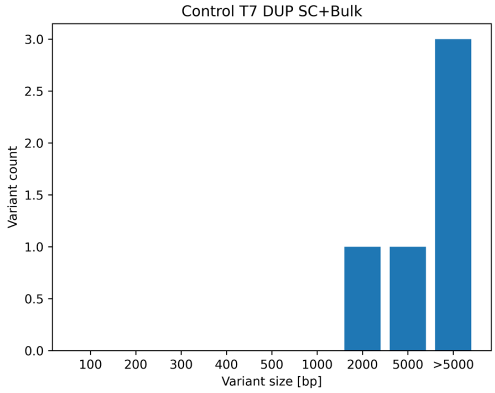


G)

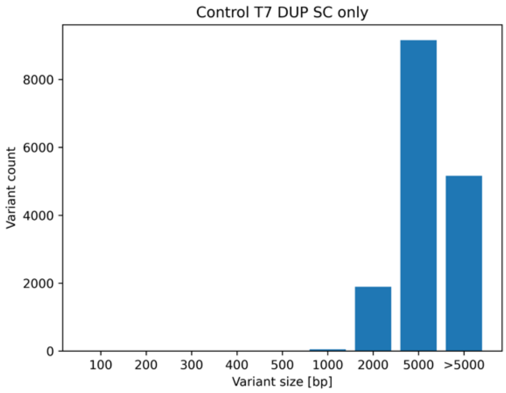


H)

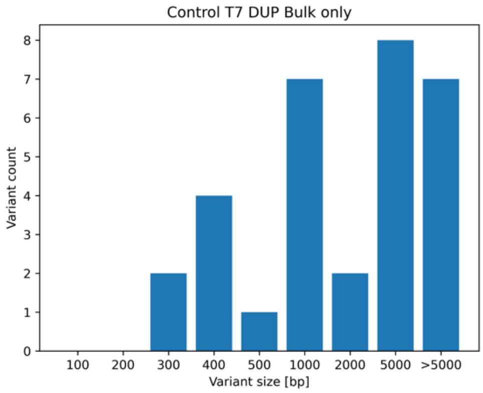

#### 52. Inversion and duplication sizes - unfiltered - MSA2 RBP

A)

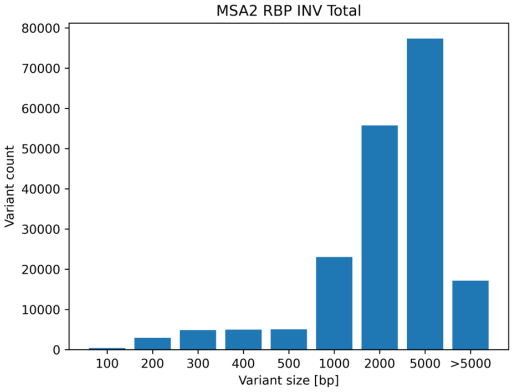


B)

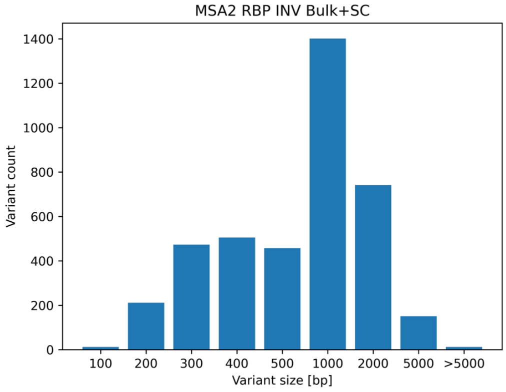


C)

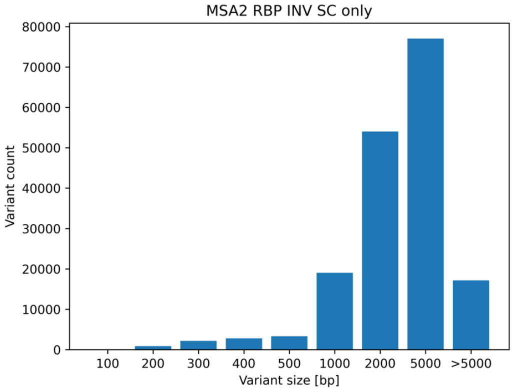


D)

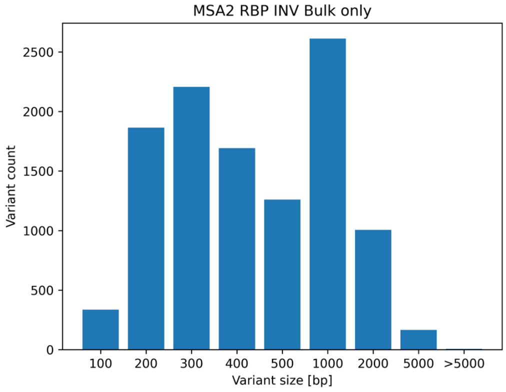


E)

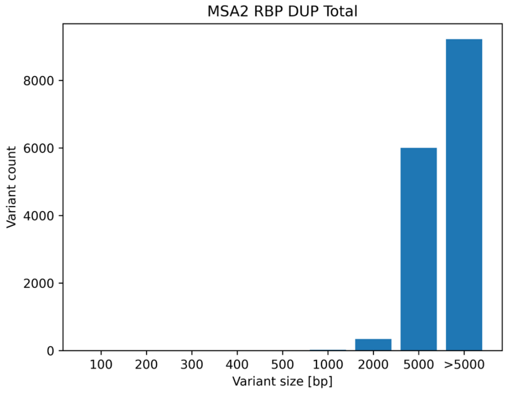


F)

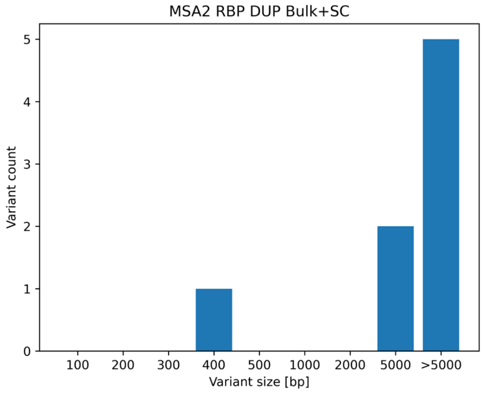


G)

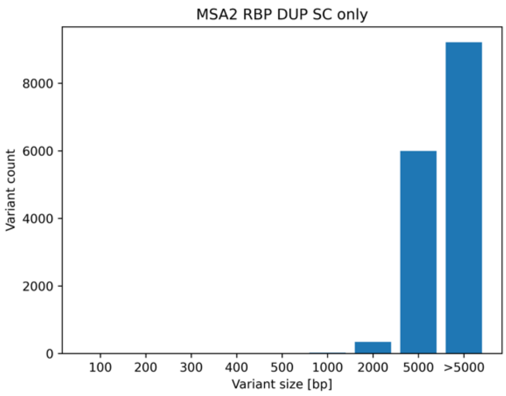


H)

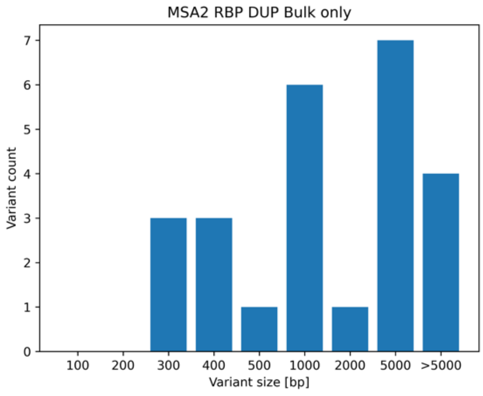

53. Inversion and duplication sizes - unfiltered - MSA2 T7

A)

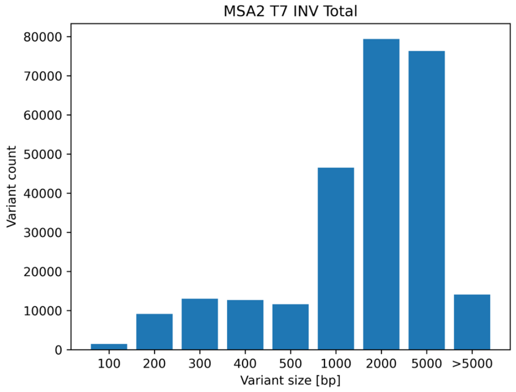


B)

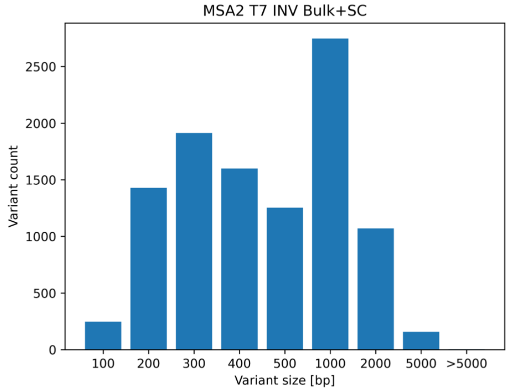


C)

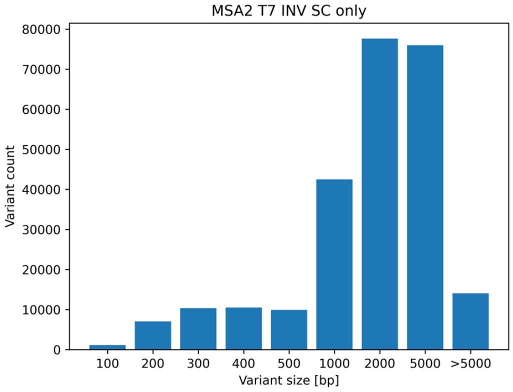


D)

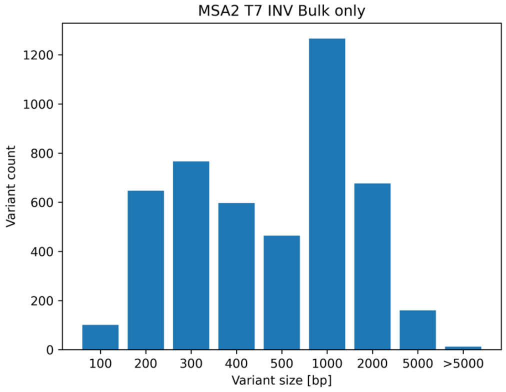


E)

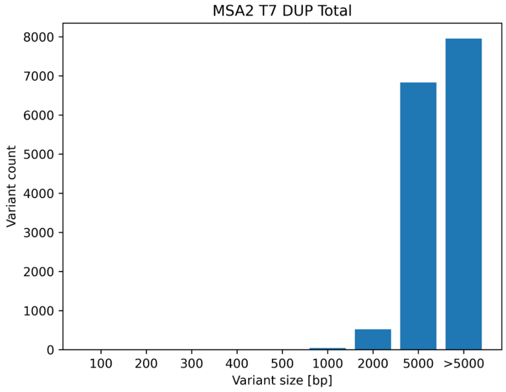


F)

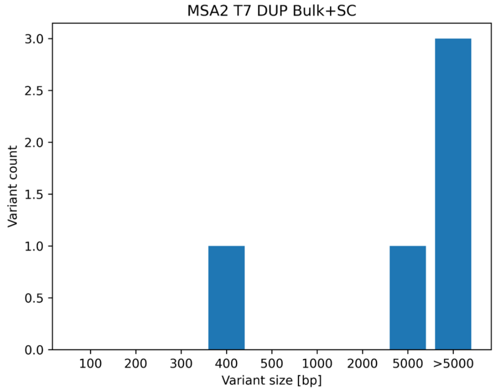


G)

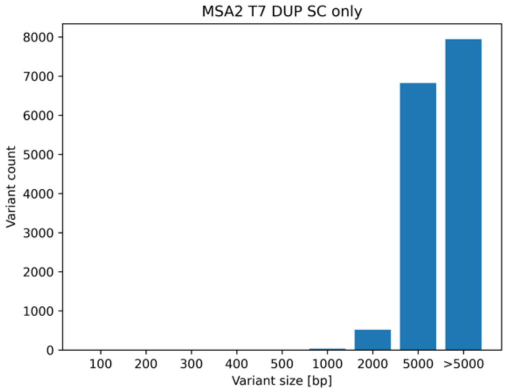


H)

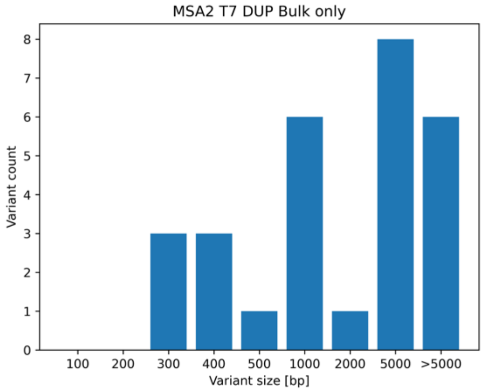

54. Variant size distribution - MSA1 T7 insertions.

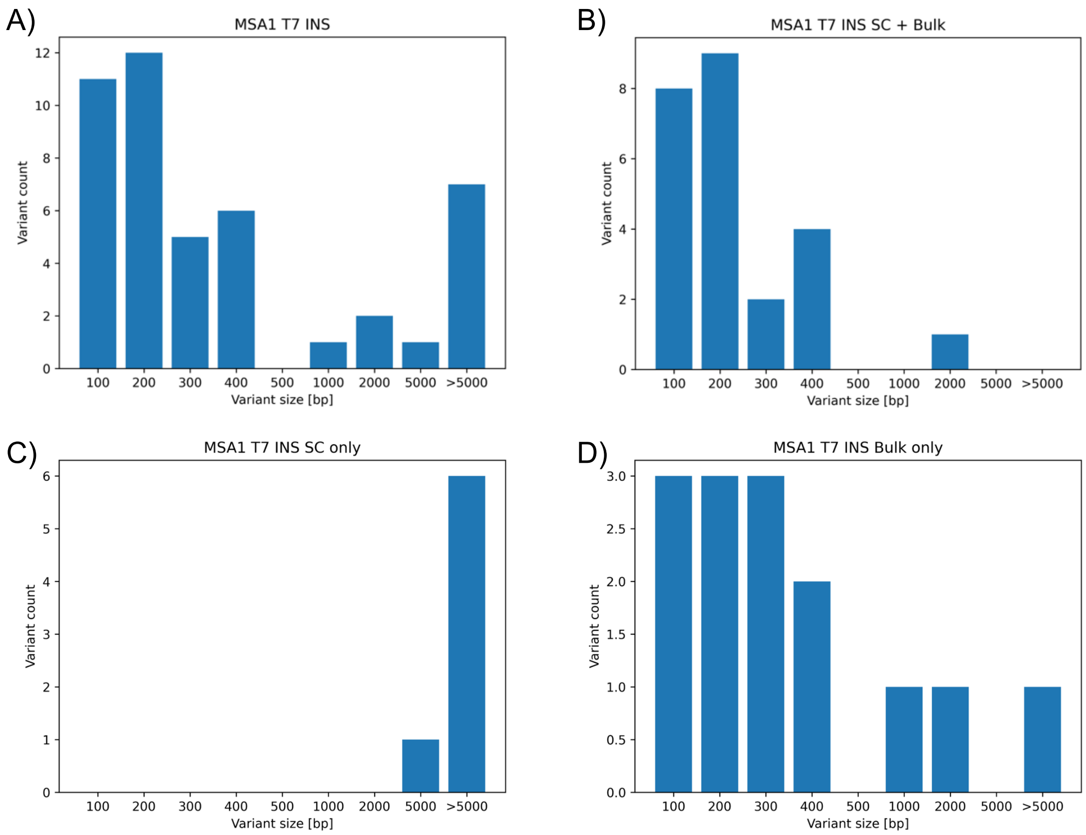


55. Variant size distribution - MSA1 T7 deletions. We report no SC-only deletions.

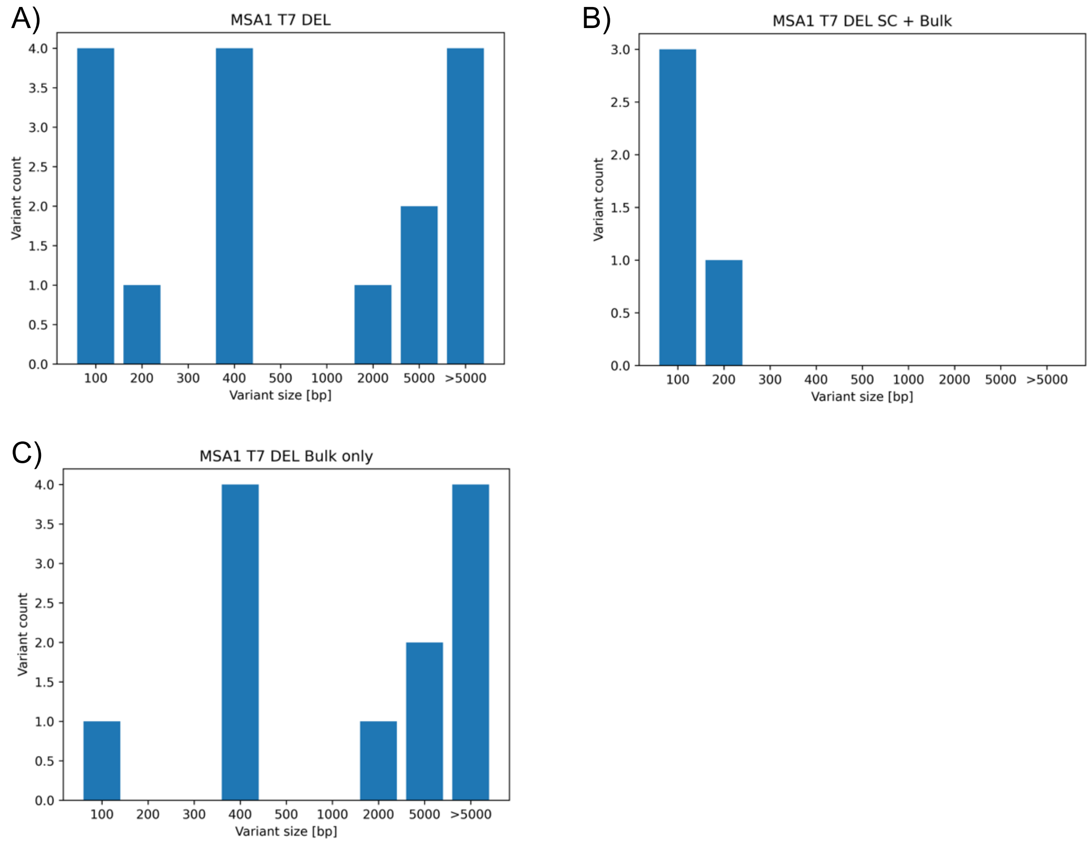

#### 56. Variant size distribution - MSA1 RBP insertions.

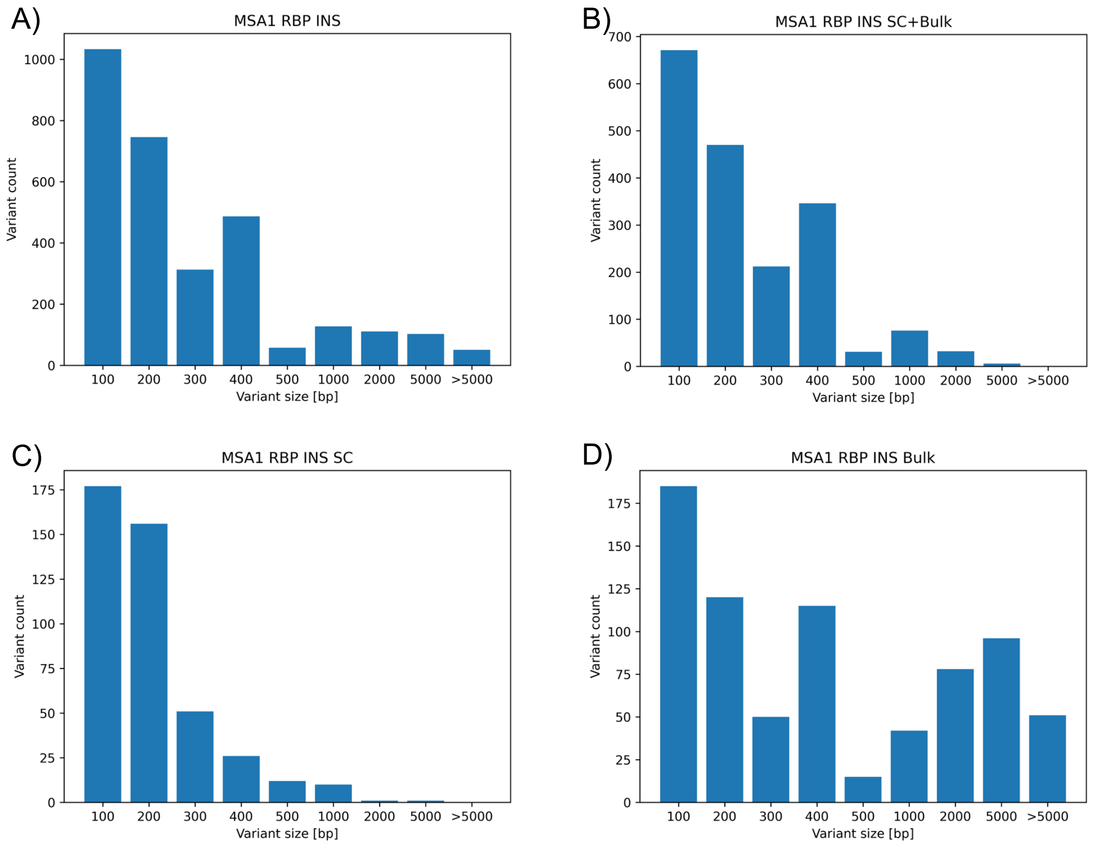


#### 57. Variant size distribution - MSA1 RBP deletions.

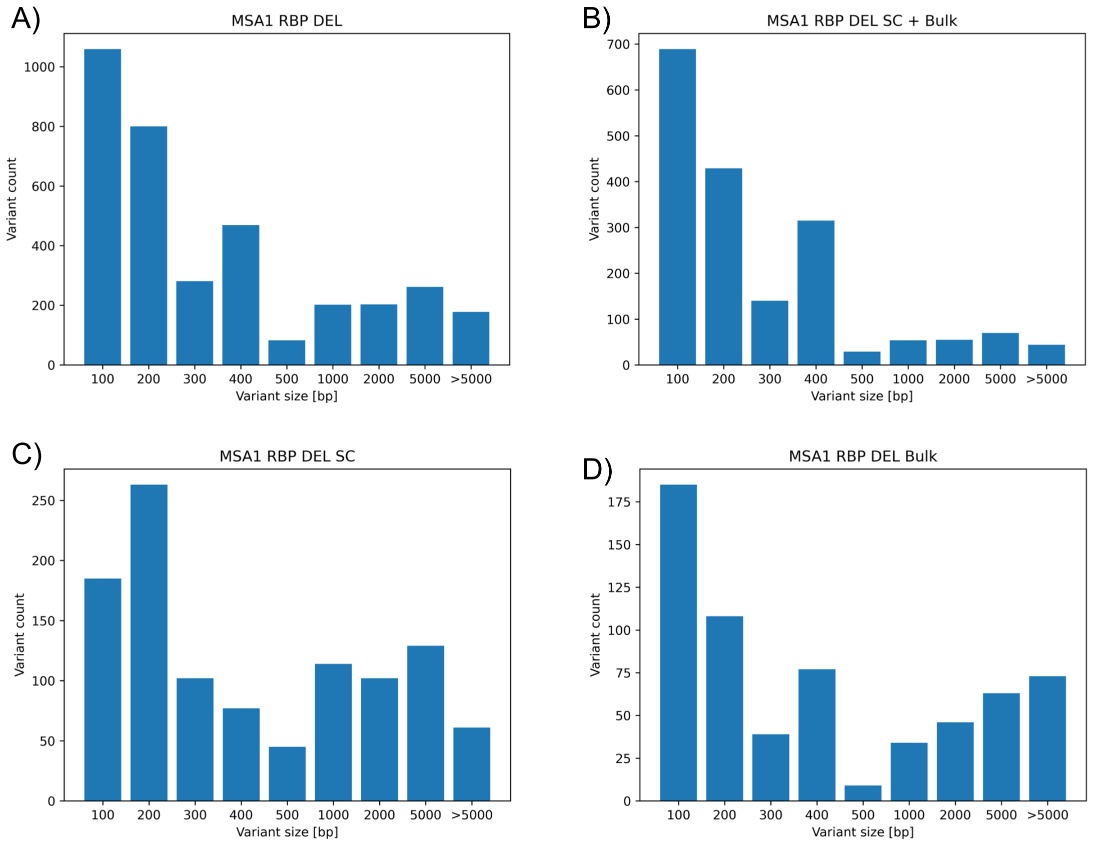

58. Variant size distribution - MSA2 RBP insertions.

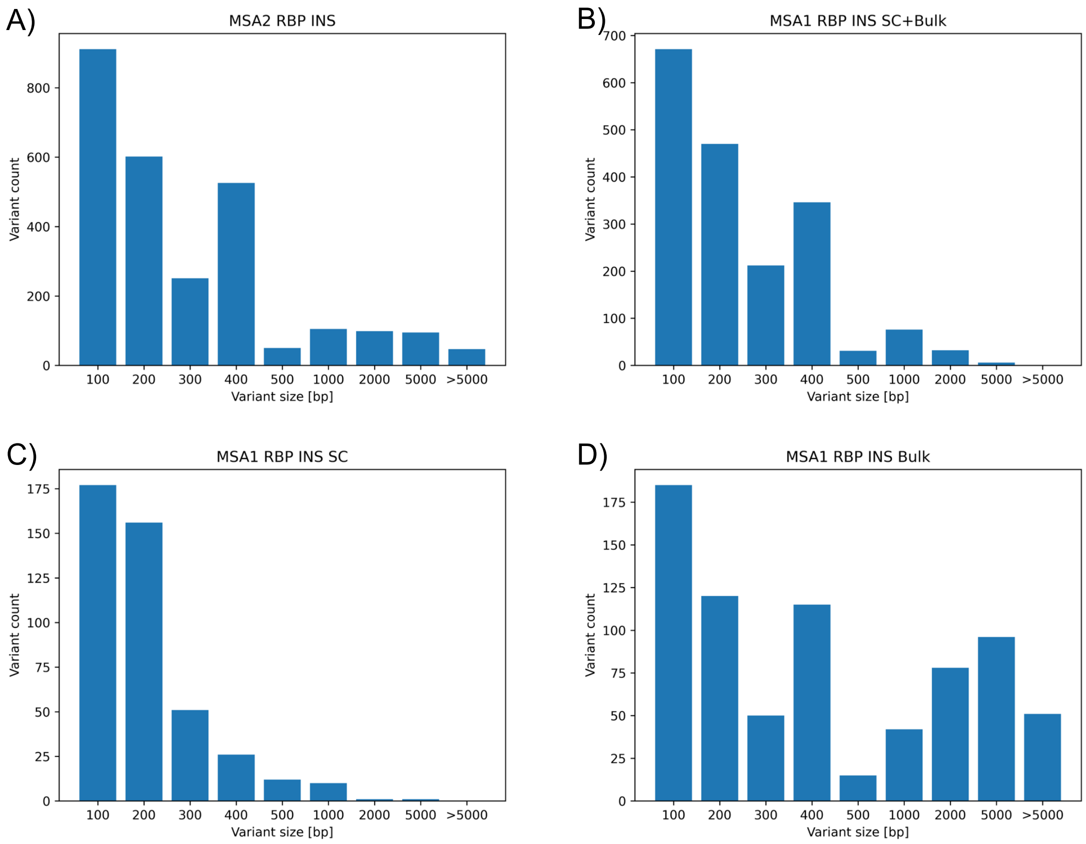


59. Variant size distribution - MSA2 RBP deletions.

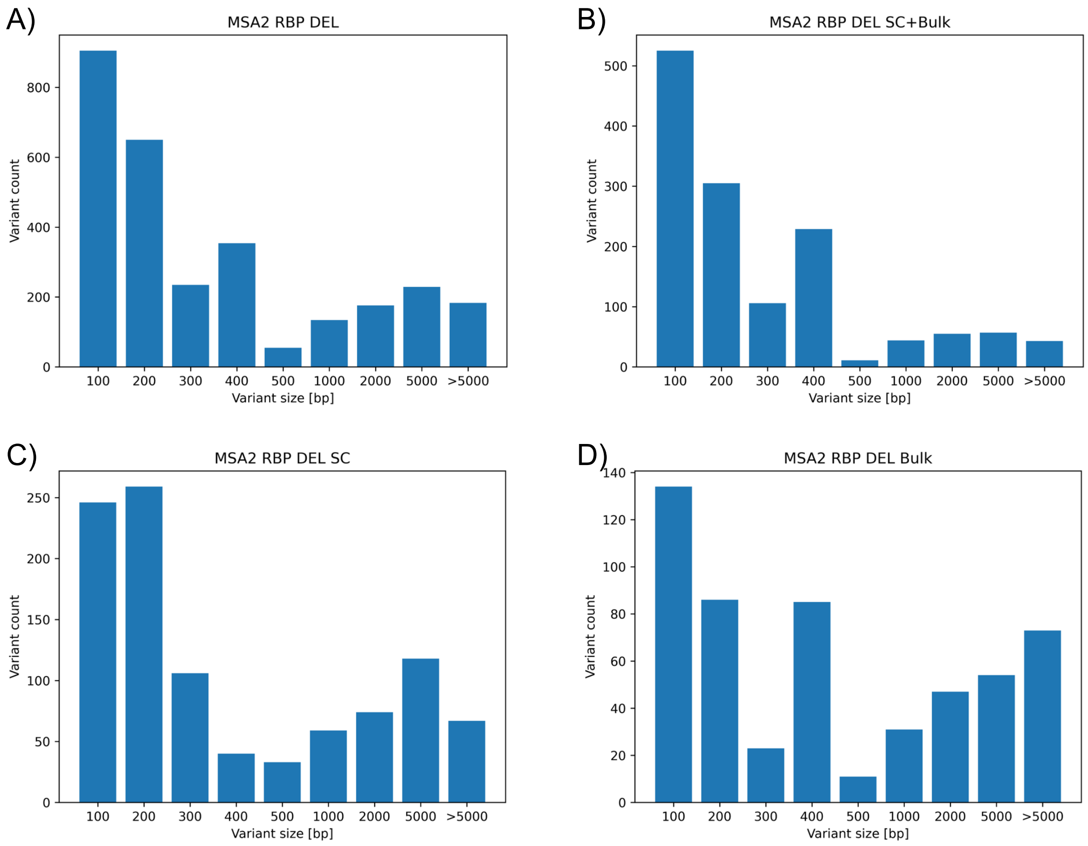

60. Variant size distribution - MSA2 T7 insertions.

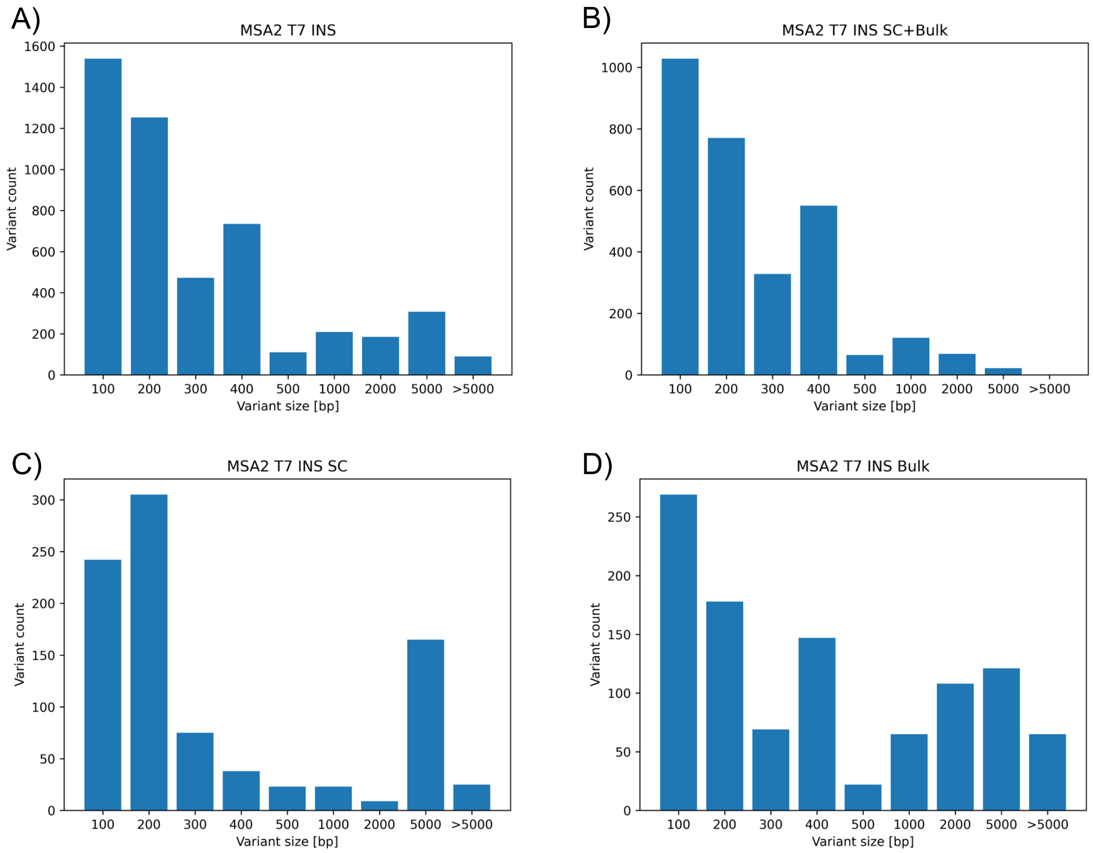


61. Variant size distribution - MSA2 T7 deletions.

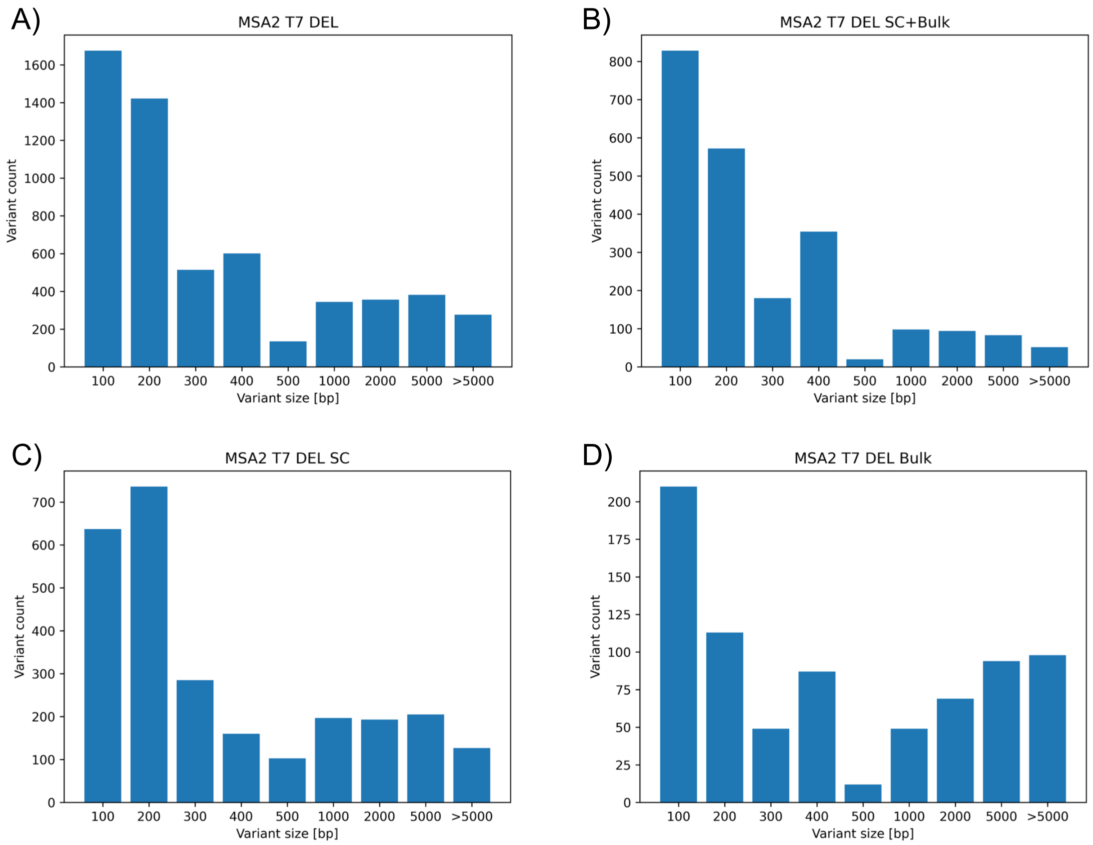

62. Variant size distribution - Control RBP insertions

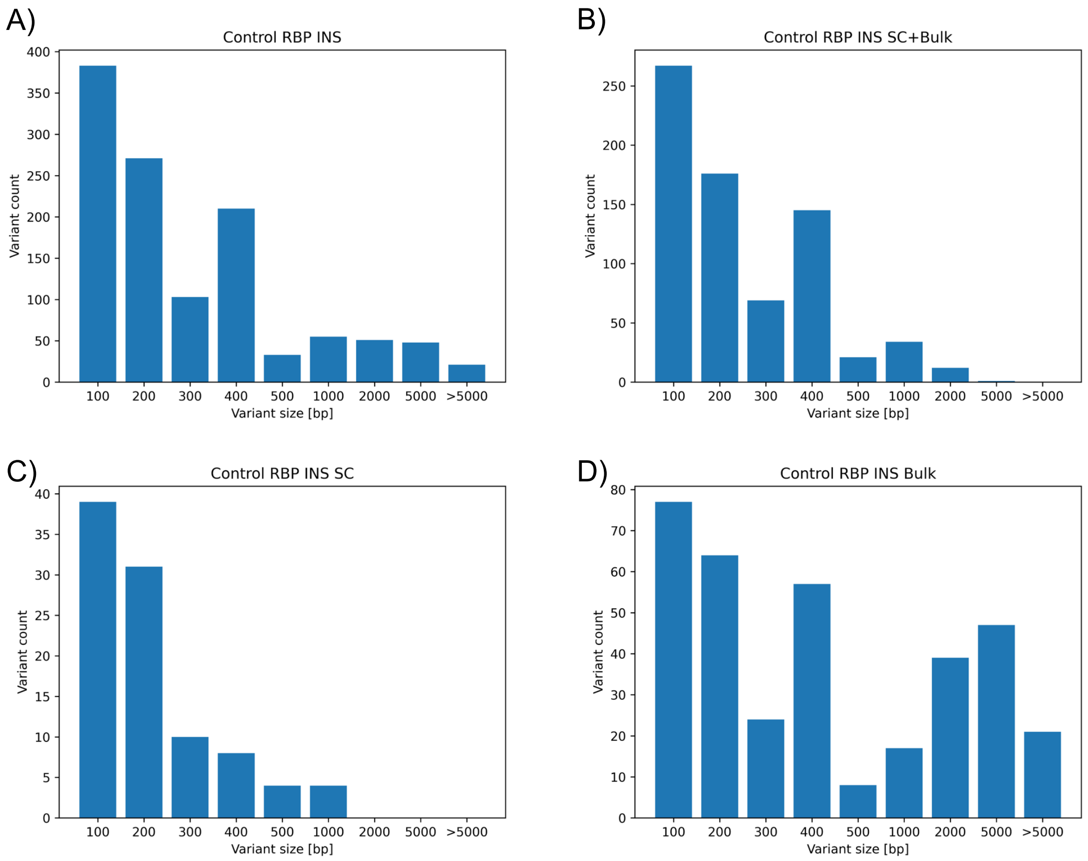


63. Variant size distribution - Control RBP deletions.

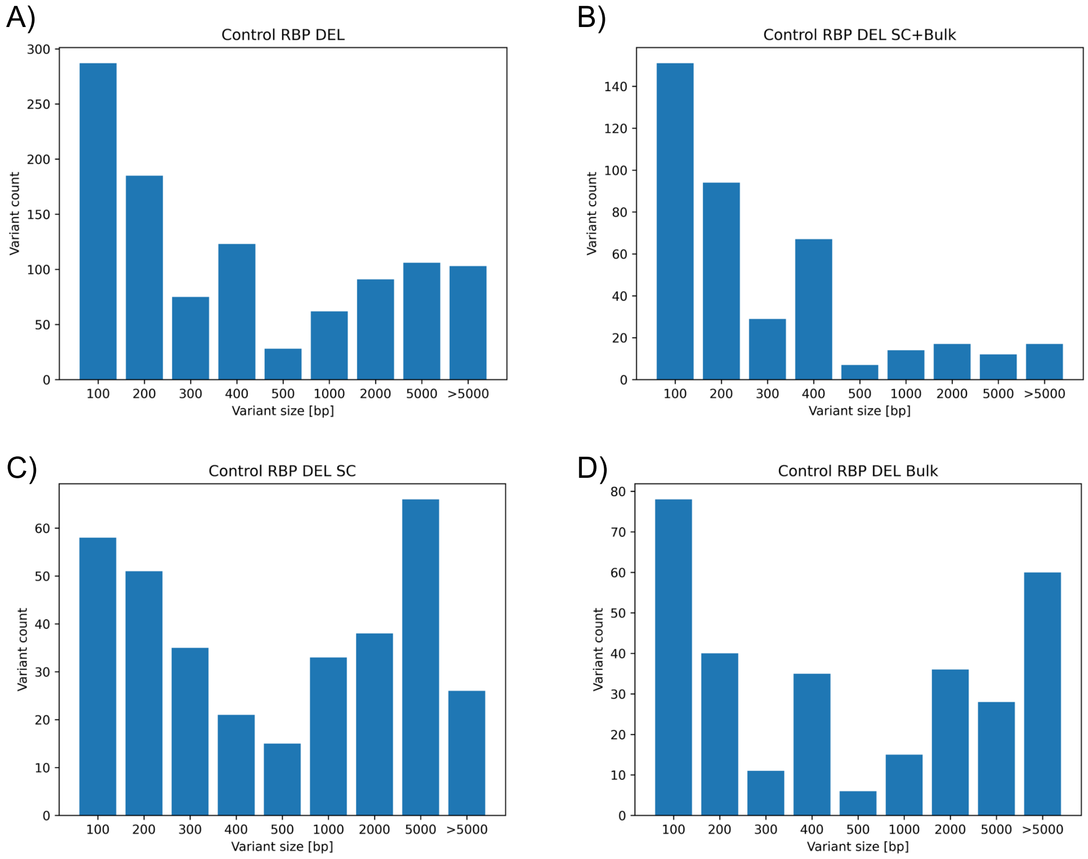

#### 64. Variant size distribution - Control T7 insertions.

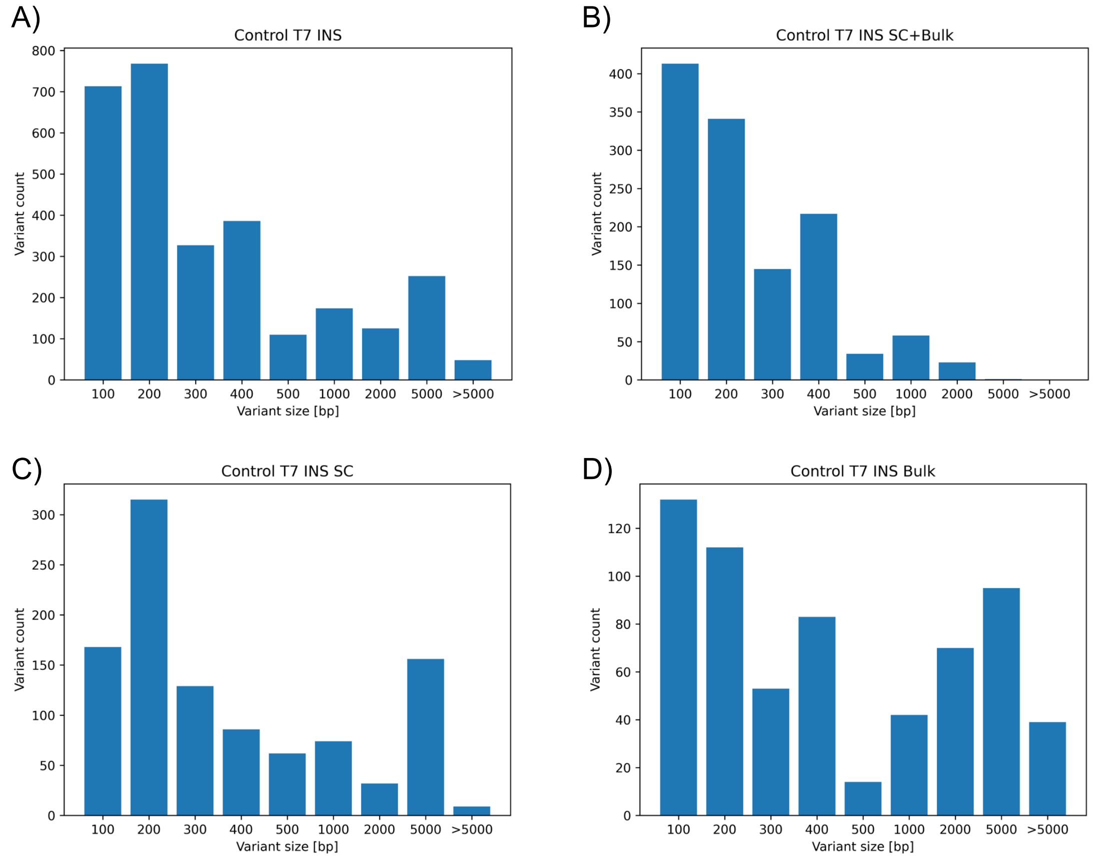


#### 65. Variant size distribution - Control T7 deletions.

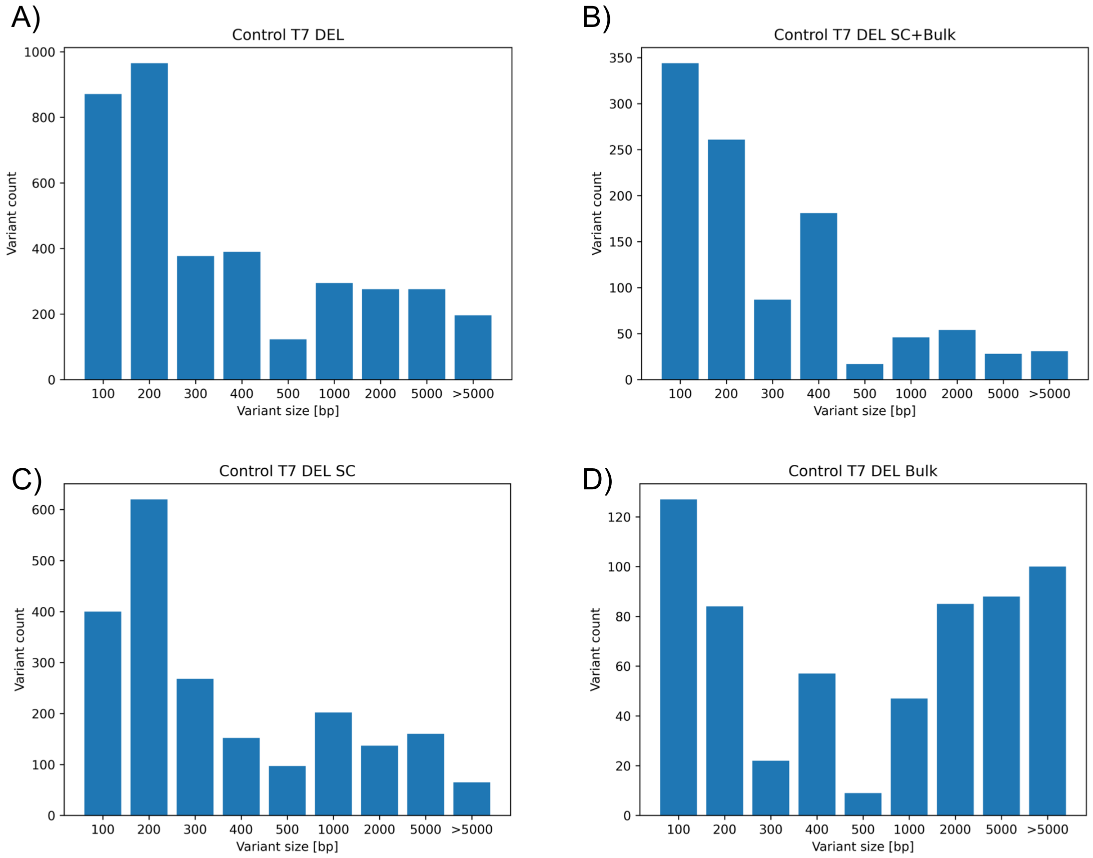

66. Coverage comparison between MSA2 MinION RBP and PromethION T7 experiments.

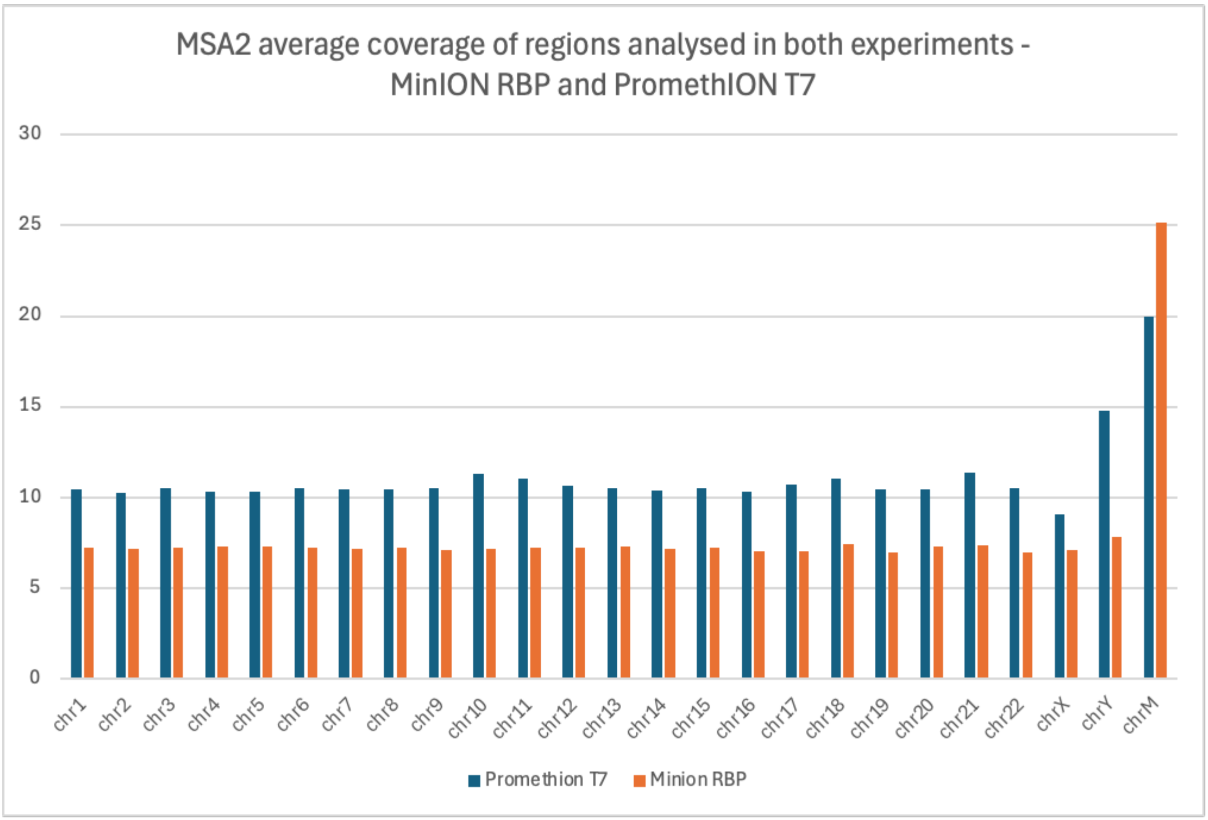


67. Coverage comparison between control MinION RBP and PromethION T7 experiments.

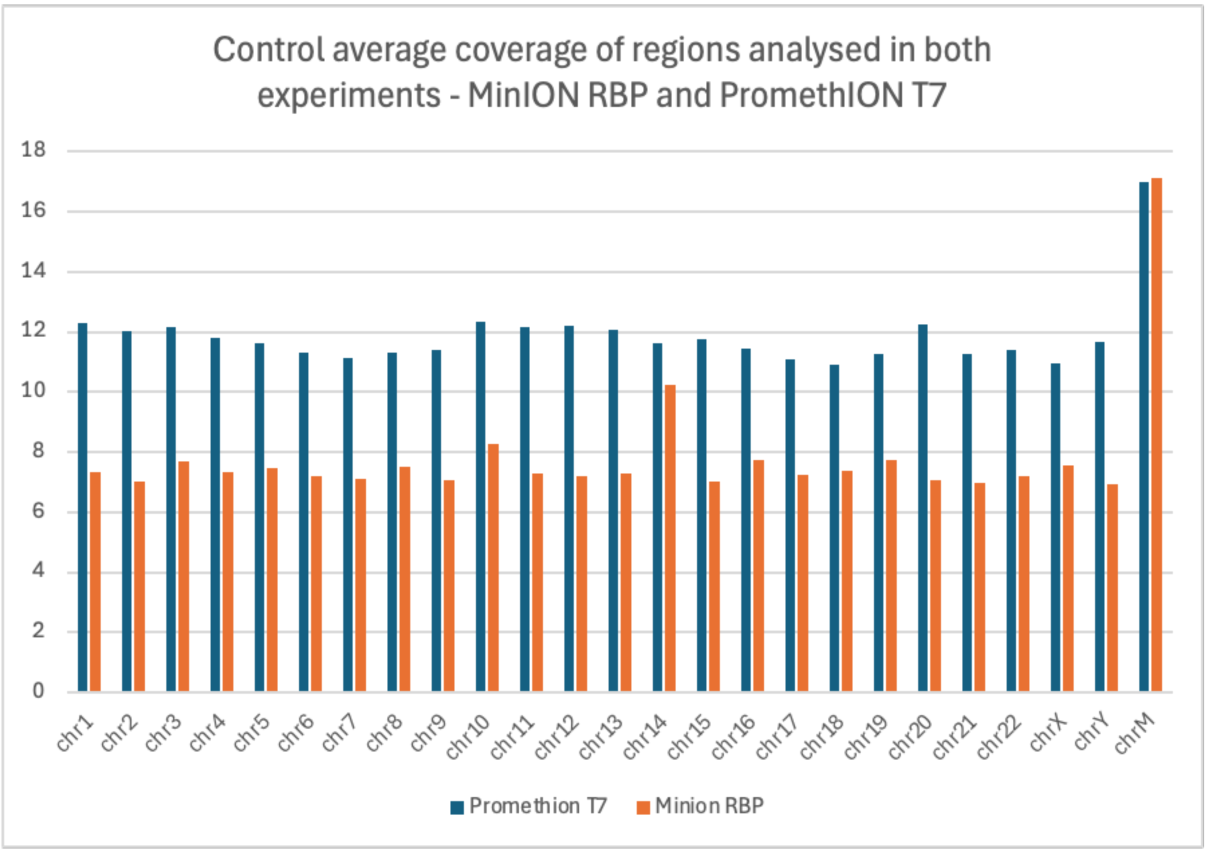

68. Examples of chimeric reads observed in MSA brains.

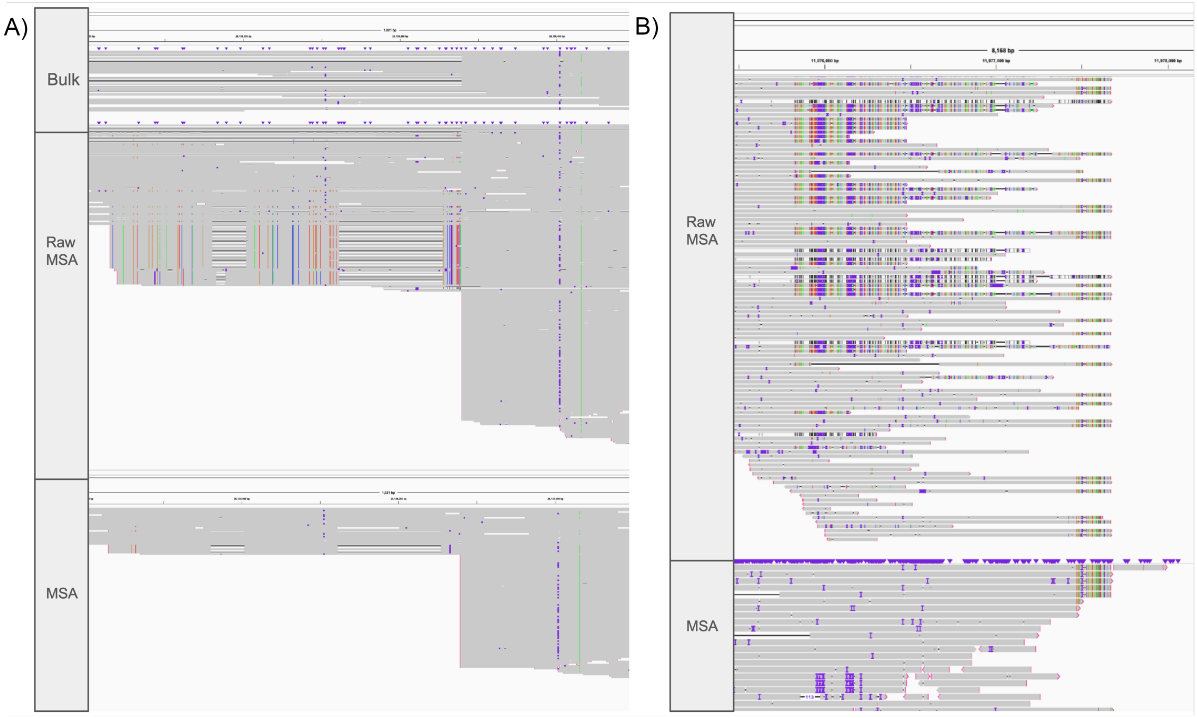
